# Antivirals for treatment of non-severe influenza: a systematic review and network meta-analysis of randomized controlled trials

**DOI:** 10.1101/2024.05.28.24307936

**Authors:** Ya Gao, Yunli Zhao, Ming Liu, Shuyue Luo, Yamin Chen, Xiaoyan Chen, Qingyong Zheng, Jianguo Xu, Yanjiao Shen, Wanyu Zhao, Zhifan Li, Sha Huang, Jie Huang, Jinhui Tian, Gordon Guyatt, Qiukui Hao

**Affiliations:** Evidence-Based Medicine Center, School of Basic Medical Sciences, Lanzhou University, Lanzhou, China; Department of Health Research Methods, Evidence, and Impact, McMaster University, Hamilton, ON, Canada; Department of Geriatric Medicine, The Second Affiliated Hospital of Chongqing Medical University, Chongqing, China; Chongqing Municipality Clinical Research Center for Geriatrics, The Second Affiliated Hospital of Chongqing Medical University, Chongqing, China; National Clinical Research Centre for Geriatrics, West China Hospital, Sichuan University, Chengdu, China; Center of Gerontology and Geriatrics, West China Hospital, Sichuan University, Chengdu, China; Clinical Nursing Teaching and Research Section, The Second Xiangya Hospital, Central South University, Changsha, China; Xiangya School of Nursing, Central South University, Changsha, China; Department of Geriatric, Zigong Affiliated Hospital of Southwest Medical University, Zigong, China; Chinese Evidence-Based Medicine Center, West China Hospital, Sichuan University, Chengdu, Sichuan, China; The First Clinical Medical College of Lanzhou University, Lanzhou, China; Department of Medicine, McMaster University, Hamilton, ON, Canada; School of Rehabilitation Science, McMaster University, Hamilton, ON, Canada

**Keywords:** Non-severe influenza, Antiviral treatment, Efficacy, Safety, Network meta-analysis

## Abstract

**Background:** The optimal antiviral drug for treatment of non-severe influenza remains unclear. To support an update of WHO guidelines on antiviral treatment for influenza, this systematic review compared effects of antiviral drugs for treating non-severe influenza.

**Methods:** We systematically searched Medline, Embase, Cochrane Central Register of Controlled Trials, Cumulative Index to Nursing and Allied Health Literature, Global Health, Epistemonikos, and ClinicalTrials.gov for randomized controlled trials published between database inception and 20 September 2023, comparing direct-acting influenza antiviral drugs, including but not limited to baloxavir, favipiravir, laninamivir, oseltamivir, peramivir, umifenovir, and zanamivir, to placebo, standard care, or another antiviral drug for treating people with non-severe influenza. We performed frequentist network meta-analyses to summarize the evidence and evaluated the certainty of evidence using the GRADE (Grading of Recommendations Assessment, Development and Evaluation) approach. We registered the protocol with PROSPERO, CRD42023456650.

**Findings:** We identified 11878 records, of which 73 trials with 34332 participants proved eligible. Compared with standard care or placebo, all antiviral drugs have little or no effect on mortality for low-risk patients (risk difference (RD) varied from 0.12 fewer to 0.02 fewer per 1000) and high-risk patients (RD varied from 1.22 fewer to 0.24 fewer per 1000) (all high certainty). All antivirals (no data for peramivir and amantadine) have little or no effect on admission to hospital (RD varied from 2 fewer to 1 more per 1000) for low-risk patients (high certainty). With respect to hospital admission, for high-risk patients, oseltamivir (RD 4 fewer per 1000, 95% CI 10 fewer to 4 more; high certainty) and zanamivir (RD 4 more per 1000, 95% CI 4 fewer to 15 more; high certainty) have little or no effect; baloxavir may reduce risk (RD 16 fewer per 1000, 95% CI 20 fewer to 4 more; low certainty); all other drugs may have little or uncertain effect. For time to alleviation of symptoms, baloxavir probably reduces symptom duration (mean difference (MD) 1.02 days lower, 95% CI 1.41 lower to 0.63 lower; moderate certainty); umifenovir may reduce symptom duration (MD 1.10 days lower, 95% CI 1.57 lower to 0.63 lower; low certainty); oseltamivir probably has no important effect (MD 0.75 days lower, 95% CI 0.93 lower to 0.57 lower, moderate certainty) and other drugs may have no important or little effect. For adverse events related to treatment, baloxavir (RD 32 fewer per 1000, 95% CI 52 fewer to 6 fewer; high certainty) has few or no such events; oseltamivir (RD 28 more per 1000, 95% CI 12 more to 48 more; moderate certainty) probably increases such events; other drugs may have little or no effect, or uncertain effect.

**Interpretation:** Baloxavir may reduce the risk of hospital admission for high-risk patients and probably reduces time to alleviation of symptoms, without increasing adverse events related to treatment in patients with non-severe influenza. All other antivirals either probably have little or no effect, or uncertain effects on patient-important outcomes.

**Funding:** WHO.

**Research in context:** *Evidence before this study:* Antiviral drugs may play a role in reducing illness duration, preventing serious complications, and lowering morbidity, particularly in high-risk populations. Previous systematic reviews and network meta-analyses have assessed the effects of antiviral drugs for treating influenza, but none assessed all approved antivirals for influenza or addressed patient-important outcomes of mortality and admission to hospital. The effect of many antiviral drugs for treating patients with non-severe influenza remains uncertain.

*Added value of this study:* This systematic review and network meta-analysis represents the most comprehensive assessment of the benefits and harms of antivirals in treating patients with non-severe influenza and demonstrates that baloxavir may reduce the risk of admission to hospital for high-risk patients and probably reduces time to alleviation of symptoms, does not increase adverse events related to treatment, but may increase emergence of resistance. Oseltamivir has little or no effect on mortality and admission to hospital, probably has no important effect on time to alleviation of symptoms, and probably increases adverse events related to treatments. Other antivirals probably have little or no effect on mortality and admission to hospital and may have no important effect on time to alleviation of symptoms.

*Implications of all the available evidence:* Our study provides evidence that baloxavir may be superior to standard care or placebo in reducing the risk of admission to hospital for high-risk patients and probably decreases time to alleviation of symptoms with few or no adverse effects. These findings support the use of baloxavir for treatment of high-risk non-severe influenza patients.

## Introduction

Influenza, a viral respiratory disease caused by influenza virus, affects people worldwide.^1^ Although most people with influenza have upper respiratory symptoms that are self-limited and recover within a week without medical attention, some individuals experience severe or fatal complications.^2,3^ This number is substantial: each year, influenza is estimated to cause 3 to 5 million cases of severe illness and 290,000 to 650,000 respiratory deaths worldwide.^3,4^

Antiviral drugs such as neuraminidase inhibitors (e.g., oseltamivir, zanamivir, laninamivir, and peramivir) and selective cap-dependent endonuclease inhibitors (e.g., baloxavir) may play a role in reducing illness duration, preventing serious complications, and lowering morbidity and mortality, particularly in high-risk populations.^5,6^ In 2022, the World Health Organization (WHO) published guidelines for clinical management of severe illness caused by influenza virus infections. These guidelines conditionally recommended the use of oseltamivir but advised against using inhaled zanamivir, inhaled Ianinamivir, or intravenous peramivir for individuals suspected or confirmed to have influenza virus infection with or at risk of severe illness.^7^ Most of the evidence supporting these recommendations was, however, of low or very low certainty, leaving optimal management in doubt.

Previous systematic reviews and network meta-analyses have assessed the effects of antiviral drugs for treating influenza^8–10^, but have been limited in failure to provide absolute effects of interventions,^8–10^ in their focus solely on specific categories of interventions such as neuraminidase inhibitors or particular subtypes of influenza such as seasonal influenza,^8–10^ or overlooked crucial patient-important outcomes including mortality and hospitalization.^8–10^ Additionally, some studies^9,10^ do not assess the certainty of the evidence and none include recent trials.^11–14^

To evaluate the effectiveness and safety of antiviral treatment for patients with non-severe influenza and thus support an update of WHO guidelines on antiviral treatment for influenza,^7^ we performed a systematic review and network meta-analysis of randomized controlled trials (RCTs).

## Methods

### Registration and reporting

We registered this systematic review protocol with PROSPERO (CRD42023456650) and reported the review according to the guideline of Preferred Reported Items for Systematic Reviews and Meta-Analyses (PRISMA) for network meta-analyses.^15^

### Search strategy and selection criteria

With the aid of a medical librarian, we searched Medline, Embase, Cochrane Central Register of Controlled Trials (CENTRAL), Cumulative Index to Nursing and Allied Health Literature (CINAHL), Global Health, Epistemonikos, ClinicalTrials.gov from databases inception to 20 September 2023 (Appendix 1). To identify additional trials, we manually searched reference lists of relevant previous systematic reviews and eligible studies.

We included RCTs that compared direct-acting influenza antiviral drugs to placebo, standard care, or another antiviral drug for people with suspected or laboratory-confirmed (by RT-PCR assay, rapid antigen test, or immunofluorescence assay) non-severe influenza. We used the WHO definitions for disease severity; severe illness from influenza infection was defined as an illness that requires hospitalisation.^7^ We defined non-severe influenza as the absence of any criteria for severe influenza and included RCTs enrolling naturally infected patients with any etiology of influenza virus, including seasonal influenza viruses, pandemic influenza viruses, and zoonotic influenza viruses.

We focused on influenza antiviral drugs approved by the US Food and Drug Administration (FDA) or approved for use in different parts of the world.^5^ Eligible antiviral drugs included baloxavir, oseltamivir, laninamivir, zanamivir, peramivir, umifenovir, favipiravir, amantadine, and rimantadine.

We did not impose any restrictions on publication language, patient age, or dose and administration route of antiviral drugs. Eligible trials reported at least one of the outcomes of interest. We excluded studies investigating the antiviral effects of Traditional Chinese medicines and antiviral drugs combined with adjunctive therapies as well as trials that used antivirals as prophylaxis against influenza in individuals who had been exposed to influenza virus.

Using Covidence (https://covidence.org/), pairs of reviewers independently screened titles and abstracts of all records and full texts of potentially eligible records. We checked the retractions for all eligible publications, if the study was retracted from the publication, we excluded the study from our review.^16^ Pairs of reviewers independently extracted data on study characteristics, patient characteristics, characteristics of interventions, and outcomes (Appendix 2). Reviewers resolved discrepancies by discussion or, if necessary, with the assistance of a third party for adjudication.

### Data analysis

Patient-important outcomes of interest to the guideline panel included mortality, admission to hospital, admission to intensive care unit (ICU), progression to mechanical ventilation, duration of mechanical ventilation, duration of hospitalization, time to alleviation of symptoms, hospital discharge destination, emergence of resistance, any adverse events, adverse events related to treatments, and serious adverse events. We defined time to alleviation of symptoms as time from the start of treatment to the alleviation of all influenza-associated symptoms.^17,18^

We conducted pairwise meta-analyses for all direct comparisons of each outcome using the Hartung-Knapp-Sidik-Jonkman (HKSJ) random-effects model in R version 4.2.1 (R Foundation for Statistical Computing). To assess the between-study heterogeneity, we visually inspected forest plots and used the I^2^ statistic. For comparisons that included at least 10 studies, to assess publication bias, we used Harbord’s test for dichotomous outcomes and Egger’s test for continuous outcomes,^19,20^ as well as a visual assessment of the funnel plot.

For emergence of resistance, because no such events occur in standard care or placebo groups, to estimate pooled emergence of resistance rates for each antiviral, we conducted meta-analyses of the proportion of actively treated patients in whom resistance occurred using the Hartung-Knapp-Sidik-Jonkman (HKSJ) random effects model with the restricted maximum likelihood (REML) heterogeneity estimator method.^21,22^ To stabilize variances, we applied the Freeman-Tukey double arcsine transformation.^23^

We drew network plots for each outcome using STATA 15.0 (StataCorp, College Station, Texas, USA). Using netmeta package in R version 4.2.1 (R Foundation for Statistical Computing), we performed frequentist random-effects network meta-analyses with a graph-theoretical approach.^24^ The estimator was based on weighted least-square regression using the Moore-Penrose pseudoinverse method.^24^ For each outcome, we used the “design-by-treatment” model (global test) to assess the coherence assumption for the entire network.^25^ We calculated indirect estimates from the network by node-splitting and a back-calculation method.^26^ To evaluate local (loop-specific) incoherence in each closed loop of the network - the difference between direct and indirect evidence - we used the node-splitting method and calculated a P-value for the test of incoherence.^27^

We calculated risk ratios (RRs) with 95% confidence intervals (CIs) for mortality, admission to hospital, any adverse events, and adverse events related to treatments. For mortality and hospital admission, we applied the continuity correction of 0.5 for trials with 0 events.^28^ We calculated absolute effects using the network RR estimates and the baseline risk estimates. To estimate absolute effects of antivirals on mortality and hospital admission, the guideline panel recommended use of two baseline risk categories for patients at low risk and high risk for severe complications. We defined high-risk patients using the definitions from the 2022 WHO guidelines.^7^ For mortality, we obtained baseline risks for low-risk (0.2 per 1000) and high-risk (2 per 1000) patients from observational studies.^29,30^ For other outcomes for which reliable observational data were not available, we used the median baseline risk in the standard care or placebo group of eligible RCTs.

For ICU admission and serious adverse events, the median event rate from the standard care or placebo arm was less than 1%, we directly calculated the absolute risk differences (RDs) with 95% CIs. We calculated mean differences (MDs) with 95% CIs for continuous outcomes. When standard deviations (SDs) were missing, we estimated them using the methods described in the Cochrane Handbook.^31^

If at least two trials provided relevant information for each subgroup, we performed the following prespecified within-trial subgroup analyses: seasonal versus zoonotic versus pandemic influenza viruses; confirmed versus suspected influenza virus infection; children < 2 years, children versus adults and adolescents versus elderly (≥ 65 years); patients at increased risk of severe complications versus at low risk (Appendix 2 presents further details). We planned to assess the credibility of the subgroup hypothesis using the Instrument for assessing the Credibility of Effect Modification Analyses (ICEMAN) tool.^32^ Pairs of reviewers independently evaluated the risk of bias of eligible RCTs using a modified Cochrane risk of bias tool (Appendix 2).^33^

To assess the certainty of evidence for each outcome, we used the Grading of Recommendations Assessment, Development and Evaluation (GRADE) approach for network meta-analysis.^34,35^ We evaluated the risk of bias, imprecision, inconsistency, indirectness, and publication bias, and rated the certainty of evidence for direct estimates as high, moderate, low, or very low.^36^ We assessed imprecision at the network level using the minimally important difference (MID) as a threshold.^37^ The guideline panel specified an MID as 0.3% for mortality, 1.5% for admission to hospital, 1% for admission to ICU, 1% for any adverse events and adverse events related to treatments, 0.5% for serious adverse events, 5% for emergence of resistance, and 1 day for time to alleviation of symptoms and duration of hospitalization. We rated imprecision following GRADE guidance (Appendix 2 presents further details).^38,39^ We developed the summary of findings tables for comparisons of each antiviral versus standard care or placebo in MAGICapp following GRADE guidance.^40,41^

### Role of the funding source

The funder had no role in study design, data collection, analysis, and interpretation, or writing of the manuscript and the decision to submit.

## Results

The electronic search identified 11878 records. After screening 8944 unique titles and abstracts and 459 full texts, 87 articles reporting 73 unique RCTs proved eligible (Figure 1). Appendix 3 lists the eligible trials.

**Figure 1.**
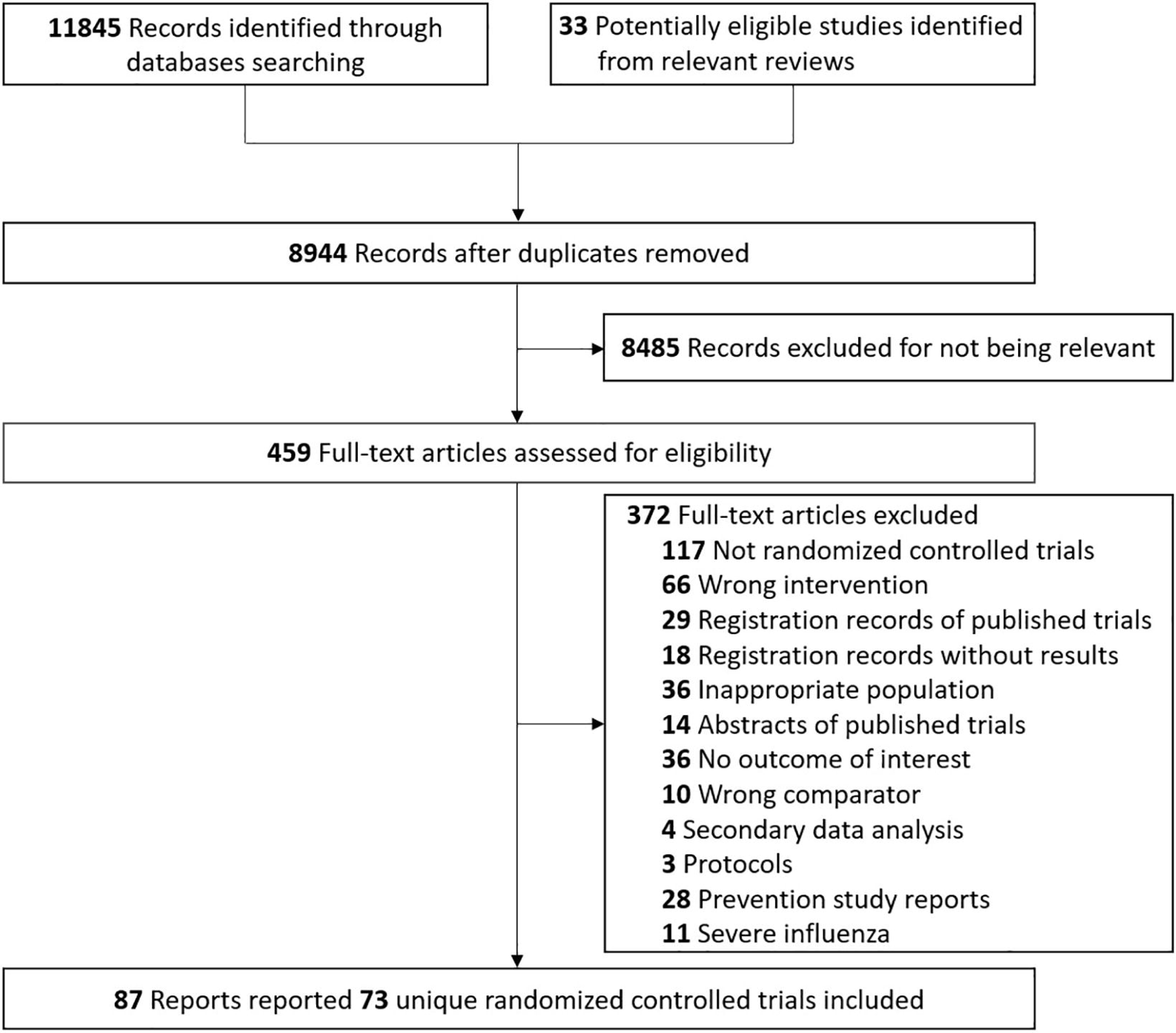
Study selection

Eligible trials were published from 1971 through 2023 and evaluated eight unique antivirals: baloxavir, favipiravir, laninamivir, oseltamivir, peramivir, umifenovir, zanamivir, and amantadine, with the most common comparison being between oseltamivir and standard care or placebo (Appendix 4). Sample sizes ranged from 14 to 3266 (a total of 34332); the median of mean age was 35.0 years, and the median proportion of men was 49.8%. The follow-up for outcomes ranged from 5 to 29 days.

Appendix 5 presents the risk of bias of eligible trials for each outcome. Nine trials were at low or probably low risk of bias for all reported outcomes. The main limitations were inadequate allocation concealment and the lack of blinding of data collectors, outcome assessors, and data analysts.

Figure 2 and Appendix 6 present network plots for each outcome. We did not find substantial between-study heterogeneity for most outcomes and most comparisons (Appendix 7). Tests of incoherence raised no concerns of global incoherence (Appendix 8) for all outcomes and no concerns of local incoherence (Appendix 9) between direct and indirect evidence except for time to alleviation of symptoms. Appendi× 10 presents network relative estimates and absolute estimates for each comparison for outcomes, with certainty of evidence. Table 1, Table 2, and Appendix 11 present the GRADE summary of findings for antivirals versus standard care or placebo. We did not find evidence of intransitivity. We found no evidence of publication bias (Appendix 12).

**Figure 2.**
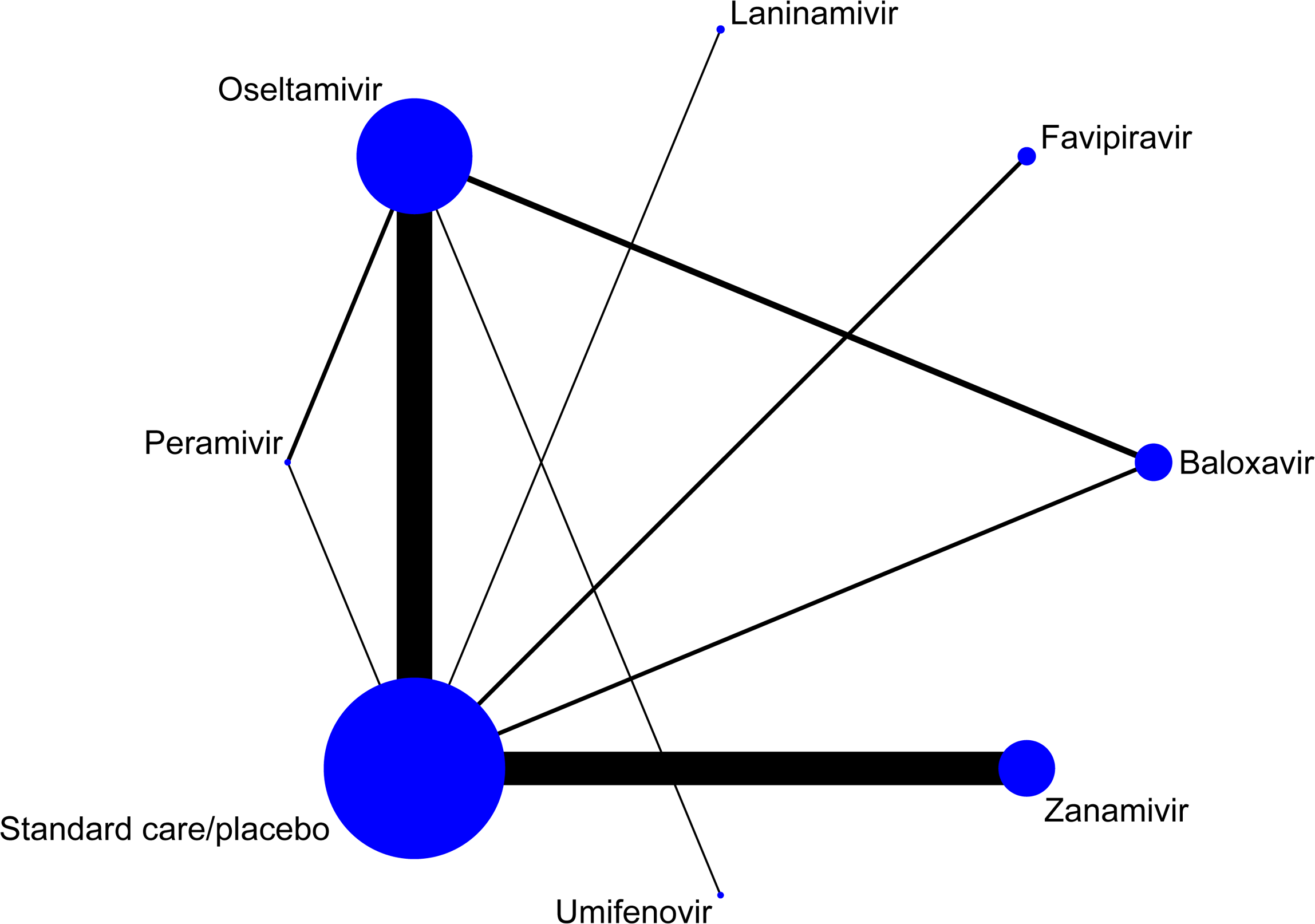
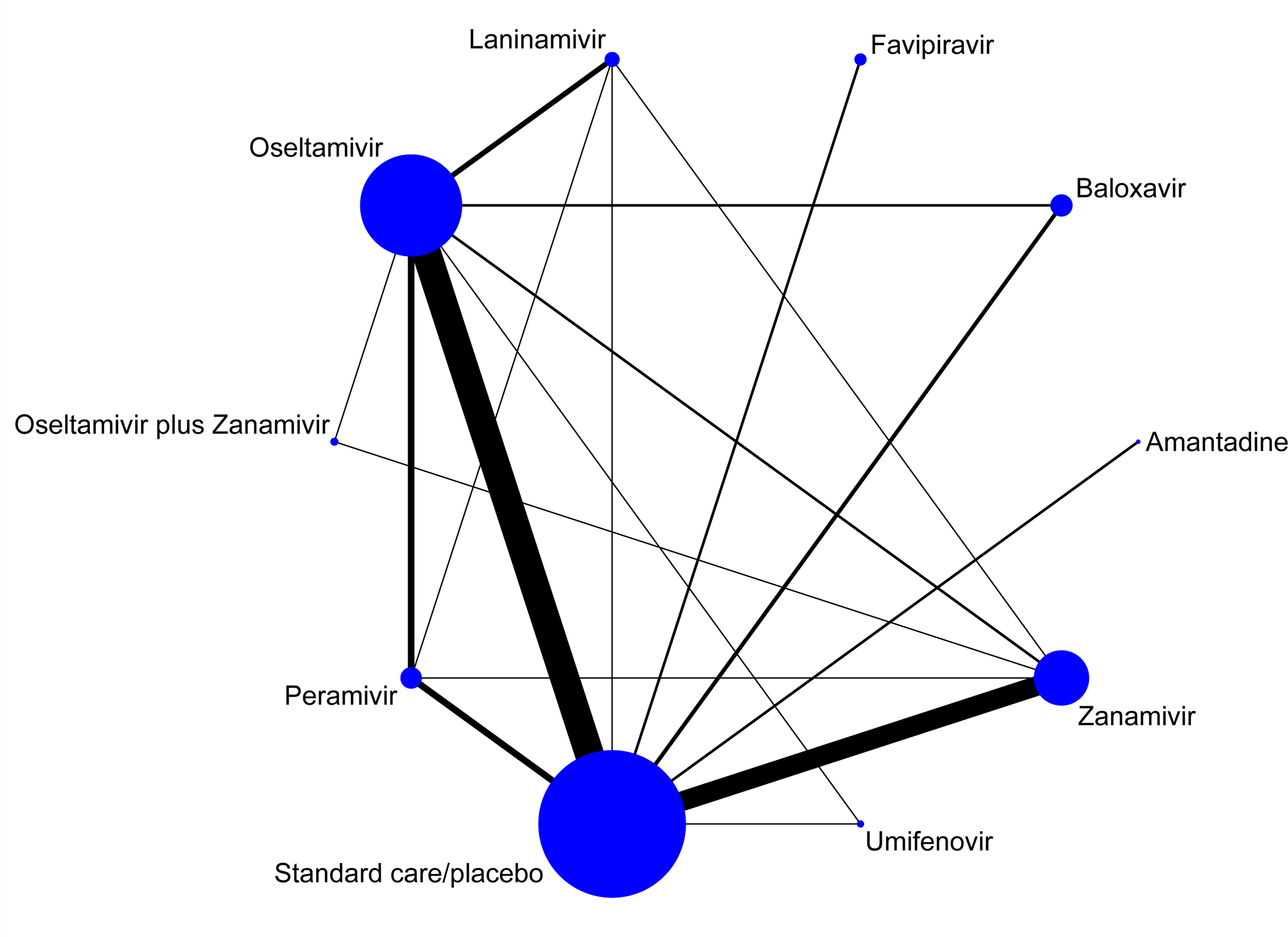
Network plot for mortality (A) time to alleviation of symptoms (B) Footnotes: The size of the circle represents the number of participants. The connecting lines represent direct comparisons. The width of the line represents the number of studies.

**Table 1.** Summary of findings table for baloxavir versus standard care or placebo.

| Outcome | Study results and measurements | Absolute effect estimates |  | Certainty of the Evidence<br>(Quality of evidence) | Summary |
| --- | --- | --- | --- | --- | --- |
|  |  | Standard care/placebo | Baloxavir |  |  |
| Mortality<br>(Low-risk) | Relative risk: 0.83<br>(CI 95% 0.14 - 4.82)<br>Based on data from 2144 participants in 2 studies | **0.20**<br>per 1000 | **0.17**<br>per 1000 | **High** | Baloxavir has little or no effect on mortality. |
|  |  | Difference: **0.03 fewer per 1000**<br>(CI 95% 0.17 fewer - 0.76 more) |  |  |  |
| Mortality<br>(High-risk) | Relative risk: 0.83<br>(CI 95% 0.14 - 4.82)<br>Based on data from 2144 participants in 2 studies | **2.0**<br>per 1000 | **1.66**<br>per 1000 | **High** | Baloxavir has little or no effect on mortality. |
|  |  | Difference: **0.34 fewer per 1000**<br>(CI 95% 1.72 fewer - 7.64 more) |  |  |  |
| Admission to hospital<br>(Low-risk) | Relative risk: 0.24<br>(CI 95% 0.05 - 1.19)<br>Based on data from 1461 participants in 2 studies | **3**<br>per 1000 | **1**<br>per 1000 | **High** | Baloxavir has little or no effect on admission to hospital. |
|  |  | Difference: **2 fewer per 1000**<br>(CI 95% 3 fewer - 1 more) |  |  |  |
| Admission to hospital<br>(High-risk) | Relative risk: 0.24<br>(CI 95% 0.05 - 1.19)<br>Based on data from 1461 participants in 2 studies | **21**<br>per 1000 | **5**<br>per 1000 | **Low**<br>Due to very serious imprecision | Baloxavir may reduce the risk of admission to hospital. |
|  |  | Difference: **16 fewer per 1000**<br>(CI 95% 20 fewer - 4 more) |  |  |  |
| Adverse events related to treatments | Relative risk: 0.74<br>(CI 95% 0.57 - 0.95)<br>Based on data from 2776 participants in 3 studies | **122**<br>per 1000 | **90**<br>per 1000 | **High** | Baloxavir does not increase adverse events related to treatments. |
|  |  | Difference: **32 fewer per 1000**<br>(CI 95% 52 fewer - 6 fewer) |  |  |  |
| Serious adverse events | Risk difference: 0.001<br>(CI 95% -0.004 – 0.005)<br>Based on data from 2776 participants in 3 studies | **5**<br>per 1000 | **6**<br>per 1000 | **Moderate**<br>Due to serious imprecision | Baloxavir probably has little or no effect on serious adverse events. |
|  |  | Difference: **1 more per 1000**<br>(CI 95% 4 fewer - 5 more) |  |  |  |
| Time to alleviation of symptoms | Measured by: day<br>Lower better<br>Based on data from 1855 participants in 3 studies | **4.92**<br>Mean | **3.90**<br>Mean | **Moderate**<br>Due to serious imprecision | Baloxavir probably reduces time to alleviation of symptoms. |
|  |  | Difference: **MD 1.02 lower**<br>(CI 95% 1.41 lower - 0.63 lower) |  |  |  |

**Table 2.** Summary of findings table for oseltamivir versus standard care or placebo.

| Outcome | Study results and measurements | Absolute effect estimates |  | Certainty of the Evidence<br>(Quality of evidence) | Summary |
| --- | --- | --- | --- | --- | --- |
|  |  | Standard care/placebo | Oseltamivir |  |  |
| Mortality<br>(Low-risk) | Relative risk: 0.84<br>(CI 95% 0.34 - 2.07)<br>Based on data from 12008 participants in 17 studies | 0.20<br>per 1000 | 0.17<br>per 1000 | High | Oseltamivir has little or no effect on mortality. |
|  |  | Difference: 0.03 fewer per 1000<br>(CI 95% 0.13 fewer - 0.21 more) |  |  |  |
| Mortality<br>(High-risk) | Relative risk: 0.84<br>(CI 95% 0.34 - 2.07)<br>Based on data from 12008 participants in 17 studies | 2.0<br>per 1000 | 1.68<br>per 1000 | High | Oseltamivir has little or no effect on mortality. |
|  |  | Difference: 0.32 fewer per 1000<br>(CI 95% 1.32 fewer - 2.14 more) |  |  |  |
| Admission to hospital<br>(Low-risk) | Relative risk: 0.8<br>(CI 95% 0.54 - 1.18)<br>Based on data from 12589 participants in 20 studies | 3<br>per 1000 | 2<br>per 1000 | High | Oseltamivir has little or no effect on admission to hospital. |
|  |  | Difference: 1 fewer per 1000<br>(CI 95% 1 fewer - 1 more) |  |  |  |
| Admission to hospital<br>(High-risk) | Relative risk: 0.80<br>(CI 95% 0.54 - 1.18)<br>Based on data from 12589 participants in 20 studies | 21<br>per 1000 | 17<br>per 1000 | High | Oseltamivir has little or no effect on admission to hospital. |
|  |  | Difference: 4 fewer per 1000<br>(CI 95% 10 fewer - 4 more) |  |  |  |
| Adverse events related to treatments | Relative risk: 1.23<br>(CI 95% 1.1 - 1.39)<br>Based on data from 6782 participants in 12 studies | 122<br>per 1000 | 150<br>per 1000 | Moderate<br>Due to serious risk of bias | Oseltamivir probably increases adverse events related to treatments. |
|  |  | Difference: 28 more per 1000<br>(CI 95% 12 more - 48 more) |  |  |  |
| Serious adverse events | Risk difference: 0.0<br>(CI 95% -0.003 - 0.002)<br>Based on data from 14718 participants in 22 studies | 5<br>per 1000 | 5<br>per 1000 | Moderate<br>Due to serious risk of bias | Oseltamivir probably has little or no effect on serious adverse events. |
|  |  | Difference: 0 fewer per 1000<br>(CI 95% 3 fewer - 2 more) |  |  |  |
| Time to alleviation of symptoms | Measured by: day<br>Lower better<br>Based on data from 9078 participants in 22 studies | 4.92<br>Mean | 4.17<br>Mean | Moderate<br>Due to serious risk of bias | Oseltamivir probably has no important effect on time to alleviation of symptoms. |
|  |  | Difference: MD 0.75 lower<br>(CI 95% 0.93 lower - 0.57 lower) |  |  |  |

Forty-one trials including 23892 patients reported mortality. High certainty evidence showed that, compared with standard care or placebo, seven antivirals (i.e. baloxavir, favipiravir, laninamivir, oseltamivir, peramivir, umifenovir, zanamivir) have little or no effect on mortality in low-risk (RD varied from 0.12 fewer to 0.02 fewer per 1000) or high-risk (RD varied from 1.22 fewer to 0.24 fewer per 1000) patients (Figure 3, Table 1, Table 2, and Appendix 11).

**Figure 3.**
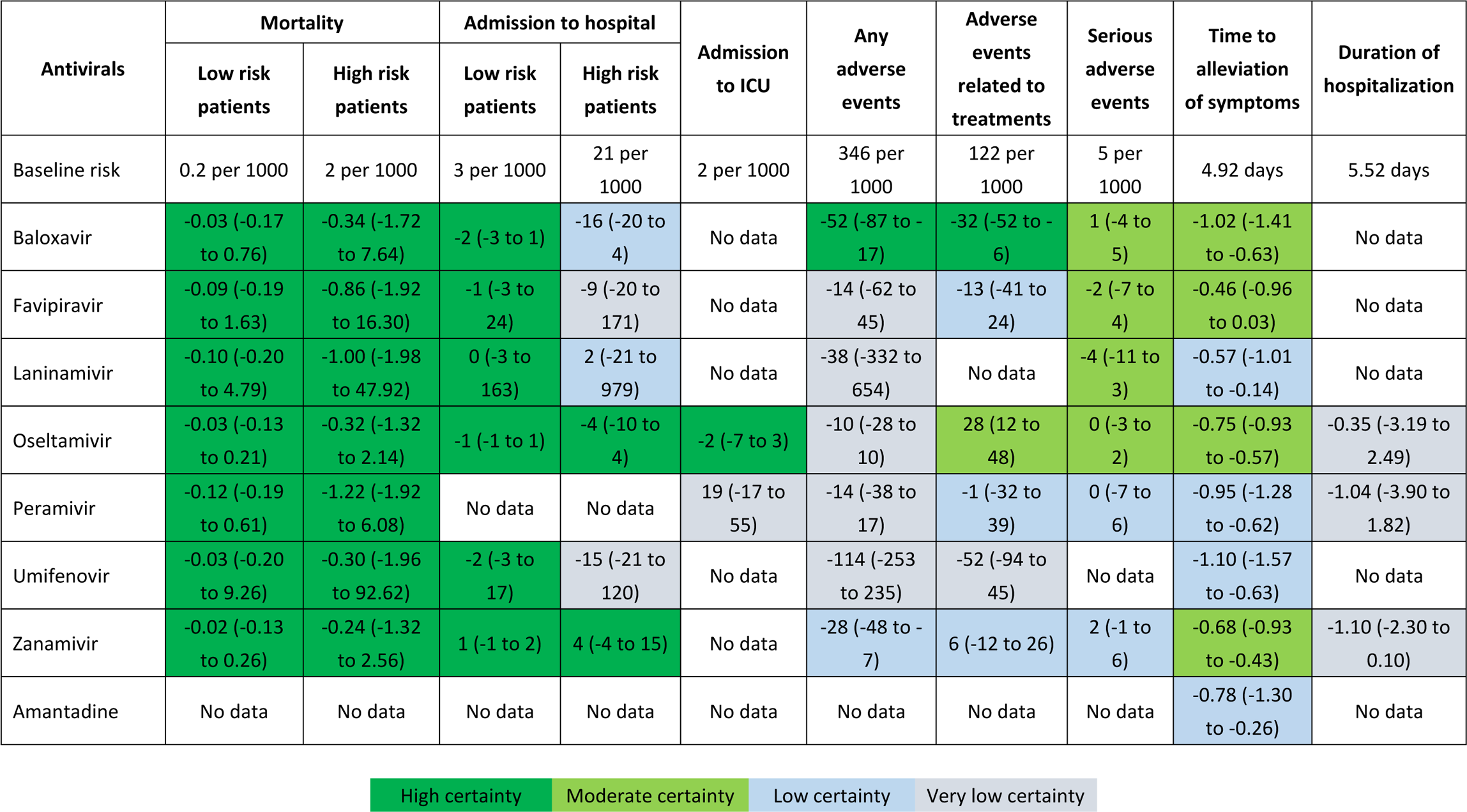
Summary of effects of antivirals versus standard care or placebo, presented as absolute effects with 95% confidence intervals (CIs) Numbers in the colored cell are the estimated risk differences (95% CI) per 1000 patients or mean difference (95% CI) in days when compared with standard care or placebo.

Twenty-eight trials involving 17262 patients reported admission to hospital for all antivirals except for peramivir and amantadine (no data available). For low-risk patients, there proved little or no difference between six antivirals (baloxavir, favipiravir, laninamivir, oseltamivir, umifenovir, and zanamivir) and standard care or placebo in admission to hospital (RD varied from 2 fewer to 1 more per 1000; high certainty). For admission to hospital in high-risk patients, compared with standard care or placebo, baloxavir may reduce risk (RD 16 fewer per 1000, 95% CI 20 fewer to 4 more; low certainty); oseltamivir (RD 4 fewer per 1000, 95% CI 10 fewer to 4 more; high certainty) and zanamivir (RD 4 more per 1000, 95% CI 4 fewer to 15 more; high certainty) have little or no effect; and laninamivir may have little or no effect (RD 2 more per 1000, 95% CI 21 fewer to 979 more; low certainty) (Figure 3, Table 1, Table 2, and Appendix 11).

Two trials with 1626 patients provided data on ICU admission for oseltamivir and peramivir. Compared with standard care or placebo, oseltamivir has little or no effect on admission to ICU (RD 2 fewer per 1000, 95% CI 7 fewer to 3 more; high certainty). We are uncertain whether peramivir reduces admission to the ICU (Figure 3). No data is available regarding the effect of other antivirals on ICU admission.

Fifty-nine trials enrolling 24086 patients reported time to alleviation of symptoms. Compared with standard care or placebo, baloxavir probably reduces symptom duration (MD 1.02 days lower, 95% CI 1.41 lower to 0.63 lower; moderate certainty); umifenovir may reduce symptom duration (MD 1.10 days lower, 95% CI 1.57 lower to 0.63 lower; low certainty); while oseltamivir (MD 0.75 days lower, 95% CI 0.93 lower to 0.57 lower, moderate certainty) and zanamivir (MD 0.68 days lower, 95% CI 0.93 lower to 0.43 lower, moderate certainty) probably have no important effect (Figure 3, Table 1, Table 2, and Appendix 11).

Three trials with 234 patients provided data on duration of hospitalization for oseltamivir, peramivir, and zanamivir. The evidence is very uncertain as to whether these drugs reduce duration of hospitalization (Figure 3). No data is available regarding the effect of other antivirals on duration of hospitalization.

Forty-nine trials with 22868 patients reported data on overall adverse events. No data is available for amantadine. Compared with standard care or placebo, baloxavir (RD 52 fewer per 1000, 95% CI 87 fewer to 17 fewer; high certainty) does not increase overall adverse events and zanamivir (RD 28 fewer per 1000, 95% CI 48 fewer to 7 fewer; low certainty) may not result in an increase. It is uncertain whether favipiravir, laninamivir, oseltamivir, peramivir, and umifenovir increase adverse events (Figure 3).

Thirty-six trials with 19298 patients reported data on adverse events related to treatments. Compared with standard care or placebo, baloxavir (RD 32 fewer per 1000, 95% CI 52 fewer to 6 fewer; high certainty) does not increase adverse events related to treatment and oseltamivir (RD 28 more per 1000, 95% CI 12 more to 48 more; moderate certainty) probably increases adverse events related to treatment. Favipiravir, peramivir, and zanamivir may not increase adverse events related to treatment. We are uncertain whether umifenovir increases adverse events related to treatment (Figure 3).

Fifty-eight trials involving 32043 patients provided low to moderate evidence that there may be little or no difference in serious adverse events between baloxavir, favipiravir, laninamivir, oseltamivir, peramivir, or zanamivir and standard care or placebo. No data is available for umifenovir and amantadine on serious adverse events. (Figure 3, Table 1, Table 2, and Appendix 11).

Twelve trials enrolling 1554 patients reported the emergence of resistance for participants who received antivirals. Baloxavir may have an important impact on emergence of drug resistance (percentage 9.97%, 95% CI 0.02% to 31.79%; low certainty). Zanamivir may have no important impact on emergence of drug resistance (percentage 0%, 95% CI 0% to 11.66%; low certainty). We are uncertain whether oseltamivir and peramivir increase emergence of drug resistance (very low certainty; Appendix 13). There was no evidence regarding drug resistance for other antivirals from included RCTs.

The within-trial subgroup analyses and meta-regression analyses did not reveal any subgroup effects (Appendix 14 and Appendix 15). Our sensitivity analyses using the RD as summary effect measure for mortality and hospital admission (Appendix 16), only including studies with confirmed influenza patients (Appendix 17), and only including studies with patients at high risk (Appendix 18) showed similar results to primary analyses of all patients.

## Discussion

In patients with non-severe influenza, we found high certainty evidence that compared with standard care or placebo, baloxavir, favipiravir, laninamivir, oseltamivir, peramivir, umifenovir, and zanamivir have little or no effect on mortality for low-risk and high-risk patients, high certainty evidence that oseltamivir and zanamivir have little or no effect on admission to hospital for high-risk patients, and low certainty evidence that baloxavir may reduce the risk of admission to hospital for high-risk patients. All antivirals have little or no effect on admission to hospital for low-risk patients. With respect to time to alleviation of symptoms, baloxavir probably reduces symptom duration; oseltamivir and zanamivir probably have no important effect; laninamivir, peramivir, and amantadine may have no important effect; and umifenovir may reduce symptom duration. Baloxavir does not increase adverse events related to treatment but may result in resistance in approximately 10% of those treated. Oseltamivir probably increases adverse events related to treatment. Other antivirals probably or may have little or no effect on adverse events.

The strengths of the review include a comprehensive synthesis of all available evidence on the benefits and harms of antivirals for non-severe influenza patients; independent duplicate study identification, selection, data extraction, and risk of bias assessment; and application of the GRADE approach to rate the certainty of evidence. To facilitate interpretation of the results, we present absolute effects. Based on the WHO guideline development panel’s discussion and suggestions, we evaluated the effects of antivirals on mortality and hospital admission for both low-risk and high-risk populations.

The WHO guideline panel provided us with the MID values for patient-important outcomes and reviewed the results of our review. We used these MID values to interpret the results, assist our imprecision rating, and draw conclusions.

During the rating of imprecision, we followed GRADE guidance and considered external indirect evidence.^38,39^ For certain outcomes, such as mortality in patients at low and high risk, and admission to hospital in low-risk patients, where the baseline risk was lower than the MID, the absolute effects between comparisons did not exceed the MID, and no intervention was superior to the other despite the certainty of evidence. Therefore, we rated the certainty of evidence for all comparisons as high certainty.

Our review has limitations. The evidence for some antiviral drugs and some outcomes was limited. The panel was also interested in progression to mechanical ventilation, duration of mechanical ventilation, and hospital discharge destination, but no trials reported these outcomes. Because within-trial subgroup information was limited, we were only able to conduct within-trial subgroup analysis for certain outcomes and comparisons and did not find any significant subgroup effects. We performed meta-regression analyses for the effect modifiers and also did not find significant subgroup effects.

For mortality and hospital admission, to estimate separately absolute effects for patients at low risk and high risk using network relative estimates and the baseline risks, we calculated risk ratios and applied the continuity correction of 0.5 for trials with 0 events.^28^ Reassuringly, our findings were robust to sensitivity analyses using RDs as the summary measure.

Due to the high risk of bias or serious imprecision, the certainty of evidence was low for many comparisons. As future trials emerge, we anticipate an improvement in the certainty of evidence and will periodically update this systematic review and network meta-analysis.

Several network meta-analyses have evaluated antivirals for treating influenza ^8–10,42,43^, but none assessed all approved antivirals for influenza and none focused on patient-important outcomes of mortality and admission to hospital. Compared with previous network meta-analyses ^8–10,42,43^, we included more trials and patients, assessed the certainty of evidence using the GRADE approach, and presented absolute effects. Our findings on baloxavir are consistent with previous network meta-analyses that baloxavir probably reduces time to alleviation of symptoms without increasing adverse events.^8,10^

Previous pairwise meta-analyses, including no more than nine studies, have evaluated the effects of oseltamivir on influenza and reported that, compared to standard care or placebo, oseltamivir might reduce hospitalization.^18,44^ The results differed from our findings mainly because the previous meta-analyses only included studies published before 2014. Another more recent meta-analysis including 12 studies with 6295 outpatients patients reported that oseltamivir may not be associated with reduced risk of hospitalization and may associated with increased nausea and vomiting.^45^ Our findings on the effects of oseltamivir for non-severe influenza are consistent with results of this meta-analysis.^45^ By increasing sample size, our review has added statistical power and improved the precision of estimates and allowed the conclusion that oseltamivir does not result in an important reduction in symptom duration.

Our findings have significant implications for practice and research. Oseltamivir has been widely used for treatment of influenza. However, this review provided moderate to high certainty evidence that, compared to standard care or placebo, oseltamivir has little or no effect on mortality and admission to hospital both in low-risk and high-risk patients, probably has no important effect on time to alleviation of symptoms, and probably increases adverse events related to treatment. Therefore, our findings did not support the use of oseltamivir for treatment of non-severe influenza.

Baloxavir, relative to standard care, may reduce the risk of admission to hospital in high-risk patients, probably reduces time to alleviation of symptoms, and does not increase adverse events related to treatment. These findings support the use of baloxavir for treatment of high-risk non-severe influenza patients. However, drug resistance monitoring may be needed as baloxavir may increase emergence of drug resistance.

Our studies identify evidence gaps regarding antivirals for non-severe influenza. Future studies could focus on patient-important outcomes (e.g. admission to ICU, progression to mechanical ventilation, duration of hospitalization) and test important subgroup analyses such as the influenza virus type, age, patients at low risk or high risk, and time from onset of symptoms to treatment. Future antiviral trials of influenza should also pay greater attention to ensuring adequate allocation concealment and blinding.

In patients with non-severe influenza, baloxavir may reduce the risk of admission to hospital for high-risk patients, probably reduces time to alleviation of symptoms, does not increase any adverse events and adverse events related to treatment, but may result in resistance in approximately 10% of those treated. Oseltamivir has little or no effect on mortality and admission to hospital, probably has no important effect on time to alleviation of symptoms, and probably increases adverse events related to treatment. All other antivirals are either probably not to be beneficial or have uncertain effects on patient-important outcomes.

## Supporting information

PRISMA

## Data Availability

All data produced in the present work are contained in the manuscript.

## Contributors

YG, GG, and QH conceived and designed the study. YG and QH designed and performed the search strategy. YG, YZ, ML, SL, and XC screened and selected the articles. YG, YZ, ML, SL, YC, XC, QZ, JX, YS, WZ, ZL, SH, and JH extracted the data and assessed the risk of bias. YG and QH analyzed the data. JT and GG supervised the data analyses. YG and QH rated the certainty of evidence. GG provided methodological advice on baseline risk selection and GRADE assessment. YG, YZ, GG, and QH interpreted the data. YG and QH drafted the manuscript. YG, GG, and QH revised the manuscript. All authors approved the final version of the manuscript. YG and QH accessed and verified the underlying data. All authors had full access to all the data in the study and had final responsibility for the decision to submit for publication.

## Data sharing statement

Data in this systematic review with meta-analysis are extracted from published studies available on the internet. All processed data are presented in this article and the appendix.

## Declaration of interests

We declare no competing interests.

## Acknowledgements

We thank Timothy M Uyeki (Influenza Division, US Centers for Disease Control and Prevention, Atlanta, Georgia) for help with identifying the drug resistance outcome, providing clinical advice, and interpreting the results. We thank members of the WHO for critical feedback on the review question, subgroup and outcome selection, and GRADE judgments: Janet Diaz (World Health Organization, Geneva, Switzerland); Steven Mcgloughlin (World Health Organization, Geneva, Switzerland); Jamie Rylance (World Health Organization, Geneva, Switzerland). We thank Rachel Couban (librarian at McMaster University) for helping with developing the search strategy. We thank Ping Liu (The First Affiliated Hospital of Chongqing Medical University, Chongqing, China) for contributing to the literature screening.

## Notes

### Competing Interest Statement

The authors have declared no competing interest.

### Clinical Protocols

https://www.crd.york.ac.uk/prospero/display_record.php?RecordID=456650

### Funding Statement

This study was funded by WHO.

## References

1. Paules C, Subbarao K. Influenza. Lancet 2017; 390(10095): 697–708.

2. Uyeki TM, Hui DS, Zambon M, Wentworth DE, Monto AS. Influenza. Lancet 2022; 400(10353): 693–706.

3. World Health Organization. WHO Influenza (seasonal) fact sheet. Geneva: World Health Organization; 2023. https://www.who.int/news-room/fact-sheets/detail/influenza-(seasonal). Accessed 6 February 2023.

4. Iuliano AD, Roguski KM, Chang HH, et al. Estimates of global seasonal influenza-associated respiratory mortality: a modelling study. Lancet 2018; 391(10127): 1285–300.

5. Kumari R, Sharma SD, Kumar A, et al. Antiviral Approaches against Influenza Virus. Clin Microbiol Rev 2023: e0004022.

6. Hsu J, Santesso N, Mustafa R, et al. Antivirals for treatment of influenza: a systematic review and meta-analysis of observational studies. Ann Intern Med 2012; 156(7): 512–24.

7. World Health Organization. Guidelines for the clinical management of severe illness from influenza virus infections. Geneva: World Health Organization; 2021. https://www.who.int/publications/i/item/9789240040816. Accessed 6 February 2023.

8. Liu JW, Lin SH, Wang LC, Chiu HY, Lee JA. Comparison of Antiviral Agents for Seasonal Influenza Outcomes in Healthy Adults and Children: A Systematic Review and Network Meta-analysis. JAMA Netw Open 2021; 4(8): e2119151.

9. Su HC, Feng IJ, Tang HJ, Shih MF, Hua YM. Comparative effectiveness of neuraminidase inhibitors in patients with influenza: A systematic review and network meta-analysis. J Infect Chemother 2022; 28(2): 158–69.

10. Zhao Y, Huang G, He W, et al. Efficacy and safety of single-dose antiviral drugs for influenza treatment: A systematic review and network meta-analysis. J Med Virol 2022; 94(7): 3270–302.

11. Butler CC, van der Velden AW, Bongard E, et al. Oseltamivir plus usual care versus usual care for influenza-like illness in primary care: an open-label, pragmatic, randomised controlled trial. Lancet (London, England) 2020; 395(10217): 42–52.

12. Hayden FG, Lenk RP, Stonis L, Oldham-Creamer C, Kang LL, Epstein C. Favipiravir Treatment of Uncomplicated Influenza in Adults: Results of Two Phase 3, Randomized, Double-Blind, Placebo-Controlled Trials. The Journal of infectious diseases 2022; 226(10): 1790–9.

13. Bai X, Xi S, Chen G, et al. Multicenter, randomized controlled, open label evaluation of the efficacy and safety of arbidol hydrochloride tablets in the treatment of influenza-like cases. BMC infectious diseases 2023; 23(1): 585.

14. Hsieh Y-H, Dugas AF, LoVecchio F, et al. Intravenous peramivir vs oral oseltamivir in high-risk emergency department patients with influenza: Results from a pilot randomized controlled study. Influenza and other respiratory viruses 2021; 15(1): 121–31.

15. Hutton B, Salanti G, Caldwell DM, et al. The PRISMA extension statement for reporting of systematic reviews incorporating network meta-analyses of health care interventions: checklist and explanations. Ann Intern Med 2015; 162(11): 777–84.

16. Brown SJ, Bakker CJ, Theis-Mahon NR. Retracted publications in pharmacy systematic reviews. J Med Libr Assoc 2022; 110(1): 47–55.

17. Ison MG, Portsmouth S, Yoshida Y, et al. Early treatment with baloxavir marboxil in high-risk adolescent and adult outpatients with uncomplicated influenza (CAPSTONE-2): a randomised, placebo-controlled, phase 3 trial. The Lancet Infectious Diseases 2020; 20(10): 1204–14.

18. Dobson J, Whitley RJ, Pocock S, Monto AS. Oseltamivir treatment for influenza in adults: a meta-analysis of randomised controlled trials. Lancet 2015; 385(9979): 1729–37.

19. Harbord RM, Egger M, Sterne JA. A modified test for small-study effects in meta-analyses of controlled trials with binary endpoints. Stat Med 2006; 25(20): 3443–57.

20. Egger M, Davey Smith G, Schneider M, Minder C. Bias in meta-analysis detected by a simple, graphical test. BMJ 1997; 315(7109): 629–34.

21. Nyaga VN, Arbyn M, Aerts M. Metaprop: a Stata command to perform meta-analysis of binomial data. Arch Public Health 2014; 72(1): 39.

22. IntHout J, Ioannidis JP, Borm GF. The Hartung-Knapp-Sidik-Jonkman method for random effects meta-analysis is straightforward and considerably outperforms the standard DerSimonian-Laird method. BMC Med Res Methodol 2014; 14: 25.

23. Barker TH, Migliavaca CB, Stein C, et al. Conducting proportional meta-analysis in different types of systematic reviews: a guide for synthesisers of evidence. BMC Med Res Methodol 2021; 21(1): 189.

24. Rucker G. Network meta-analysis, electrical networks and graph theory. Res Synth Methods 2012; 3(4): 312–24.

25. Higgins JP, Jackson D, Barrett JK, Lu G, Ades AE, White IR. Consistency and inconsistency in network meta-analysis: concepts and models for multi-arm studies. Res Synth Methods 2012; 3(2): 98–110.

26. Konig J, Krahn U, Binder H. Visualizing the flow of evidence in network meta-analysis and characterizing mixed treatment comparisons. Stat Med 2013; 32(30): 5414–29.

27. Dias S, Welton NJ, Caldwell DM, Ades AE. Checking consistency in mixed treatment comparison meta-analysis. Stat Med 2010; 29(7-8): 932–44.

28. Friedrich JO, Adhikari NKJ, Beyene J. Inclusion of zero total event trials in meta-analyses maintains analytic consistency and incorporates all available data. Bmc Medical Research Methodology 2007; 7: 5.

29. McDonald SA, Teirlinck AC, Hooiveld M, et al. Inference of age-dependent case-fatality ratios for seasonal influenza virus subtypes A(H3N2) and A(H1N1)pdm09 and B lineages using data from the Netherlands. Influenza and Other Respiratory Viruses 2023; 17(6): e13146.

30. Meier CR, Napalkov PN, Wegmüller Y, Jefferson T, Jick H. Population-based study on incidence, risk factors, clinical complications and drug utilisation associated with influenza in the United Kingdom. Eur J Clin Microbiol 2000; 19(11): 834–42.

31. Higgins JPT, Thomas J, Chandler J, Cumpston M, Li T, Page MJ, Welch VA, editors. Cochrane Handbook for Systematic Reviews of Interventions version 6.3 (updated February 2022). Cochrane Collaboration; 2022. www.training.cochrane.org/handbook.

32. Schandelmaier S, Briel M, Varadhan R, et al. Development of the Instrument to assess the Credibility of Effect Modification Analyses (ICEMAN) in randomized controlled trials and meta-analyses. CMAJ 2020; 192(32): E901–e6.

33. Guyatt GH, Busse JW. Modification of cochrane tool to assess risk of bias in randomized trials. https://www.evidencepartners.com/resources/methodological-resources/. Accessed January 30, 2023.

34. Puhan MA, Schünemann HJ, Murad MH, et al. A GRADE Working Group approach for rating the quality of treatment effect estimates from network meta-analysis. BMJ 2014; 349: g5630.

35. Brignardello-Petersen R, Bonner A, Alexander PE, et al. Advances in the GRADE approach to rate the certainty in estimates from a network meta-analysis. J Clin Epidemiol 2018; 93: 36–44.

36. Guyatt GH, Oxman AD, Vist GE, et al. GRADE: an emerging consensus on rating quality of evidence and strength of recommendations. BMJ 2008; 336(7650): 924–6.

37. Zeng L, Brignardello-Petersen R, Hultcrantz M, et al. GRADE guidelines 32: GRADE offers guidance on choosing targets of GRADE certainty of evidence ratings. J Clin Epidemiol 2021; 137: 163–75.

38. Zeng L, Brignardello-Petersen R, Hultcrantz M, et al. GRADE Guidance 34: update on rating imprecision using a minimally contextualized approach. J Clin Epidemiol 2022; 150: 216–24.

39. Hao Q, Gao Y, Zhao Y, et al. GRADE concept 6: a novel application of external indirect evidence into GRADE ratings of evidence certainty in network meta-analysis. J Clin Epidemiol 2023; 163: 95–101.

40. Carrasco-Labra A, Brignardello-Petersen R, Santesso N, et al. Improving GRADE evidence tables part 1: a randomized trial shows improved understanding of content in summary of findings tables with a new format. J Clin Epidemiol 2016; 74: 7–18.

41. Santesso N, Glenton C, Dahm P, et al. GRADE guidelines 26: informative statements to communicate the findings of systematic reviews of interventions. J Clin Epidemiol 2020; 119: 126–35.

42. Taieb V, Ikeoka H, Ma FF, et al. A network meta-analysis of the efficacy and safety of baloxavir marboxil versus neuraminidase inhibitors for the treatment of influenza in otherwise healthy patients. Curr Med Res Opin 2019; 35(8): 1355–64.

43. Taieb V, Ikeoka H, Wojciechowski P, et al. Efficacy and safety of baloxavir marboxil versus neuraminidase inhibitors in the treatment of influenza virus infection in high-risk and uncomplicated patients - a Bayesian network meta-analysis. Curr Med Res Opin 2021; 37(2): 225–44.

44. Shim SJ, Chan M, Owens L, Jaffe A, Prentice B, Homaira N. Rate of use and effectiveness of oseltamivir in the treatment of influenza illness in high-risk populations: A systematic review and meta-analysis. Health Sci Rep 2021; 4(1): e241.

45. Hanula R, Bortolussi-Courval E, Mendel A, Ward BJ, Lee TC, McDonald EG. Evaluation of Oseltamivir Used to Prevent Hospitalization in Outpatients With Influenza: A Systematic Review and Meta-Analysis. JAMA Intern Med 2024; 184(1): 18–27.

