## Appendix for "Antivirals for treatment of non-severe influenza: a systematic review and network meta-analysis of randomized controlled trials"

### Appendix 1. Search strategy of databases

**Ovid MEDLINE(R) ALL**

1	exp Influenza, Human/

2	exp Influenza A virus/

3	exp Influenza B virus/

4	exp Influenzavirus C/

5	(Influenza or flu or H1N1 or PH1N1 or H3N2 or AH1N1 or AH3N2 or H5N1 or H7N9).mp. [mp=title, book title, abstract, original title, name of substance word, subject heading word, floating sub-heading word, keyword heading word, organism supplementary concept word, protocol supplementary concept word, rare disease supplementary concept word, unique identifier, synonyms, population supplementary concept word, anatomy supplementary concept word]

6	or/1-5

7	Antiviral agents/

8	Antiviral*.tw.

9	(neuraminidase inhibitor* or NA inhibitor*).tw.

10	Oseltamivir/ or Zanamivir/

11	(oseltamivir or tamiflu or "GS 4104" or GS4104 or GS-4104 or "GS 4071" or GS4071 or GS-4071 or zanamivir or relenza or "GG 167" or GG167 or GG-167 or CS-8958 or Dectova or Laninamivir or R-125489 or R125489 or "R 125489" or Inavir or peramivir or "BCX 1812" or BCX1812 or BCX-1812 or "RWJ 270201" or RWJ270201 or RWJ-270201 or Rapivab or rapiacta).ti,ab.

12	Viral Polymerase Complex Inhibitor*.tw.

13	(Favipiravir or T-705 or Avigan or FabiFlu or Pimodivir or VX-787 or JNJ-63623872 or AL-794 or ALS-033719 or ZSP1273 or Enisamium iodide or FAV00A or TG-1000 or GP681).ti,ab.

14	matrix protein 2 ion channel inhibitor*.tw.

15	(Radavirsen or AVI-7100).ti,ab.

16	cap-dependent endonuclease inhibitor*.tw.

17	("Baloxavir marboxil" or Baloxavir or S-033188 or Xofluza).ti,ab.

18	(Umifenovir or Arbidol or Arbidole).ti,ab.

19	Amantadine/ or Rimantadine/

20	(Amantadine or Symmetrel or Symetrel or Rimantadine or Flumadine or Roflual).ti,ab.

21	or/7-20

22	6 and 21

23	randomized controlled trial.pt.

24	controlled clinical trial.pt.

25	randomized.ab.

26	placebo.ab.

27	drug therapy.fs.

28	randomly.ab.

29	trial.ti.

30	groups.ab.

31	or/23-30

32	(animals not (humans and animals)).sh.

33	31 not 32

34	22 and 33

**Ovid Embase**

1	exp Influenza/ or Influenza virus/

2	exp Influenza A virus/ or exp Influenza A virus/

3	exp Influenza B/ or exp Influenza B virus/

4	exp Influenza C/ or exp Influenza C virus/

5	(Influenza or flu or H1N1 or PH1N1 or H3N2 or AH1N1 or AH3N2 or H5N1 or H7N9).mp. [mp=title, abstract, heading word, drug trade name, original title, device manufacturer, drug manufacturer, device trade name, keyword heading word, floating subheading word, candidate term word]

6	or/1-5

7	Antivirus agent/

8	Antiviral*.tw.

9	(neuraminidase inhibitor* or NA inhibitor*).tw.

10	Sialidase inhibitor/ or Oseltamivir/ or Zanamivir/ or Laninamivir/ or Peramivir/

11	(oseltamivir or tamiflu or "GS 4104" or GS4104 or GS-4104 or "GS 4071" or GS4071 or GS-4071 or zanamivir or relenza or "GG 167" or GG167 or GG-167 or CS-8958 or Dectova or Laninamivir or R-125489 or R125489 or "R 125489" or Inavir or peramivir or "BCX 1812" or BCX1812 or BCX-1812 or "RWJ 270201" or RWJ270201 or RWJ-270201 or Rapivab or rapiacta).ti,ab.

12	Viral Polymerase Complex Inhibitor*.tw.

13	Favipiravir/ or Pimodivir/ or (Favipiravir or T-705 or Avigan or FabiFlu or Pimodivir or VX-787 or JNJ-63623872 or AL-794 or ALS-033719 or ZSP1273 or Enisamium iodide or FAV00A or TG-1000 or GP681).ti,ab.

14	matrix protein 2 ion channel inhibitor*.tw.

15	Radavirsen/ or (Radavirsen or AVI-7100).ti,ab.

16	cap-dependent endonuclease inhibitor*.tw.

17	Baloxavir marboxil/ or ("Baloxavir marboxil" or Baloxavir or S-033188 or Xofluza).ti,ab.

18	Umifenovir/ or (Umifenovir or Arbidol or Arbidole).ti,ab.

19	Amantadine/ or Rimantadine/

20	(Amantadine or Symmetrel or Symetrel or Rimantadine or Flumadine or Roflual).ti,ab.

21	or/7-20

22	6 and 21

23	Randomized controlled trial/

24	Controlled clinical study/

25	random$.ti,ab.

26	randomization/

27	intermethod comparison/

28	placebo.ti,ab.

29	(compare or compared or comparison).ti.

30	((evaluated or evaluate or evaluating or assessed or assess) and (compare or compared or comparing or comparison)).ab.

31	(open adj label).ti,ab.

32	((double or single or doubly or singly) adj (blind or blinded or blindly)).ti,ab.

33	double blind procedure/

34	parallel group$1.ti,ab.

35	(crossover or cross over).ti,ab.

36	((assign$ or match or matched or allocation) adj5 (alternate or group$1 or intervention$1 or patient$1 or subject$1 or participant$1)).ti,ab.

37	(assigned or allocated).ti,ab.

38	(controlled adj7 (study or design or trial)).ti,ab.

39	(volunteer or volunteers).ti,ab.

40	human experiment/

41	trial.ti.

42	or/23-41

43	(random$ adj sampl$ adj7 ("cross section$" or questionnaire$1 or survey$ or database$1)).ti,ab. not (comparative study/ or controlled study/ or randomi?ed controlled.ti,ab. or randomly assigned.ti,ab.)

44	Cross-sectional study/ not (randomized controlled trial/ or controlled clinical study/ or controlled study/ or randomi?ed controlled.ti,ab. or control group$1.ti,ab.)

45	(((case adj control$) and random$) not randomi?ed controlled).ti,ab.

46	(Systematic review not (trial or study)).ti.

47	(nonrandom$ not random$).ti,ab.

48	"Random field$".ti,ab.

49	(random cluster adj3 sampl$).ti,ab.

50	(review.ab. and review.pt.) not trial.ti.

51	"we searched".ab. and (review.ti. or review.pt.)

52	"update review".ab.

53	(databases adj4 searched).ab.

54	(rat or rats or mouse or mice or swine or porcine or murine or sheep or lambs or pigs or piglets or rabbit or rabbits or cat or cats or dog or dogs or cattle or bovine or monkey or monkeys or trout or marmoset$1).ti. and animal experiment/

55	Animal experiment/ not (human experiment/ or human/)

56	or/43-55

57	42 not 56

58	22 and 57

**Cochrane Central Register of Controlled Trials**

1	exp Influenza, Human/

2	exp Influenza A virus/

3	exp Influenza B virus/

4	exp Influenzavirus C/

5	(Influenza or flu or H1N1 or PH1N1 or H3N2 or AH1N1 or AH3N2 or H5N1 or H7N9).mp. [mp=title, original title, abstract, floating sub-heading word, mesh headings, heading words, keyword]

6	or/1-5

7	Antiviral agents/

8	Antiviral*.tw.

9	(neuraminidase inhibitor* or NA inhibitor*).tw.

10	Oseltamivir/ or Zanamivir/

11	(oseltamivir or tamiflu or "GS 4104" or GS4104 or GS-4104 or "GS 4071" or GS4071 or GS-4071 or zanamivir or relenza or "GG 167" or GG167 or GG-167 or CS-8958 or Dectova or Laninamivir or R-125489 or R125489 or "R 125489" or Inavir or peramivir or "BCX 1812" or BCX1812 or BCX-1812 or "RWJ 270201" or RWJ270201 or RWJ-270201 or Rapivab or rapiacta).ti,ab.

12	Viral Polymerase Complex Inhibitor*.tw.

13	(Favipiravir or T-705 or Avigan or FabiFlu or Pimodivir or VX-787 or JNJ-63623872 or AL-794 or ALS-033719 or ZSP1273 or Enisamium iodide or FAV00A or TG-1000 or GP681).ti,ab.

14	matrix protein 2 ion channel inhibitor*.tw.

15	(Radavirsen or AVI-7100).ti,ab.

16	cap-dependent endonuclease inhibitor*.tw.

17	("Baloxavir marboxil" or Baloxavir or S-033188 or Xofluza).ti,ab.

18	(Umifenovir or Arbidol or Arbidole).ti,ab.

19	Amantadine/ or Rimantadine/

20	(Amantadine or Symmetrel or Symetrel or Rimantadine or Flumadine or Roflual).ti,ab.

21	or/7-20

22	6 and 21

23	randomized controlled trial.pt.

24	controlled clinical trial.pt.

25	randomized.ab.

26	placebo.ab.

27	drug therapy.fs.

28	randomly.ab.

29	trial.ti.

30	groups.ab.

31	or/23-30

32	(animals not (humans and animals)).sh.

33	31 not 32

34	22 and 33

**Global Health**

1	exp Influenza/ or Influenza viruses/

2	exp Influenza A virus/ or exp Influenza A virus/

3	exp Influenza B/ or exp Influenza B virus/

4	exp Influenza C/ or exp Influenza C virus/

5	(Influenza or flu or H1N1 or PH1N1 or H3N2 or AH1N1 or AH3N2 or H5N1 or H7N9).mp. [mp=abstract, title, original title, heading words, cabicodes words]

6	or/1-5

7	Antiviral agents/

8	Antiviral*.tw.

9	(neuraminidase inhibitor* or NA inhibitor*).tw.

10	Sialidase inhibitors/ or Oseltamivir/ or Zanamivir/ or Laninamivir/ or Peramivir/

11	(oseltamivir or tamiflu or "GS 4104" or GS4104 or GS-4104 or "GS 4071" or GS4071 or GS-4071 or zanamivir or relenza or "GG 167" or GG167 or GG-167 or CS-8958 or Dectova or Laninamivir or R-125489 or R125489 or "R 125489" or Inavir or peramivir or "BCX 1812" or BCX1812 or BCX-1812 or "RWJ 270201" or RWJ270201 or RWJ-270201 or Rapivab or rapiacta).ti,ab.

12	Viral Polymerase Complex Inhibitor*.tw.

13	Favipiravir/ or (Favipiravir or T-705 or Avigan or FabiFlu or Pimodivir or VX-787 or JNJ-63623872 or AL-794 or ALS-033719 or ZSP1273 or Enisamium iodide or FAV00A or TG-1000 or GP681).ti,ab.

14	matrix protein 2 ion channel inhibitor*.tw.

15	(Radavirsen or AVI-7100).ti,ab.

16	cap-dependent endonuclease inhibitor*.tw.

17	("Baloxavir marboxil" or Baloxavir or S-033188 or Xofluza).ti,ab.

18	(Umifenovir or Arbidol or Arbidole).ti,ab.

19	Amantadine/ or Rimantadine/

20	(Amantadine or Symmetrel or Symetrel or Rimantadine or Flumadine or Roflual).ti,ab.

21	or/7-20

22	6 and 21

23	exp randomized controlled trials/

24	(randomized controlled trial or random* or blind* or placebo*).mp. [mp=abstract, title, original title, heading words, cabicodes words]

25	23 or 24

26	22 and 25

**CINAHL**

| **#** | **Query** |
| --- | --- |
| S36 | S23 AND S26 AND S35 |
| S35 | S27 OR S28 OR S29 OR S30 OR S31 OR S32 OR S33 OR S34 |
| S34 | TI ( matrix protein 2 ion channel inhibitor* OR Radavirsen or AVI-7100 OR cap-dependent endonuclease inhibitor* OR "Baloxavir marboxil" or Baloxavir or S-033188 or Xofluza OR Umifenovir or Arbidol or Arbidole OR Amantadine or Symmetrel or Symetrel or Rimantadine or Flumadine or Roflual ) OR AB ( matrix protein 2 ion channel inhibitor* OR Radavirsen or AVI-7100 OR cap-dependent endonuclease inhibitor* OR "Baloxavir marboxil" or Baloxavir or S-033188 or Xofluza OR Umifenovir or Arbidol or Arbidole OR Amantadine or Symmetrel or Symetrel or Rimantadine or Flumadine or Roflual ) |
| S33 | (MH "Amantadine") |
| S32 | TI ( Viral Polymerase Complex Inhibitor* OR Favipiravir or T-705 or Avigan or FabiFlu or Pimodivir or VX-787 or JNJ-63623872 or AL-794 or ALS-033719 or ZSP1273 or Enisamium iodide or FAV00A or TG-1000 or GP681 ) OR AB ( Viral Polymerase Complex Inhibitor* OR Favipiravir or T-705 or Avigan or FabiFlu or Pimodivir or VX-787 or JNJ-63623872 or AL-794 or ALS-033719 or ZSP1273 or Enisamium iodide or FAV00A or TG-1000 or GP681 ) |
| S31 | TI ( oseltamivir or tamiflu or "GS 4104" or GS4104 or GS-4104 or "GS 4071" or GS4071 or GS-4071 or zanamivir or relenza or "GG 167" or GG167 or GG-167 or CS-8958 or Dectova or Laninamivir or R-125489 or R125489 or "R 125489" or Inavir or peramivir or "BCX 1812" or BCX1812 or BCX-1812 or "RWJ 270201" or RWJ270201 or RWJ-270201 or Rapivab or rapiacta ) OR AB ( oseltamivir or tamiflu or "GS 4104" or GS4104 or GS-4104 or "GS 4071" or GS4071 or GS-4071 or zanamivir or relenza or "GG 167" or GG167 or GG-167 or CS-8958 or Dectova or Laninamivir or R-125489 or R125489 or "R 125489" or Inavir or peramivir or "BCX 1812" or BCX1812 or BCX-1812 or "RWJ 270201" or RWJ270201 or RWJ-270201 or Rapivab or rapiacta ) |
| S30 | (MH "Oseltamivir") |
| S29 | TI(neuraminidase inhibitor* or NA inhibitor*) OR AB(neuraminidase inhibitor* or NA inhibitor*) |
| S28 | TI Antiviral* OR AB Antiviral* |
| S27 | (MH "Antiviral Agents") |
| S26 | S24 OR S25 |
| S25 | TI ( Influenza or flu or H1N1 or PH1N1 or H3N2 or AH1N1 or AH3N2 or H5N1 or H7N9 ) OR AB ( Influenza or flu or H1N1 or PH1N1 or H3N2 or AH1N1 or AH3N2 or H5N1 or H7N9 ) |
| S24 | (MH "Influenza+") OR (MH "Influenza A Virus+") OR (MH "Influenzavirus C") OR (MH "Influenza B Virus") |
| S23 | S22 NOT S21 |
| S22 | S1 OR S2 OR S3 OR S4 OR S5 OR S6 OR S7 OR S8 OR S9 OR S10 OR S11 OR S12 OR S13 OR S14 OR S15 |
| S21 | S19 NOT S20 |
| S20 | MH (human) |
| S19 | S16 OR S17 OR S18 |
| S18 | TI (animal model*) |
| S17 | MH (animal studies) |
| S16 | MH animals+ |
| S15 | AB (cluster W3 RCT) |
| S14 | MH (crossover design) OR MH (comparative studies) |
| S13 | AB (control W5 group) |
| S12 | PT (randomized controlled trial) |
| S11 | MH (placebos) |
| S10 | MH (sample size) AND AB (assigned OR allocated OR control) |
| S9 | TI (trial) |
| S8 | AB (random*) |
| S7 | TI (randomised OR randomized) |
| S6 | MH cluster sample |
| S5 | MH pretest‐posttest design |
| S4 | MH random assignment |
| S3 | MH single‐blind studies |
| S2 | MH double‐blind studies |
| S1 | MH randomized controlled trials |

**Epistemonikos**

Influenza antivirals

**ClinIcaltrial.gov**

Influenza antivirals

### Appendix 2. Details of methods

#### 2.1. Details of data extraction

Pairs of reviewers independently extracted the following information: study characteristics (first author, trial registration, publication year, publication status, country, and sample size); patient characteristics (age, sex, disease severity, comorbidities, type of influenza); characteristics of interventions (dosing, frequency, route of administration, treatment duration, and length of follow-up); and outcome data of interest. Reviewers resolved discrepancies by discussion or, if necessary, with the assistance of a third party for adjudication.

#### 2.2. Details of subgroup analysis

To explore differences in pooled effect estimates related to different subgroups, we planned to perform subgroup analyses or meta-regression analyses with an interaction test using pairwise comparisons unless the same subgroup was present across all comparisons. When possible, we prioritized the use of within rather than between trial analyses. If data were available (at least two trials providing relevant information for each subgroup), we performed the following prespecified subgroup analyses:

1. Influenza etiology: seasonal versus zoonotic versus pandemic influenza viruses (hypothesis: reduced treatment effect in patients with zoonotic influenza virus infection than seasonal or pandemic influenza virus infection).
2. Confirmed versus suspected influenza virus infection: (hypothesis: reduced treatment effect in patients with suspected influenza compared to patients with confirmed influenza virus infection).
3. Age: children < 2 years, children versus adults and adolescents versus elderly (≥ 65 years) (hypothesis: reduced treatment effect in elderly).
4. Patients at increased risk of severe complications (people who are pregnant or up to 2 weeks postpartum; obesity (BMI > 40); patients with underlying health conditions – including chronic respiratory, cardiovascular disease; patients who are immunosuppressed): patients at increased risk of poor outcomes versus not at increased risk (hypothesis: reduced treatment effect in patients at increased risk of poor outcomes).

Due to limited within-trial subgroup information available, we also performed network meta-regression analyses for mean age, proportion of patients with influenza A, time from onset of symptoms to treatment, and proportion of patients receiving influenza vaccination, and conducted sensitivity analysis only including studies with confirmed influenza patients and studies with patients at high-risk. For mortality and admission to hospital, to assess the robustness of results, we performed sensitivity analyses calculating the RDs with 95% CIs.

#### 2.3. Details of risk of bias assessment

Pairs of reviewers independently evaluated the risk of bias of eligible RCTs using a modified Cochrane risk of bias tool. The assessment included following domains: random sequence generation; allocation concealment; blinding of participants, healthcare providers, data collectors, outcome assessor/adjudicator, and data analysts; incomplete outcome data (≥ 10% missing data was considered high risk of bias); selective outcome reporting; and other sources of bias (i.e. baseline imbalance, early trial discontinuation). We rated each domain at the outcome level as either: low, probably low, probably high, or high risk of bias. Because lack of blinding is unlikely to bias assessment of mortality and admission to hospital and ICU, we rated the blinding for these outcomes as low risk of bias regardless of blinding status. Reviewers resolved discrepancies by discussion or, if necessary, with adjudication by a third party.

#### 2.4. Details of certainty of evidence rating

To assess the certainty of evidence for each outcome, we used the Grading of Recommendations Assessment, Development and Evaluation (GRADE) approach for network meta-analysis.^1,2^ We evaluated the risk of bias, inconsistency, indirectness, and publication bias, and rated the certainty of evidence for direct estimates as high, moderate, low, or very low.^3^

Certainty ratings of indirect estimates were based on the lowest rating of the direct comparisons that contributed to the most-dominant first-order loop. We further rated down for intransitivity only if there was evidence of plausible effect modification between the direct comparisons that inform the indirect comparison. To ensure the validity of the transitivity assumption underlying network meta-analysis, we compared the distribution of potential effect modifiers (i.e. age, influenza virus type, time from onset of symptoms to treatment, patients at high risk) across treatment comparisons.

For the certainty of network estimates, we started with the estimate - direct or indirect - that dominated the network estimate or, if they both contributed importantly to the network estimate, used the higher of the direct and indirect estimates. We assessed imprecision at the network level using the minimally important difference (MID) as a threshold.^4^ The guideline panel specified an MID as 0.3% for mortality, 1.5% for admission to hospital, 1% for admission to ICU, 5% for emergence of resistance, 1% for any adverse events and adverse events related to treatments, 0.5% for serious adverse events, and 1 day for time to alleviation of symptoms and duration of hospitalization. We rated imprecision following GRADE guidance and concept.^5,6^

If incoherence was present, we used the estimate with the higher certainty of direct and indirect evidence as the best estimate. We developed the summary of findings tables using optimal formats in MAGICapp, presenting both relative and absolute effects and including plain language summaries with wording following GRADE guidance.^7,8^

**References**

1. Puhan MA, Schünemann HJ, Murad MH, et al. A GRADE Working Group approach for rating the quality of treatment effect estimates from network meta-analysis. *BMJ* 2014; **349**: g5630.
2. Brignardello-Petersen R, Bonner A, Alexander PE, et al. Advances in the GRADE approach to rate the certainty in estimates from a network meta-analysis. *J Clin Epidemiol* 2018; **93**: 36-44.
3. Guyatt GH, Oxman AD, Vist GE, et al. GRADE: an emerging consensus on rating quality of evidence and strength of recommendations. *BMJ* 2008; **336**(7650): 924-6.
4. Zeng L, Brignardello-Petersen R, Hultcrantz M, et al. GRADE guidelines 32: GRADE offers guidance on choosing targets of GRADE certainty of evidence ratings. *J Clin Epidemiol* 2021; **137**: 163-75.
5. Zeng L, Brignardello-Petersen R, Hultcrantz M, et al. GRADE Guidance 34: update on rating imprecision using a minimally contextualized approach. *J Clin Epidemiol* 2022; **150**: 216-24.
6. Hao Q, Gao Y, Zhao Y, et al. GRADE concept 6: a novel application of external indirect evidence into GRADE ratings of evidence certainty in network meta-analysis. *J Clin Epidemiol* 2023; **163**: 95-101.
7. Carrasco-Labra A, Brignardello-Petersen R, Santesso N, et al. Improving GRADE evidence tables part 1: a randomized trial shows improved understanding of content in summary of findings tables with a new format. *J Clin Epidemiol* 2016; **74**: 7-18.
8. Santesso N, Glenton C, Dahm P, et al. GRADE guidelines 26: informative statements to communicate the findings of systematic reviews of interventions. *J Clin Epidemiol* 2020; **119**: 126-35.

### Appendix 3. References of included studies

1. Anonymous. Randomised trial of efficacy and safety of inhaled zanamivir in treatment of influenza A and B virus infections. The MIST (Management of Influenza in the Southern Hemisphere Trialists) Study Group. Lancet (London, England) 1998; 352(9144): 1877-81.
2. Bai X, Xi S, Chen G, et al. Multicenter, randomized controlled, open label evaluation of the efficacy and safety of arbidol hydrochloride tablets in the treatment of influenza-like cases. BMC infectious diseases 2023; 23(1): 585.
3. Baker J, Block SL, Matharu B, et al. Baloxavir Marboxil Single-dose Treatment in Influenza-infected Children: A Randomized, Double-blind, Active Controlled Phase 3 Safety and Efficacy Trial (miniSTONE-2). The Pediatric infectious disease journal 2020; 39(8): 700-5.
4. Beigel JH, Manosuthi W, Beeler J, et al. Effect of Oral Oseltamivir on Virological Outcomes in Low-risk Adults With Influenza: A Randomized Clinical Trial. Clinical infectious diseases: an official publication of the Infectious Diseases Society of America 2020; 70(11): 2317-24.
5. Butler CC, van der Velden AW, Bongard E, et al. Oseltamivir plus usual care versus usual care for influenza-like illness in primary care: an open-label, pragmatic, randomised controlled trial. Lancet (London, England) 2020; 395(10217): 42-52.
6. Dharan NJ, Fry AM, Kieke BA, et al. Clinical and virologic outcomes in patients with oseltamivir-resistant seasonal influenza A (H1N1) infections: results from a clinical trial. Influenza and other respiratory viruses 2012; 6(3): 153-8.
7. Duval X, van der Werf S, Blanchon T, et al. Efficacy of oseltamivir-zanamivir combination compared to each monotherapy for seasonal influenza: a randomized placebo-controlled trial. PLoS medicine 2010; 7(11): e1000362.
8. Escuret V, Cornu C, Boutitie F, et al. Oseltamivir-zanamivir bitherapy compared to oseltamivir monotherapy in the treatment of pandemic 2009 influenza A(H1N1) virus infections. Antiviral research 2012; 96(2): 130-7.
9. Fan HW, Han Y, Liu W, et al. [A randomized controlled study of peramivir, oseltamivir and placebo in patients with mild influenza]. Zhonghua Nei Ke Za Zhi 2019; 58(8): 560-5.
10. Fry AM, Goswami D, Nahar K, et al. Efficacy of oseltamivir treatment started within 5 days of symptom onset to reduce influenza illness duration and virus shedding in an urban setting in Bangladesh: a randomised placebo-controlled trial. The Lancet Infectious diseases 2014; 14(2): 109-18.
11. Galbraith AW, Oxford JS, Schild GC, Potter CW, Watson GI. Therapeutic effect of 1-adamantanamine hydrochloride in naturally occurring influenza A 2 -Hong Kong infection. A controlled double-blind study. Lancet (London, England) 1971; 2(7716): 113-5.
12. Galbraith AW, Schild GC, Potter CW, Watson GI. The therapeutic effect of amantadine in influenza occurring during the winter of 1971-2 assessed by double-blind study. The Journal of the Royal College of General Practitioners 1973; 23(126): 34-7.
13. Hayden FG, Gubareva LV, Monto AS, et al. Inhaled zanamivir for the prevention of influenza in families. New England Journal of Medicine 2000; 343(18): 1282-9.
14. Hayden FG, Lenk RP, Stonis L, Oldham-Creamer C, Kang LL, Epstein C. Favipiravir Treatment of Uncomplicated Influenza in Adults: Results of Two Phase 3, Randomized, Double-Blind, Placebo-Controlled Trials. The Journal of infectious diseases 2022; 226(10): 1790-9.
15. Hayden FG, Osterhaus AD, Treanor JJ, et al. Efficacy and safety of the neuraminidase inhibitor zanamivir in the treatment of influenzavirus infections. GG167 Influenza Study Group. The New England journal of medicine 1997; 337(13): 874-80.
16. Hayden FG, Sugaya N, Hirotsu N, et al. Baloxavir Marboxil for Uncomplicated Influenza in Adults and Adolescents. The New England journal of medicine 2018; 379(10): 913-23.
17. Hedrick JA, Barzilai A, Behre U, et al. Zanamivir for treatment of symptomatic influenza A and B infection in children five to twelve years of age: a randomized controlled trial. The Pediatric infectious disease journal 2000; 19(5): 410-7.
18. Heinonen S, Silvennoinen H, Lehtinen P, et al. Early oseltamivir treatment of influenza in children 1-3 years of age: a randomized controlled trial. Clinical infectious diseases: an official publication of the Infectious Diseases Society of America 2010; 51(8): 887-94.
19. Hirotsu N, Saisho Y, Hasegawa T, Shishido T. Clinical and virologic effects of four neuraminidase inhibitors in influenza A virus-infected children (aged 4-12 years): an open-label, randomized study in Japan. Expert review of anti-infective therapy 2018; 16(2): 173-82.
20. Hsieh Y-H, Dugas AF, LoVecchio F, et al. Intravenous peramivir vs oral oseltamivir in high-risk emergency department patients with influenza: Results from a pilot randomized controlled study. Influenza and other respiratory viruses 2021; 15(1): 121-31.
21. Ison MG, Portsmouth S, Yoshida Y, et al. Early treatment with baloxavir marboxil in high-risk adolescent and adult outpatients with uncomplicated influenza (CAPSTONE-2): a randomised, placebo-controlled, phase 3 trial. The Lancet Infectious diseases 2020; 20(10): 1204-14.
22. Johnston SL, Ferrero F, Garcia ML, Dutkowski R. Oral oseltamivir improves pulmonary function and reduces exacerbation frequency for influenza-infected children with asthma. The Pediatric infectious disease journal 2005; 24(3): 225-32.
23. Kashiwagi S, Kudoh S, Watanabe A, Yoshimura I. [Clinical efficacy and safety of the selective oral neuraminidase inhibitor oseltamivir in treating acute influenza--placebo-controlled double-blind multicenter phase III trial]. Kansenshogaku zasshi The Journal of the Japanese Association for Infectious Diseases 2000; 74(12): 1044-61.
24. Kato M, Saisho Y, Tanaka H, Bando T. Improvement of respiratory symptoms and health-related quality of life with peramivir in influenza patients with chronic respiratory disease: Additional outcomes of a randomized, open-label study. Influenza and other respiratory viruses 2021; 15(5): 651-60.
25. Kato M, Saisho Y, Tanaka H, Bando T. Effect of peramivir on respiratory symptom improvement in patients with influenza virus infection and pre-existing chronic respiratory disease: Findings of a randomized, open-label study. Influenza and other respiratory viruses 2021; 15(1): 132-41.
26. Katsumi Y, Otabe O, Matsui F, et al. Effect of a single inhalation of laninamivir octanoate in children with influenza. Pediatrics 2012; 129(6): e1431-6.
27. Kiselev OI, Maleev VV, Deeva EG, et al. [Clinical efficacy of arbidol (umifenovir) in the therapy of influenza in adults: preliminary results of the multicenter double-blind randomized placebo-controlled study ARBITR]. Terapevticheskii arkhiv 2015; 87(1): 88-96.
28. Kohno S, Kida H, Mizuguchi M, Shimada J. Efficacy and safety of intravenous peramivir for treatment of seasonal influenza virus infection. Antimicrobial agents and chemotherapy 2010; 54(11): 4568-74.
29. Kohno S, Yen M-Y, Cheong H-J, et al. Phase III randomized, double-blind study comparing single-dose intravenous peramivir with oral oseltamivir in patients with seasonal influenza virus infection. Antimicrobial agents and chemotherapy 2011; 55(11): 5267-76.
30. Li LY, Cai BQ, Wang MZ, Zhu YJ. A double-blind, randomized, placebo-controlled multicenter study of oseltamivir phosphate for treatment of influenza infection in China. Chinese Medical Journal 2003; 116(1): 44-8.
31. Li X, Bilcke J, van der Velden AW, et al. Direct and Indirect Costs of Influenza-Like Illness Treated with and Without Oseltamivir in 15 European Countries: A Descriptive Analysis Alongside the Randomised Controlled ALIC4E Trial. Clinical drug investigation 2021; 41(8): 685-99.
32. Li Z, Zhang H, Chen J, Zhang Q, Xie L, Li X. Curative effect of peramivir and oseltamivir on influenza viral pneumonia. Chinese Journal of Nosocomiology 2021; 31(24): 3717-21.
33. Lin J-T, Yu X-Z, Cui D-J, et al. A multicentre, randomized, controlled trial of oseltamivir in the treatment of influenza in a high-risk Chinese population. Current medical research and opinion 2006; 22(1): 75-82.
34. Makela MJ, Pauksens K, Rostila T, et al. Clinical efficacy and safety of the orally inhaled neuraminidase inhibitor zanamivir in the treatment of influenza: a randomized, double-blind, placebo-controlled European study. The Journal of infection 2000; 40(1): 42-8.
35. Matsumoto K, Ogawa N, Nerome K, et al. Safety and efficacy of the neuraminidase inhibitor zanamivir in treating influenza virus infection in adults: results from Japan. GG167 Group. Antiviral therapy 1999; 4(2): 61-8.
36. McLean HQ, Belongia EA, Kieke BA, Meece JK, Fry AM. Impact of Late Oseltamivir Treatment on Influenza Symptoms in the Outpatient Setting: Results of a Randomized Trial. Open forum infectious diseases 2015; 2(3): ofv100.
37. Monto AS, Fleming DM, Henry D, et al. Efficacy and safety of the neuraminidase inhibitor zanamivir in the treatment of influenza A and B virus infections. The Journal of infectious diseases 1999; 180(2): 254-61.
38. Murphy KR, Eivindson A, Pauksens K, et al. Efficacy and safety of inhaled zanamivir for the treatment of influenza in patients with asthma or chronic obstructive pulmonary disease: A double-blind, randomised, placebo-controlled, multicentre study. Clinical Drug Investigation 2000; 20(5): 337-49.
39. Nakamura S, Miyazaki T, Izumikawa K, et al. Efficacy and Safety of Intravenous Peramivir Compared With Oseltamivir in High-Risk Patients Infected With Influenza A and B Viruses: A Multicenter Randomized Controlled Study. Open forum infectious diseases 2017; 4(3): ofx129.
40. Nicholson KG, Aoki FY, Osterhaus AD, et al. Efficacy and safety of oseltamivir in treatment of acute influenza: a randomised controlled trial. Neuraminidase Inhibitor Flu Treatment Investigator Group. Lancet (London, England) 2000; 355(9218): 1845-50.
41. Popov AF, Shchelkanov MY, Dmitrenko KA, Simakova AI. Combined therapy of influenza with antiviral drugs with a different mechanism of action in comparison with monotherapy. Journal of Pharmaceutical Sciences and Research 2018; 10(2): 357-60.
42. Portsmouth S, Hayden FG, Kawaguchi K, et al. Baloxavir Treatment in Adolescents With Acute Influenza: Subgroup Analysis From the CAPSTONE-1 Trial. Journal of the Pediatric Infectious Diseases Society 2021; 10(4): 477-84.
43. Puhakka T, Lehti H, Vainionpaa R, et al. Zanamivir: a significant reduction in viral load during treatment in military conscripts with influenza. Scandinavian journal of infectious diseases 2003; 35(1): 52-8.
44. Roberts G, Chen S, Yates P, et al. Randomized, Double-Blind, Placebo-Controlled Study of the Safety, Tolerability, and Clinical Effect of Danirixin in Adults With Acute, Uncomplicated Influenza. Open forum infectious diseases 2019; 6(4): ofz072.
45. Sugaya N, Ohashi Y. Long-acting neuraminidase inhibitor laninamivir octanoate (CS-8958) versus oseltamivir as treatment for children with influenza virus infection. Antimicrobial agents and chemotherapy 2010; 54(6): 2575-82.
46. Takazono T, Miyazaki T, Yoshida Y, Kojima S, Hosogaya N, Mukae H. Efficacy and safety of baloxavir marboxil in influenza patients aged 75 or over: a sub-group analysis of CAPSTONE-2 study. Journal of the Japanese Association for Infectious Diseases 2021; 95(1): 1-8.
47. Treanor JJ, Hayden FG, Vrooman PS, et al. Efficacy and safety of the oral neuraminidase inhibitor oseltamivir in treating acute influenza: a randomized controlled trial. US Oral Neuraminidase Study Group. JAMA 2000; 283(8): 1016-24.
48. Wang MZ, Cai B1, Li LY, et al. [Efficacy and safety of arbidol in treatment of naturally acquired influenza]. Zhongguo Yi Xue Ke Xue Yuan Xue Bao 2004; 26(3): 289-93.
49. Watanabe A. A randomized double-blind controlled study of laninamivir compared with oseltamivir for the treatment of influenza in patients with chronic respiratory diseases. Journal of infection and chemotherapy: official journal of the Japan Society of Chemotherapy 2013; 19(1): 89-97.
50. Watanabe A, Chang S-C, Kim MJ, Chu DW-S, Ohashi Y. Long-acting neuraminidase inhibitor laninamivir octanoate versus oseltamivir for treatment of influenza: A double-blind, randomized, noninferiority clinical trial. Clinical infectious diseases: an official publication of the Infectious Diseases Society of America 2010; 51(10): 1167-75.
51. Watanabe A, Ishida T, Hirotsu N, et al. Baloxavir marboxil in Japanese patients with seasonal influenza: Dose response and virus type/subtype outcomes from a randomized phase 2 study. Antiviral research 2019; 163(6i7, 8109699): 75-81.
52. Whitley RJ, Hayden FG, Reisinger KS, et al. Oral oseltamivir treatment of influenza in children. The Pediatric infectious disease journal 2001; 20(2): 127-33.
53. NCT00705406. A Phase II, Multicenter, Randomized, Placebo -Controlled, Study To Evaluate The Efficacy and Safety Of Intramuscular Peramivir 600 mg In Subjects With Uncomplicated Acute Influenza. 2009. https://clinicaltrials.gov/study/NCT00705406.
54. NCT02369159. A Phase 3, Randomized, Open Label, Active-controlled Study to Evaluate the Safety, Pharmacokinetics and Effectiveness of IV Peramivir Compared to Oral Oseltamivir in Pediatric Subjects With Acute Uncomplicated Influenza. 2015. https://clinicaltrials.gov/study/NCT02369159.
55. NCT00610935. Intramuscular Peramivir in Subjects With Uncomplicated Acute Influenza. 2008. https://clinicaltrials.gov/study/NCT00610935?term=NCT00610935&rank=1.
56. NCT00419263. To evaluate the efficacy of peramivir administered intramuscularly compared to placebo in adult subjects with uncomplicated acute influenza. 2007. https://clinicaltrials.gov/study/NCT00419263?tab=results.
57. NCT01068912. Dose-Finding Study of Favipiravir in the Treatment of Uncomplicated Influenza. 2010. https://clinicaltrials.gov/study/NCT01068912?term=NCT01068912&rank=1&tab=results.
58. NCT01793883. Efficacy and Safety Study of Laninamivir Octanoate TwinCaps® Dry Powder Inhaler in Adults With Influenza (Igloo). 2013. https://clinicaltrials.gov/study/NCT01793883?intr=Laninamivir&rank=2.
59. Man CY, Keene ON, Challoner T (Challoner Associates). A double-blind, randomised, placebo-controlled, parallel-group, multicentre study to investigate the efficacy and safety of inhaled zanamivir (GG167) 10 mg administered twice a day for five days in the treatment of symptomatic influenza A and B viral infections in adolescents and adults. 1998. https://www.gsk-studyregister.com/en/trial-details/?id=NAIB3002.
60. GlaxoSmithKline. Investigation of the efficacy of CG167 in the treatment of influenza viral infections (phase II study) (Protocol No. JNAI-01). Double blind, double dummy, randomized, placebo controlled, parallel group, multicenter study to investigate safety and route of administration of CG167 when inhaled, CG167 10 mg, or the combination of inhaled CG167 10 mg plus intranasal CG167 6.4 mg, administered twice daily for 5 days in the treatment of influenza A and B viral infections. 2000. https://www.gsk-studyregister.com/en/trial-details/?id=JNAI-01.
61. GlaxoSmithKline. A multicenter two way layout randomized placebo controlled double-blind trial parallel group comparative trial on the efficacy and safety of GG167 (zanamivir) 10 mg twice a day and 20 mg twice a day in the treatment of influenza type A and type B infections (late Phase II study: dose comparison study) (Protocol No. JNAI-04). 1999. https://www.gsk-studyregister.com/en/trial-details/?id=JNAI-04.
62. GlaxoSmithKline. A multicenter two-way layout randomized placebo-controlled double-blind trial parallel group comparative trial on the efficacy and safety of GG167 (zanamivir) 10 mg twice a day and 20 mg twice a day in the treatment of influenza type A and type B infections (late Phase II study: dose comparison study) (Protocol No. JNAI-07). 1999. https://www.gsk-studyregister.com/en/trial-details/?id=JNAI-07.
63. Roche Pharmaceuticals. Clinical Study Report – Protocol JV15823 - A randomized, placebo-controlled, multicenter study of oseltamivir (Ro 64-0796) in the treatment of influenza in Japanese subjects. (Translation of summary of Japanese report.) 2000. https://datadryad.org/stash/dataset/doi:10.5061/dryad.77471.
64. McGarty T. Final clinical study report—protocol M76001: a randomized, double-blind, placebo-controlled, multicenter study of efficacy based on the time to treatment of influenza infection with the nuraminidase inhibitor Ro 64-0796 (also known as GS 4104). 2000. https://datadryad.org/stash/dataset/doi:10.5061/dryad.77471.
65. GlaxoSmithKline. A double-blind, randomized, placebo controlled, parallel group, multi-center study to investigate the efficacy and safety of inhaled zanamivir 10 mg administered twice daily for five days in the treatment of influenza in patients 12 years or over diagnosed with asthma or chronic obstructive pulmonary disease. 2000. https://www.gsk-studyregister.com/en/trial-details/?id=NAI30008.
66. Alfors S, Keene O, Grice R, Hammond J, Hendricks V, Martin N, et al. A double-blind, randomized, placebo-controlled, parallel group, multicenter study to investigate the efficacy and safety of zanamivir (GG167) 10 mg administered by inhalation twice daily for five days in the treatment of symptomatic influenza A and B viral infections in children ages 5-12. 1999. https://www.gsk-studyregister.com/en/trial-details/?id=NAI30009.
67. Hunter S, Reilly L, Sharp S, West M, Alfors S, Hammond J, et al. A double-blind, randomized, placebo-controlled, parallel group, multicenter study to investigate the efficacy and safety of inhaled zanamivir (GG167) 10 mg administered once a day for 10 days in the prevention of transmission of symptomatic influenza A and B viral infections within families. 1999. https://www.gsk-studyregister.com/en/trial-details/?id=NAI30010.
68. GlaxoSmithKline. A randomized, double-blind, placebo-controlled study to evaluate the impact of inhaled zanamivir treatment on workplace attendance and healthcare outcomes due to influenza A and B infections. 2001. https://www.gsk-studyregister.com/en/trial-details/?id=NAI30011.
69. GlaxoSmithKline. A Double-Blind, Randomised, Placebo-Controlled, Parallel-Group, Multicentre Study to Investigate the Efficacy and Safety of Inhaled Zanamivir 10mg Administered Twice Daily for Five Days in the Treatment of Symptomatic Influenza A and B Viral Infections in Subjects Aged >= 65 Years. 2002. https://www.gsk-studyregister.com/en/trial-details/?id=NAI30012.
70. GlaxoSmithKline. A double-blind, randomised, placebo-controlled, multicenter study in 2 parallel groups, to investigate the efficacy and safety of inhaled Zanamivir (10 mg bd. via Diskhaler), for 5 days, in high risk patients with symptomatic Influenza A and/or B infection. 2002. https://www.gsk-studyregister.com/en/trial-details/?id=NAI30020.
71. GlaxoSmithKline. A double-blind, randomised, placebo-controlled, multicenter study in 2 parallel groups, to investigate the efficacy and safety of inhaled zanamivir (10 mg bd via Diskhaler), for 5 days, in children aged 5 to 12 years with symptomatic Influenza A and/or B infection. 2003. https://www.gsk-studyregister.com/en/trial-details/?id=NAI30028.
72. GlaxoSmithKline. A double-blind, randomized, placebo controlled, multicenter, parallel-group study to investigate the efficacy and safety of zanamivir administered twice or four times a day for the treatment of influenza A and B viral infections. 1998. https://www.gsk-studyregister.com/en/trial-details/?id=NAIA2008; https://www.gsk-studyregister.com/en/trial-details/?id=NAIB2008.
73. MacLeod A, Gummer M, Raniga K, Hirst H, Keene O, Ossi M, et al. A double-blind, randomised, placebo-controlled multicentre study to investigate the efficacy and safety of inhaled and intranasal zanamivir in the treatment of influenza A and B viral infections. 1998. https://www.gsk-studyregister.com/en/trial-details/?id=NAIA2005.
74. Elliott M, Flack N, Keene O, Szymborski P, Vega R (PharmaResearch, Inc). A double-blind, randomized, placebo-controlled, parallel-group, multicenter study to investigate the efficacy and safety of inhaled zanamivir (GG167) 10 mg administered twice a day for five days in the treatment of symptomatic influenza A and B viral infections in adolescents and adults. 1998. https://www.gsk-studyregister.com/en/trial-details/?id=NAIA3002.
75. Leong J, Brennan J, Gummer M, Keene O, Wightman K. A double-blind, randomised, placebo-controlled, parallel-group, multi-centre study to investigate the efficacy and safety of inhaled plus intranasal zanamivir in the treatment of influenza A and B viral infections. 1998. https://www.gsk-studyregister.com/en/trial-details/?id=NAIB2005.
76. Perich R, Solterbeck A, Keene O, Leong J, Raniga K, MacLeod A. A double-blind, randomised, placebo-controlled, parallel-group, multi-centre study to investigate the efficacy and safety of inhaled and inhaled plus intranasal zanamivir in the treatment of influenza A and B viral infections. 1998. https://www.gsk-studyregister.com/en/trial-details/?id=NAIB2007.
77. Campion K, Gummer M, Keene O. A double-blind, randomized, placebo-controlled, parallel-group, multicenter study to investigate the efficacy and safety of zanamivir administered twice daily in the treatment of influenza A and B viral infections in adults. 1998. https://www.gsk-studyregister.com/en/trial-details/?id=NAIB3001.
78. Roche Pharmaceuticals. Clinical Study Report – Protocol NV16871: A double-blind, randomized, stratified, placebo –controlled study of oseltamivir in the treatment of influenza in children with asthma. 2004. https://datadryad.org/stash/dataset/doi:10.5061/dryad.77471.
79. Dorkings J. Final clinical study report—protocol WV15670: a double-blind, randomized, placebo controlled study of oral Ro 64-0796 (also known as GS 4104) in the treatment of influenza infection. 1998. https://datadryad.org/stash/dataset/doi:10.5061/dryad.77471.
80. Dorkings J. Clinical study report—protocol GS97-803 (WV15671): a double-blind, randomized, placebo controlled study of GS 4104 (Ro 64-0796) in the treatment of influenza infection. 1999. https://datadryad.org/stash/dataset/doi:10.5061/dryad.77471.
81. Grosse M. Clinical study report—protocol WV15707: a double-blind, stratified, randomized, placebo controlled study of Ro 64-0796 (GS4104) in the treatment of influenza infection in elderly adults. 1999. https://datadryad.org/stash/dataset/doi:10.5061/dryad.77471.
82. Dorkings J. Clinical study report—protocol WV15730: a double-blind, stratified, randomized, placebo controlled study of Ro 64-0796 (GS4104) in the treatment of influenza infection in adults. 1999. https://datadryad.org/stash/dataset/doi:10.5061/dryad.77471.
83. Roche Pharmaceuticals. Clinical Study Report - Protocol WV15758. A double-blind, randomized, stratified, placebo-controlled study of Ro 64-0796 (also known as GS 4104) in the treatment of children with influenza. 2000. https://datadryad.org/stash/dataset/doi:10.5061/dryad.77471.
84. Gerster T. A double-blind, randomized, stratified, placebo controlled study of oseltamivir phosphate (Ro 64-0796, also known as GS 4104) in the treatment of influenza in children with chronic asthma. 2000. https://datadryad.org/stash/dataset/doi:10.5061/dryad.77471.
85. McCarvil M. Final clinical study report (protocols WV15812 and WV15872)—a double-blind, stratified, randomised, placebo controlled study of Ro 64-0796 (also known as GS4104) in the treatment of influenza in chronically ill adults. 2000. https://datadryad.org/stash/dataset/doi:10.5061/dryad.77471.
86. Roche Pharmaceuticals. Final clinical study report—protocols WV15819, WV15876 and WV15978: a double- blind, randomized, stratified, placebo-controlled study of Ro 64-0796 (also known as GS4104) in the treatment of influenza infection in elderly patients. 2000. https://datadryad.org/stash/dataset/doi:10.5061/dryad.77471.
87. Roche Pharmaceuticals. Clinical study report—protocol WV16277: a double-blind, randomized, stratified, placebo-controlled study of oseltamivir in the treatment of influenza infection in patients. 2003. https://datadryad.org/stash/dataset/doi:10.5061/dryad.77471.

### Appendix 4: Basic characteristics of eligible studies

| **Study** | **Publication status** | **Country** | **Patients randomized** | **Mean age (years)** | **Age range (years)** | **Male %** | **Pregnant %** | **High risk patients %** | **Comorbidities %** | **Type of influenza (%)** | **Confirmed influenza %** | **Time from onset of symptoms to treatment/random (days)** | **Influenza vaccination %** | **Treatments** |
| --- | --- | --- | --- | --- | --- | --- | --- | --- | --- | --- | --- | --- | --- | --- |
| MIST 1998, NAIB3001 | Peer-reviewed publication | Australia, New Zealand, South Africa | 455 | 36.95 | ≥ 12 | 52.97 | 0 | 16.70 | 12.53 (respiratory disorders, mostly mild asthma), 1.76 (cardiovascular disorder other than hypertension) | 47.03 (A), 23.52 (B) | 70.55 | NR | 5.71 | Zanamivir inhaled 10 mg twice a day for 5 days Placebo |
| Bai 2023 | Peer-reviewed publication | China | 412 | 33.00 | 18–87 | 52.63 | 0 | NR | (0.24) interstitial pneumonia, 0.49 (coronary heart disease), 0.97 (diabetes), 2.67 (hypertension) | NR | NR | NR | 0 | Oseltamivir oral 75 mg twice a day for 5 days Umifenovir 200 mg three times a day for 5 days |
| Baker 2020 | Peer-reviewed publication | USA, Poland, Spain, Costa Rica, Mexico, Russia | 176 | 6.10 | 1–12 | 46.80 | 0 | NR | NR | 16.18 (A/H1N1 pdm09), 43.93 (A/H3N2), 4.05 (B), 0.58 (mixed), 2.31 (unknown) | 67.05 | NR | 49.13 | Baloxavir oral a single dose on day 1 (2 mg/kg for those weighing <20 kg and a single dose of 40 mg for those weighing ≥20 kg) Oseltamivir oral twice a day for 5 days. 30 mg for patients weighing ≤15 kg, 45 mg for >15–≤23 kg, 60 mg for >23–≤40 kg and 75 mg for >40 kg |
| Beigel 2020 | Peer-reviewed publication | Thailand, USA, Argentina | 556 | 36.67 | 18–64 | 37.59 | 0 | 0 | NR | 45.14 (H3N2), 16.73 (H1N1), 28.06 (B), 0.18 (coinfection) | 90.11 | 1.21 | 10.25 | Oseltamivir oral 75 mg twice a day for 5 days Placebo |
| Butler 2020, Li 2021 (a) | Peer-reviewed publication | 15 European countries | 3266 | 35.45 | ≥ 1 | 44.12 | 0 | NR | 6.01 (chronic respiratory condition), 4.45 (heart disease), 2.52 (diabetes) | 29.09 (A), 22.28 (B) | 51.34 | NR | 9.42 | Oseltamivir oral 75 mg twice a day for 5 days for patients aged 13 years or more. For children younger than 13 years, oseltamivir was given in oral suspension according to weight; children weighing 10–15 kg received 30 mg, >15–23 kg received 45 mg, >23–40 kg received 60 mg, and >40 kg received 75 mg Standard care |
| Dharan 2012 | Peer-reviewed publication | USA | 19 | 9.00 | 1–54 | 52.63 | NR | NR | NR | 100 (seasonal influenza A/H1N1) | 100 | 2.90 | NR | Oseltamivir oral 75 mg twice a day for 5 days Placebo |
| Duval 2010 | Peer-reviewed publication | France | 541 | 39.34 | 18–84.2 | 49.72 | 0 | NR | NR | 3.88 (A/H1N1), 73.01 (A/H3N2), 5.73 (A/not determined) | 100 | NR | 0 | Oseltamivir oral 75 mg twice a day for 5 days Zanamivir inhaled 10 mg once a day for 5 days Oseltamivir oral 75 mg twice a day for 5 days plus zanamivir inhaled 10 mg once a day for 5 days |
| Escuret 2012 | Peer-reviewed publication | France | 24 | 34.40 | 19.1–55.2 | 41.67 | 0 | NR | NR | 100 (A/H1N1pmd09) | 100 | 1.02 | 0 | Oseltamivir oral 75 mg twice a day for 5 days Oseltamivir oral 75 mg twice a day for 5 days plus zanamivir inhaled 10 mg twice a day for 5 days |
| Fan 2019 | Peer-reviewed publication | China | 129 | 32.05 | 15–68 | 38.76 | 0 | NR | NR | NR | 100 | NR | NR | Oseltamivir oral 75 mg twice a day for 5 days Peramivir single intravenous 300 mg |
| Fry 2014 | Peer-reviewed publication | Bangladesh | 1190 | 5.00 | > 1 | 52.69 | 0 | NR | NR | 35.13 (A/H3N2), 11.01 (seasonal A/H1N1), 17.90 (A/H1N1 pdm09), 33.36 (B), 0.34 (mixed) | 100 | 2.00 | NR | Oseltamivir oral twice a day for 5 days; body weight <15 kg received 30 mg/dose (2.5 ml syrup), body weight 15 - 22 kg received 45 mg/dose (3.75ml syrup), body weight 23 - 39 kg received 60 mg/dose (5ml syrup), and body weight ≥ 40kg received 75 mg tablets Placebo |
| Galbraith 1971 | Peer-reviewed publication | UK | 153 | 38.30 | > 0 | 55.56 | NR | NR | NR | 100 (A2/Hong Kong/68) | 100 | NR | NR | Amantadine 100 mg capsules or as a syrup containing 50 mg in 5 ml. Adults received one capsule of soft gelatin containing a suspension of amantadine in oil every 12 hours. Children aged 10-15 years received one capsule every 24 hours and younger children (2-10 years) a proportional dose of syrup. for 7 days Placebo |
| Galbraith 1973 | Peer-reviewed publication | UK | 65 | 39.65 | > 2 | 60.00 | NR | NR | NR | 100 (A2/Hong Kong/68) | 100 | NR | 0 | Amantadine 100 mg capsules or as a syrup containing 50 mg in 5 ml. Adults received one capsule of soft gelatin containing a suspension of amantadine in oil every 12 hours. Children aged 10-15 years received one capsule every 24 hours and younger children (2-10 years) a proportional dose of syrup. for 7 days Placebo |
| Hayden 2018 (a), Watanabe 2019 | Peer-reviewed publication | Japan | 400 | 36.88 | 20–64 | 61.75 | 0 | 0 | No | 66.75 (A/H1N1 pdm09), 0.09 (A/H3N2), 22.75 (B), 1 (mixed), 0.05 (unknown) | 100 | NR | 30.50 | Baloxavir oral single dose of 10 mg Baloxavir oral single dose of 20 mg Baloxavir oral single dose of 40 mg Placebo |
| Hayden 2018 (b), Portsmouth 2021 | Peer-reviewed publication | USA, Japan | 1436 | 33.33 | 12–64 | 50.00 | 0 | 0 | No | 1.13 (A/H1N1 pdm09), 64.86 (A/H3N2), 6.48 (B), 1.20 (mixed), 1.27 (A/uncertain subtype) | 74.93 | NR | 21.62 | Baloxavir oral single dose of 40 mg for patients weighing <80 kg or 80 mg for those weighing 280 kg Oseltamivir oral 75 mg twice a day for 5 days Placebo |
| Hayden 2022 (a) | Peer-reviewed publication | Belgium, Bulgaria, Hungary, Poland, Russia, Spain, Sweden, Netherlands, Turkey, Ukraine, USA, Australia, New Zealand, South Africa | 855 | 41.30 | 18–80 | 40.93 | 0 | 0 | No | 7.13 (A/H1N1 pdm09), 55.20 (A/H3N2), 0.23 (A/H1 and A/H3), 10.29 (missing or negative subtyping) | 100 | 1.23 | 21.19 | Favipiravir 1800 mg administered twice on study day 1 (total dose 3600 mg), followed by 800 mg twice on days 2 to 5 (1600 mg/day) Placebo |
| Hayden 2022 (b) | Peer-reviewed publication | Argentina, Brazil, Canada, Colombia, the Dominican Republic, El Salvador, Guatemala, Mexico, Peru, USA, Puerto Rico | 1144 | 39.90 | 18–80 | 43.60 | 0 | 0 | No | 18.88 (A/H1N1 pdm09), 25.44 (A/H3N2), 16.43 (missing or negative subtyping) | 100 | 1.23 | 9.35 | Favipiravir 1800 mg administered twice on study day 1 (total dose 3600 mg), followed by 800 mg twice on days 2 to 5 (1600 mg/day) Placebo |
| Hedrick 2000, NAI30009 | Peer-reviewed publication | USA, Canada, UK, Belgium, Finland, Spain, Russia, Sweden, France, Germany, Israel | 471 | 8.71 | 5–12 | 54.78 | 0 | NR | 7.64 (respiratory diseases) | 47.98 (A), 25.05 (B) | 73.46 | 0.84 | 2.34 | Zanamivir inhaled 10 mg twice a day for 5 days Placebo |
| Heinonen 2010 | Peer-reviewed publication | Finland | 409 | 2.40 | 1–4 | 62.20 | 0 | NR | 13.30 (asthma) | 19.32 (A), 4.64 (B) | 23.96 | 0.40 | 13.27 | Oseltamivir oral twice a day for 5 days. 30 mg twice daily for children weighing ≤ 15.0 kg, 45 mg twice daily for children weighing 15.1–23.0 kg Placebo |
| Hirotsu 2017 | Peer-reviewed publication | Japan | 123 | NR | 4–12 | 51.75 | 0 | NR | No | 78.82 (H3N2), 15.79 (A/H1N1 pdm09), 1.75 (B) | 100 | NR | 57.89 | Oseltamivir oral twice a day at a dose of 2 mg/kg per administration (66.7 mg/kg as a dry syrup). In children weighing 37.5 kg or more, oral capsules of 75 mg each were administered orally twice a day for 5 days Peramivir intravenously infused 10 mg/kg (600 mg in patients whose body weight was 60 kg or more) over more than 15 min once and a repeated dose (treated for >1 day) was administered in accordance with persisting symptoms in pediatric patients Zanamivir inhaled 10 mg twice a day for 5 days Laninamivir inhalation a single dose of 40 mg for children aged 10 years and older and at a single dose of 20 mg for children under the age of 10 years |
| Hsieh 2019 | Peer-reviewed publication | USA | 180 | 50.00 | ≥ 18 | 40.80 | 0 | 67.60 | 60.33 (chronic pulmonary disease), 6.70 (pneumonia), 22.90 (cardiovascular disease) | 75.41 (A), 24.59 (B) | 100 | NR | 43.60 | Oseltamivir oral 30 mg once daily, 30 mg twice daily or 75 mg twice daily for 5 days Peramivir intravenous 100 mg, 200 mg, or 600 mg of one-time |
| Ison 2020, Takazono 2021 | Peer-reviewed publication | 17 countries | 2184 | 51.77 | 12–89 | 48.50 | 0 | 100 | 39.20 (asthma or chronic lung disease), 12.73 (cardiovascular disease) | 6.88 (A/H1N1pdm), 47.89 (A/H3N2), 41.62 (B), 1.2 (mixed infection), 2.4 (Other) | 100 | NR | 25.28 | Baloxavir oral a single dose of 40 mg for patients weighing <80 kg and 80 mg for those weighing ≥80 kg Placebo |
| Johnston 2005, WV15759/WV15871 | Peer-reviewed publication | Argentina, Australia, Belgium, Canada, Chile, Finland, Germany, China, Israel, New Zealand, Spain, South Africa, Sweden, UK, USA | 334 | 9.00 | 6–12 | 64.52 | 0 | 100 | 100 (asthma) | NR | 53.59 | 1.13 | 19.46 | Oseltamivir oral 2 mg/kg twice a day for 5 days Placebo |
| Kashiwagi 2000, JV15823 | Peer-reviewed publication | Japan | 316 | 34.09 | 16–89 | 48.24 | 0 | NR | NR | 0.64 (A/unknown), 17.25 (A/H1N1), 58.79 (A/H3N2), 3.51 (B), 0.32 (infection, unknown) | 80.50 | 0.98 | 0 | Oseltamivir oral 75 mg twice a day for 5 days Placebo |
| Kato 2021 | Peer-reviewed publication | Japan | 208 | NR | 16–79 | 44.71 | NR | 100 | 7.21 (COPD), 91.35 (bronchial asthma), 1.40 (pulmonary fibrosis), 1.91 (pneumonia), 1.43 (bronchitis) | 62.50 (A), 37.50 (B) | 100 | NR | 37.80 | Peramivir intravenous 600 mg repeat-dose (peramivir 600 mg for 2 days) Peramivir intravenous 300 mg single-dose for day 1 Oseltamivir oral 75 mg twice a day for 5 days |
| Katsumi 2012 | Peer-reviewed publication | Japan | 112 | 9.98 | ≤ 15 | 57.14 | 0 | NR | 27.68 (history of asthmatic symptoms) | 71.42 (A), 28.58 (B) | 100 | NR | 26.79 | Laninamivir inhaled a signle dose of 20 mg for patients <10 years of age and 40 mg for patients 10 to 15 years of age Zanamivir inhaled 10 mg twice a day for 5 days Placebo |
| Kiselev 2015 | Peer-reviewed publication | Russia | 119 | 37.61 | 18–65 | 37.82 | 0 | NR | 2.52 (respiratory disease), 14.29 (cardiovascular disease) | NR | 100 | NR | 0 | Umifenovir 200 mg three times a day for 5 days Placebo |
| Kohno 2010 | Peer-reviewed publication | Japan | 296 | 34.17 | 20–64 | 50.68 | 0 | NR | No | 72.64 (A/H1), 23.65 (A/H3), 2.70 (A/-), 1.01 (B) | 100 | NR | NR | Peramivir intravenous a single dose of 300 mg Peramivir intravenous a single dose of 600 mg Placebo |
| Kohno 2011 | Peer-reviewed publication | Japan, South Korea, China | 1099 | 35.13 | 20–80 | 51.52 | NR | NR | NR | 54.81 (A/H1), 0.09 (A/H1, H3), 30.06 (A/H3), 4.86 (A/-), 6.42 (B), 3.76 (unknown) | 100 | NR | 16.77 | Peramivir intravenous a single dose of 300 mg Peramivir intravenous a single dose of 600 mg Oseltamivir oral 75 mg twice a day for 5 days |
| Li 2004 | Peer-reviewed publication | China | 273 | 31.04 | 18–65 | 49.82 | 0 | NR | NR | 61.90 (A), 36.63 (B), 1.47 (unknown) | 100 | NR | NR | Oseltamivir oral 75 mg twice a day for 5 days Placebo |
| Lin 2006 | Peer-reviewed publication | China | 118 | 50.28 | NR | 58.93 | 0 | 100 | 56.90 (COPD), 20.69 (bronchial asthma), 3.45 (bronchiectasis), 13.79 (coronary heart disease), 1.72 (chronic heart insufficiency) | NR | 47.46 | NR | 0 | Oseltamivir oral 75 mg twice a day for 5 days Standard care |
| Li 2021 (b) | Peer-reviewed publication | China | 130 | 44.81 | ＞ 16 | 52.31 | NR | NR | NR | 53.85 (A), 46.15 (B) | 100 | NR | NR | Oseltamivir oral 75 mg twice a day for 5 days Peramivir intravenous 10 mg/kg/d up to 5 days |
| Makela 2000, NAIB3002 | Peer-reviewed publication | Belgium, Denmark, Finland, France, Germany, Holland, Italy, Norway, Spain, Sweden, UK | 356 | 37.20 | 12–81 | NR | 0 | 8.99 | NR | 74.44 (A/H3N2), 3.37 (B) | 77.81 | NR | NR | Zanamivir inhaled 10 mg twice a day for 5 days Placebo |
| Matsumoto 1999 | Peer-reviewed publication | Japan | 116 | 29.74 | 16–65 | 37.07 | 0 | NR | NR | 28.45 (A/H3N2), 34.48 (B) | 62.93 | NR | 0.86 | Zanamivir inhaled 10 mg twice a day for 5 days Zanamivir 10 mg zanamivir powder for inhalation plus 6.4 mg zanamivir nasal spray for days Placebo |
| McLean 2015 | Peer-reviewed publication | USA | 165 | 17.69 | 1–79 | 40.00 | NR | 11.52 | NR | 74.55 (A), 24.85 (B) | 100 | NR | NR | Oseltamivir oral 75 mg twice daily for 5 days for persons > 88 pounds. Children ≤ 88 pounds received a liquid form of oseltamivir at a concentration of 15 mg/ml Placebo |
| Monto 1999, NAIA/B2008 | Peer-reviewed publication | Belgium, Canada, Denmark, Finland, France, Germany, Italy, Netherlands, Norway, Spain, Sweden, USA, UK | 1256 | 35.01 | ≥ 13 | 44.03 | 0 | 12.56 | NR | 16.80 (A/H3N2), 15.61 (A/H1N1), 3.11 (B) | 57.00 | 1.21 | 1.51 | Zanamivir 10 mg oral inhalation plus 6.4 mg nasal spray twice a day for 5 days Zanamivir 10 mg oral inhalation plus 6.4 mg nasal spray four times a day for 5 days Placebo |
| Murphy 2000, NAI30008 | Peer-reviewed publication | USA, Canada, Europe, Australia, Chile, South Africa | 525 | 38.00 | ≥ 12 | 42.48 | 0 | 100 | 6.10 (asthma and COPD), 76.00 (asthma), 16.57 (COPD) | 54.10 (A), 5.30 (B) | 59.62 | NR | 23.24 | Zanamivir inhaled 10 mg twice a day for 5 days Placebo |
| Nakamura 2017 | Peer-reviewed publication | Japan | 92 | 71.15 | ≥ 20 | 46.74 | 0 | 100 | 42.39 (chronic respiratory illness), 15.22 (cardiovascular disease), 23.91 (diabetes) | 66.30 (A/H3N2), 19.57 (B), 11.96 (A/H1N1 pdm09), 2.17 (not detected) | 100 | 1.13 | 50.00 | Oseltamivir oral 75 mg twice a day for 5 days Peramivir intravenous a single of 600 mg (a second infusion at >2 days later, if necessary, was permitted) |
| Nicholson 2000, WV15670 | Peer-reviewed publication | Europe, Canada, China | 726 | 37.43 | 18–65 | 51.01 | 0 | NR | NR | 63.84 (A), 2.23 (B) | 66.06 | 1.00 | 0 | Oseltamivir oral 75 mg twice a day for 5 days Oseltamivir oral 150 mg twice a day for 5 days Placebo |
| Popov 2018 | Peer-reviewed publication | Russia | 100 | 26.50 | 21–60 | 50.00 | 0 | NR | NR | 100 (A) | 100 | NR | 0 | Oseltamivir oral 75 mg twice a day for 5 days Umifenovir 200 mg four times a day for 5 days |
| Puhakka 2003, NAI30015 | Peer-reviewed publication | Finland | 588 | 19.25 | 17–29 | 99.15 | 0 | NR | NR | 73.30 (A), 0.68 (B) | 73.98 | 1.00 | 0.51 | Zanamivir inhaled 10 mg twice a day for 5 days Placebo |
| Roberts 2019 | Peer-reviewed publication | Australia, South Africa, USA | 14 | 34.85 | 18–64 | 78.57 | 0 | NR | NR | 50 (A), 50 (B) | 100 | NR | 0 | Oseltamivir oral 75 mg twice a day for 5 days Placebo |
| Sugaya 2010 | Peer-reviewed publication | Japan | 184 | 6.80 | ≤ 9 | 55.43 | 0 | NR | NR | 60.87 (A/H1N1), 21.20 (A/H3N2), 15.76 (B) | 97.83 | 0.77 | 46.74 | Laninamivir inhaled a single dose of 40 mg on day 1 Laninamivir inhaled a single dose of 20 mg on day 1 Oseltamivir oral twice a day for 5 days. 2 mg/kg of body weight for body weight < 37.5 kg or 75 mg for body weight ≥ 37.5 kg |
| Treanor 2000, WV15671 | Peer-reviewed publication | USA | 627 | 32.63 | 18–65 | 49.28 | 0 | 0 | NR | 54.70 (A), 1.44 (B), 3.51 (unknown) | 59.65 | 1.07 | 0 | Oseltamivir oral 75 mg twice a day for 5 days Oseltamivir oral 150 mg twice a day for 5 days Placebo |
| Wang 2004 | Peer-reviewed publication | China | 232 | 32.40 | 18–65 | 47.20 | NR | 0 | NR | NR | 100 | NR | 0 | Umifenovir 200 mg three times a day for 5 days Placebo |
| Watanabe 2010 | Peer-reviewed publication | Japan, China, Korea | 996 | 35.06 | ≥ 20 | 52.11 | 0 | NR | NR | 64.76 (A/H1N1), 32.33 (A/H3N2), 0.30 (B), 2.61 (negative) | 97.39 | 1.00 | 18.17 | Laninamivir inhaled a single dose of 40 mg on day 1 Laninamivir inhaled a single dose of 20 mg on day 1 Oseltamivir oral 75 mg twice a day for 5 days |
| Watanabe 2013 | Peer-reviewed publication | Japan | 201 | 41.56 | ≥ 20 | 44.28 | 0 | 100 | 97.51 (asthma), 2.49 (COPD), 1.99 (cardiovascular disease), 4.98 (diabetes) | 84.08 (A/H1N1 2009), 10.95 (A/H3N2), 0.50 (A/H1N1 2009+H3N2), 4.48 (negative) | 95.52 | 0.95 | 34.83 | Laninamivir inhaled a single dose of 40 mg on day 1 Oseltamivir oral 75 mg twice a day for 5 days |
| Whitley 2001 | Peer-reviewed publication | USA, Canada | 695 | 5.00 | 1–12 | 50.36 | 0 | 0 | NR | 43.60 (A), 21.29 (B), 0.14 (A+B) | 65.04 | 1.41 | 3.00 | Oseltamivir oral 2 mg/kg/dose twice daily for 5 days Placebo |
| Hayden 2000, NAI30010 | Peer-reviewed publication | USA, Canada, UK, Finland | 321 | 19.46 | ≥ 5 | 42.37 | 0 | NR | 6.54 (underlying respiratory conditions) | 32.09 (A), 16.82 (B) | 48.91 | NR | 10.28 | Zanamivir inhaled 10 mg twice a day for 5 days Placebo |
| NCT00419263 2007 | Trial registration | USA, Canada | 344 | 35.43 | ≥ 18 | 46.49 | 0 | 0 | NR | NR | 100 | NR | 0 | Peramivir single dose administered as bilateral 2-mL intramuscular injections in each gluteal muscle, one injection of peramivir 150 mg Peramivir single dose administered as bilateral 2-mL intramuscular injections in each gluteal muscle, 2 injections of peramivir 150 mg Placebo |
| NCT01068912 2010 | Trial registration | USA, Australia, Chile, New Zealand, Peru, South Africa | 528 | 42.54 | 20–80 | 45.17 | NR | NR | NR | 60.62 (A), 22.78 (B), 2.51 (both) | 100 | NR | NR | Favipiravir 1000 mg twice a day for 1 day, followed by 400 mg twice a day for 4 days Favipiravir 1200 mg twice a day for 1 day, followed by 800 mg twice a day for 4 days Placebo |
| NCT01793883 2013 | Trial registration | Hungary, USA, UK, New Zealand, Canada, Latvia, Belgium, Mexico, South Africa, Bulgaria, Germany, Estonia | 639 | 39.33 | 18–64 | 43.35 | NR | NR | NR | NR | 36.26 | NR | NR | Laninamivir inhaled a single dose of 40 mg on day 1 Laninamivir inhaled a single dose of 80 mg on day 1 Placebo |
| NCT02369159 2015 | Trial registration | USA, South Africa | 137 | 8.32 | 0–17 | 46.72 | 0 | 0 | No | NR | 70.80 | NR | NR | Oseltamivir oral twice a day for 5 days. Subjects ≥ 13 years - 75 mg dose as a capsule or oral suspension. Subjects < 13 years - weight-based dose as a capsule or oral suspension Peramivir a single short intravenous infusion. Subjects ≥12 years - 600 mg. Subjects <12 years - 12 mg/kg (max. 600 mg). Subjects < 6 months - 8 mg/kg |
| NCT00705406 2009 | Trial registration | USA, Australia, New Zealand, South Africa | 405 | 35.00 | ≥ 18 | 49.14 | 0 | 0 | No | NR | 99.26 | NR | 0 | Peramivir 600 mg administered as bilateral 2-mL intramuscular injection Placebo |
| NCT00610935 2008 | Trial registration | USA | 82 | 33.00 | 18–75 | 48.78 | 0 | 0 | No | 25.61 (A/H1N1), 58.54 (A/H3N2), 1.22 (A (not determined), 12.20 (B) | 97.56 | NR | 0 | Peramivir single intramuscular injection of 300 mg Placebo |
| JNAI-01 2000 | Clinical Study Report | Japan | 116 | NR | 16–65 | NR | NR | NR | NR | NR | 48.28 | NR | NR | Zanamivir inhaled 10 mg twice a day for 5 days Zanamivir inhaled 10 mg and intranasal 6.4 mg twice daily for 5 days Placebo |
| JNAI-04 1999 | Clinical Study Report | Japan | 49 | 38.38 | ≥ 16 | 52.08 | 0 | NR | NR | 39.58 (A/H1N1), 4.17 (A/H3N2), 2.08 (A/-) | 45.83 | NR | 0 | Zanamivir inhaled 10 mg twice a day for 5 days Zanamivir inhaled 20 mg twice a day for 5 days Placebo |
| JNAI-07 1999 | Clinical Study Report | Japan | 329 | 34.17 | ≥ 16 | 50.00 | 0 | NR | NR | 0.97 (A), 0.32 (A/H1N1), 69.48 (A/H3N2), 2.27 (B) | 73.04 | NR | 1.30 | Zanamivir inhaled 10 mg twice a day for 5 days Zanamivir inhaled 20 mg twice a day for 5 days Placebo |
| M76001 2000 | Clinical Study Report | USA | 1447 | 35.00 | 13–80 | 44.16 | 0 | 3.87 | 3.73 (asthma), 0.14 (COPD), 1.24 (diabetes), 7.05 (hypertension) | 52.52 (A), 0.28 (A/H1N1), 7.05 (A/H3N2), 13.61 (B) | 73.46 | NR | NR | Oseltamivir oral 75 mg twice a day for 5 days Placebo |
| NAI30011 2001 | Clinical Study Report | USA | 466 | 36.15 | 18–99 | 43.35 | 0 | NR | 7.08 (respiratory diseases), 10.09 (cardiovascular disease) | 39.27 (A), 5.36 (B) | 45.28 | 1.18 | 9.01 | Zanamivir inhaled 10 mg twice a day for 5 days Placebo |
| NAI30012 2002 | Clinical Study Report | Australia, Canada, Chile, Czech Republic, Denmark, Finland, France, Germany, Israel, Italy, Lithuania, Netherlands, Norway, Romania, Russia, Slovakia, South Africa, Spain, UK, USA | 358 | 73.35 | > 65 (almost) | 41.06 | 0 | 100 | 8.66 (chronic respiratory condition), 34.64 (cardiovascular disease), 12.01 (diabetes) | 62.84 (A), 2.23 (B) | 57.54 | NR | 45.81 | Zanamivir inhaled 10 mg twice a day for 5 days Placebo |
| NAI30020 2002 | Clinical Study Report | Germany | 331 | 50.01 | ≥ 18 | 49.30 | 0 | 100 | 43.01 (COPD and asthma), 44.06 (cardiovascular disease) | 100 (A or B) | 100 | NR | NR | Zanamivir inhaled 10 mg twice a day for 5 days Placebo |
| NAI30028 2003 | Clinical Study Report | Germany | 266 | 7.00 | 5–14 | 61.28 | 0 | NR | 12.41 (respiratory diseases), 2.63 (cardiovascular disease) | 100 (A or B) | 100 | NR | NR | Zanamivir inhaled 10 mg twice a day for 5 days Placebo |
| NAIA2005 1998, Hayden 1997 | Clinical Study Report | Canada, USA | 220 | 32.00 | ≥ 13 | 60.91 | 0 | 0 | NR | 37.27 (A), 13.18 (B) | 50.45 | 1.26 | 0 | Zanamivir inhaled 5 mg twice a day for 5 days Zanamivir inhaled 5 mg and spray 0.1 ml twice a day for 5 days Placebo |
| NAIA3002 1998 | Clinical Study Report | Canada, USA | 777 | 35.00 | 12–86 | 48.13 | 0 | 14.03 | 7.33 (respiratory diseases), 3.22 (cardiovascular disease) | 71.81 (A), 1.03 (B) | 73.23 | NR | 14.00 | Zanamivir inhaled 10 mg twice a day for 5 days Placebo |
| NAIB2005 1998, Hayden 1997 | Clinical Study Report | Belgium, Finland, France, Germany, Italy, Netherlands, Norway, Spain, Sweden, UK | 197 | 33.67 | ≥ 18 | 52.55 | 0 | 0 | 5.1 (respiratory diseases), 6.12 (cardiovascular disease) | 32.99 (A), 43.65 (B) | 79.70 | 1.29 | 0 | Zanamivir inhaled 5 mg twice a day for 5 days Zanamivir inhaled 5 mg and spray 0.1 ml twice a day for 5 days Placebo |
| NAIB2007 1998 | Clinical Study Report | Australia, New Zealand, South Africa | 554 | 30.00 | ≥ 13 | 52.00 | 0 | 11.91 | 8.3 (respiratory diseases), 1.81 (cardiovascular disease), 1.44 (diabetes) | 92 (A), 6 (B), 2 (unknow) | 100 | 1.23 | 1.62 | Zanamivir inhaled 5 mg twice a day for 5 days Zanamivir inhaled 5 mg and spray 0.1 ml twice a day for 5 days Placebo |
| WV15707 1999 | Clinical Study Report | Australia, South Africa, South America | 27 | 71.62 | 65–84 | 53.85 | 0 | 100 | 15.38 (chronic obstructive airways disease), 19.23 (cardiac disorders), 7.69 (diabetes), 30.77 (hypertension) | 42.30 (A-H3N2), 3.85 (A/non-specific) | 46.15 | NR | 65.38 | Oseltamivir oral 75 mg twice a day for 5 days Placebo |
| WV15730 1999 | Clinical Study Report | Australia, South Africa | 58 | 35.17 | 18–65 | 51.72 | 0 | NR | 1.72 (bronchospasm), 1.72 (bronchitis), 5.17 (hypertension) | 5.17 (A/H1N1), 56.90 (A/H3N2), 3.44 (A/non-specific) | 65.51 | NR | 0 | Oseltamivir oral 75 mg twice a day for 5 days Placebo |
| WV15758 2000 | Clinical Study Report | USA, Canada | 695 | 5.34 | 1–12 | 50.36 | 0 | NR | 0.57 (pneumonia), 0.43 (bronchitis), 2.30 (asthma) | 43.41 (A), 21.2 (B), 0.14 (A and B) | 64.76 | NR | 3.02 | Oseltamivir oral 2.0 mg/kg (to a maximum of 100 mg per dose) twice a day for 5 days Placebo |
| WV15812/WV15872 2000 | Clinical Study Report | Belgium, Canada, Denmark, Finland, France, Israel, New Zealand, Norway, Sweden, UK, USA, Australia, Netherlands, Russia | 401 | 51.82 | 13–88 | 44.14 | 0 | 100 | 75.81 (COPD), 75.81 (chronic obstructive airways disease), 46.38 (asthma), 17.96 (bronchitis), 5.99 (emphysema), 40.89 (cardiovascular disease), 11.22 (diabetes), 26.93 (hypertension) | 51.00 (A), 11.44 (B) | 62.44 | NR | 27.68 | Oseltamivir oral 75 mg twice a day for 5 days Placebo |
| WV15819/WV15876/WV15978 2000 | Clinical Study Report | France, Netherlands, Belgium, Germany, Switzerland, UK, Norway, Sweden, Denmark, Israel, USA, Canada, South Africa, New Zealand, Australia, Finland, Lithuania, Estonia, Poland | 736 | 72.95 | 65–97 | 42.99 | 0 | 100 | 8.43 (chronic obstructive airways disease), 2.04 (asthma), 23.13 (cardiovascular disease), 11.43 (diabetes), 35.78 (hypertension) | 61.22 (A), 3.54 (B) | 64.67 | NR | 42.86 | Oseltamivir oral 75 mg twice a day for 5 days Placebo |
| WV16277 2003 | Clinical Study Report | Belgium, Denmark, Estonia, Finland, France, Germany, Lithuania, Norway, Sweden | 454 | 34.92 | 13–86 | 49.67 | 0 | NR | 2.89 (chronic obstructive airways disease), 1.33 (asthma), 2.67 (cardiovascular disease), 1.78 (diabetes), 6.65 (hypertension) | 45.45 (A), 4.66 (B), 0.44 (multiple) | 51.00 | NR | 6.87 | Oseltamivir oral 75 mg twice a day for 5 days Placebo |
| NV16871 2004 | Clinical Study Report | Czech Republic, Estonia, Germany, Israel, Latvia, Poland, Russia, Slovakia, Sweden, Ukraine | 329 | 11.00 | 6–17 | 65.35 | 0 | 100 | 100 (asthma) | 24.01 (A), 4.56 (B) | 28.57 | 0.81 | 6.08 | Oseltamivir oral twice a day for 5 days. Body weight > 15 kg – 23 kg: 45 mg; > 23 kg – 40 kg: 60 mg; > 40 kg: 75 mg Placebo |

NR, not reported.

### Appendix 5. Risk of bias for eligible studies

| **Study** | **Sequence generation** | **Allocation concealment** | **Blinding of patients** | **Blinding of health care providers** | **Blinding of** **data collectors** | **Blinding of outcome assessors/**  **adjudicators** | **Blinding of data analysts** | **Incomplete outcome data** | **Selective outcome reporting** | **Other bias** |
| --- | --- | --- | --- | --- | --- | --- | --- | --- | --- | --- |
| **Mortality** | | | | | | | | | | |
| MIST 1998, NAIB3001 | Low | Low | Low | Low | Low | Low | Low | Low | Probably Low | Low |
| Bai 2023 | Low | Probably High | Low | Low | Low | Low | Low | Low | Low | Low |
| Baker 2020 | Probably Low | Probably High | Low | Low | Low | Low | Low | Low | Low | Low |
| Beigel 2020 | Low | Low | Low | Low | Low | Low | Low | Low | Low | Low |
| Butler 2020, Li 2021 (a) | Low | Low | Low | Low | Low | Low | Low | Low | Low | Low |
| Hayden 2018 (b), Portsmouth 2021 | Low | Low | Low | Low | Low | Low | Low | Low | Low | Low |
| Hayden 2022 (a) | Probably High | Probably High | Low | Low | Low | Low | Low | Low | Probably Low | Low |
| Hayden 2022 (b) | Probably High | Probably High | Low | Low | Low | Low | Low | Low | Probably Low | Low |
| Hedrick 2000, NAI30009 | Low | Probably Low | Low | Low | Low | Low | Low | Low | Probably Low | Low |
| Hsieh 2019 | Low | Probably Low | Low | Low | Low | Low | Low | Low | Low | Low |
| Ison 2020, Takazono 2021 | Low | Low | Low | Low | Low | Low | Low | Low | Low | Low |
| Johnston 2005, WV15759/WV15871 | Low | Low | Low | Low | Low | Low | Low | Low | Low | Low |
| Kashiwagi 2000, JV15823 | Low | Probably Low | Low | Low | Low | Low | Low | Low | Probably Low | Low |
| Makela 2000, NAIB3002 | Low | Low | Low | Low | Low | Low | Probably Low | Low | Probably Low | Low |
| Monto 1999, NAIA/B2008 | Low | Probably Low | Low | Low | Low | Low | Low | Low | Low | Low |
| Murphy 2000, NAI30008 | Low | Probably Low | Low | Low | Low | Low | Low | Low | Low | Low |
| Nicholson 2000, WV15670 | Low | Low | Low | Low | Low | Low | Low | Low | Low | Low |
| Puhakka 2003, NAI30015 | Low | Probably Low | Low | Low | Low | Low | Low | Low | Low | Low |
| Roberts 2019 | Probably High | Probably High | Low | Low | Low | Low | Low | Low | Low | Low |
| Treanor 2000, WV15671 | Low | Low | Low | Low | Low | Low | Low | Low | Low | Low |
| NCT01793883 2013 | Probably High | Probably High | Low | Low | Low | Low | Low | Low | Probably Low | Low |
| NCT02369159 2015 | Probably High | Probably High | Low | Low | Low | Low | Low | Low | Probably Low | Low |
| Hayden 2000, NAI30010 | Probably Low | Probably Low | Low | Low | Low | Low | Low | Low | Low | Low |
| NCT00610935 2008 | Probably High | Probably High | Low | Low | Low | Low | Low | Low | Probably Low | Probably High |
| JNAI-04 1999 | Probably High | Low | Low | Low | Low | Low | Low | Low | Low | Probably Low |
| JNAI-07 1999 | Probably High | Low | Low | Low | Low | Low | Low | Low | Low | High |
| M76001 2000 | Low | Low | Low | Low | Low | Low | Low | Low | Low | Low |
| NAI30011 2001 | Low | Low | Low | Low | Low | Low | Low | Low | Low | Low |
| NAI30012 2002 | Probably Low | Low | Low | Low | Low | Low | Low | Low | Low | Low |
| NAI30028 2003 | Low | Low | Low | Low | Low | Low | Low | Low | Probably Low | Low |
| NAIA2005 1998, Hayden 1997 | Low | Low | Low | Low | Low | Low | Low | Low | Low | Low |
| NAIA3002 1998 | Low | Low | Low | Low | Low | Low | Low | Low | Low | Low |
| NAIB2005 1998, Hayden 1997 | Low | Low | Low | Low | Low | Low | Low | Low | Low | Low |
| NAIB2007 1998 | Low | Low | Low | Low | Low | Low | Low | Low | Low | Low |
| WV15707 1999 | Low | Low | Low | Low | Low | Low | Low | Low | Low | Probably High |
| WV15730 1999 | Low | Low | Low | Low | Low | Low | Low | Low | Low | Probably High |
| WV15758 2000 | Low | Low | Low | Low | Low | Low | Low | Low | Low | Low |
| WV15812/WV15872 2000 | Low | Low | Low | Low | Low | Low | Low | Low | Low | Probably High |
| WV15819/WV15876/WV15978 2000 | Low | Low | Low | Low | Low | Low | Low | Low | Low | Probably Low |
| WV16277 2003 | Low | Low | Low | Low | Low | Low | Low | Low | Low | Probably High |
| NV16871 2004 | Low | Low | Low | Low | Low | Low | Low | Low | Low | Low |
| **Admission to hospital** | | | | | | | | | | |
| Bai 2023 | Low | Probably High | Low | Low | Low | Low | Low | Low | Low | Low |
| Baker 2020 | Probably Low | Probably High | Low | Low | Low | Low | Low | Low | Low | Low |
| Beigel 2020 | Low | Low | Low | Low | Low | Low | Low | Low | Low | Low |
| Butler 2020, Li 2021 (a) | Low | Low | Low | Low | Low | Low | Low | Low | Low | Low |
| Dharan 2012 | Probably High | Probably High | Low | Low | Low | Low | Low | Low | Probably Low | Low |
| Fry 2014 | Low | Low | Low | Low | Low | Low | Low | Low | Probably Low | Low |
| Hayden 2018 (b), Portsmouth 2021 | Low | Low | Low | Low | Low | Low | Low | Low | Low | Low |
| Hayden 2022 (a) | Probably High | Probably High | Low | Low | Low | Low | Low | Low | Probably Low | Low |
| Hayden 2022 (b) | Probably High | Probably High | Low | Low | Low | Low | Low | Low | Probably Low | Low |
| Heinonen 2010 | Low | Probably Low | Low | Low | Low | Low | Low | Low | Low | Low |
| Ison 2020, Takazono 2021 | Low | Low | Low | Low | Low | Low | Low | Low | Low | Low |
| Johnston 2005, WV15759/WV15871 | Low | Low | Low | Low | Low | Low | Low | Low | Low | Low |
| Katsumi 2012 | Probably High | Probably High | Low | Low | Low | Low | Low | High | Probably Low | Low |
| Lin 2006 | Probably Low | Probably High | Low | Low | Low | Low | Low | Low | Probably Low | Low |
| Murphy 2000, NAI30008 | Low | Probably Low | Low | Low | Low | Low | Low | Low | Low | Low |
| Nicholson 2000, WV15670 | Low | Low | Low | Low | Low | Low | Low | Low | Low | Low |
| Puhakka 2003, NAI30015 | Low | Probably Low | Low | Low | Low | Low | Low | Low | Low | Low |
| Roberts 2019 | Probably High | Probably High | Low | Low | Low | Low | Low | Low | Low | Low |
| Treanor 2000, WV15671 | Low | Low | Low | Low | Low | Low | Low | Low | Low | Low |
| Whitley 2001 | Probably Low | Probably High | Low | Low | Low | Low | Low | Low | Probably Low | Low |
| JNAI-04 1999 | Probably High | Low | Low | Low | Low | Low | Low | Low | Low | Probably Low |
| M76001 2000 | Low | Low | Low | Low | Low | Low | Low | Low | Low | Low |
| WV15707 1999 | Low | Low | Low | Low | Low | Low | Low | Low | Low | Probably High |
| WV15758 2000 | Low | Low | Low | Low | Low | Low | Low | Low | Low | Low |
| WV15812/WV15872 2000 | Low | Low | Low | Low | Low | Low | Low | Low | Low | Probably High |
| WV15819/WV15876/WV15978 2000 | Low | Low | Low | Low | Low | Low | Low | Low | Low | Probably Low |
| WV16277 2003 | Low | Low | Low | Low | Low | Low | Low | Low | Low | Probably High |
| NV16871 2004 | Low | Low | Low | Low | Low | Low | Low | Low | Low | Low |
| **Admission to ICU** | | | | | | | | | | |
| Hsieh 2019 | Low | Probably Low | Low | Low | Low | Low | Low | Low | Low | Low |
| M76001 2000 | Low | Low | Low | Low | Low | Low | Low | Low | Low | Low |
| **Emergence of resistance** | | | | | | | | | | |
| Baker 2020 | Probably Low | Probably High | Low | Low | Low | Low | Low | Low | Low | Low |
| Escuret 2012 | Low | Low | Low | Low | Low | Low | Low | Low | Probably Low | Low |
| Fry 2014 | Low | Low | Low | Low | Low | Low | Low | Low | Probably Low | Low |
| Hayden 2018 (b), Portsmouth 2021 | Low | Low | Low | Low | Low | Low | Low | Low | Low | Low |
| Hedrick 2000, NAI30009 | Low | Probably Low | Low | Low | Low | Low | Low | Low | Probably Low | Low |
| Ison 2020, Takazono 2021 | Low | Low | Low | Low | Low | Low | Low | Low | Probably Low | Low |
| Nakamura 2017 | Low | Probably Low | Low | Low | Low | Low | Low | Low | Probably Low | Probably High |
| Roberts 2019 | Probably High | Probably High | Low | Low | Low | Low | Low | Low | Low | Low |
| Whitley 2001 | Probably Low | Probably High | Low | Low | Low | Low | Low | Low | Probably Low | Low |
| NAI30012 2002 | Probably Low | Low | Low | Low | Low | Low | Low | Low | Low | Low |
| WV15730 1999 | Low | Low | Low | Low | Low | Low | Low | Low | Low | Probably High |
| WV15812/WV15872 2000 | Low | Low | Low | Low | Low | Low | Low | Low | Low | Probably High |
| **Time to alleviation of symptoms** | | | | | | | | | | |
| MIST 1998, NAIB3001 | Low | Low | Low | Low | Probably High | Probably High | Probably High | Low | Probably Low | Low |
| Bai 2023 | Low | Probably High | High | High | High | High | High | Low | Low | Low |
| Baker 2020 | Probably Low | Probably High | Low | Low | Probably High | Probably High | Probably High | Low | Low | Low |
| Beigel 2020 | Low | Low | Low | Low | Low | Low | Low | Low | Low | Low |
| Dharan 2012 | Probably High | Probably High | Probably High | Probably High | Probably High | Probably High | Probably High | Low | Probably Low | Low |
| Duval 2010 | Low | Low | Low | Low | Low | Low | Low | Low | Low | Probably High |
| Fan 2019 | Low | Low | Low | Low | Probably High | Probably High | Probably High | Low | Probably Low | Low |
| Fry 2014 | Low | Low | Low | Low | Low | Low | Probably High | Low | Low | Low |
| Galbraith 1971 | Probably Low | Probably High | Low | Low | Probably High | Probably High | Probably High | High | Probably Low | Low |
| Galbraith 1973 | Low | Probably High | Low | Low | Low | Probably High | Probably High | Low | Probably Low | Low |
| Hayden 2018 (a), Watanabe 2019 | Low | Low | Low | Low | Low | Low | Low | Low | Low | Low |
| Hayden 2018 (b), Portsmouth 2021 | Low | Low | Low | Low | Low | Low | Low | Low | Probably Low | Low |
| Hayden 2022 (a) | Probably High | Probably High | Low | Low | Low | Low | Probably High | Low | Low | Low |
| Hayden 2022 (b) | Probably High | Probably High | Low | Low | Low | Low | Probably High | Low | Low | Low |
| Hedrick 2000, NAI30009 | Low | Probably Low | Low | Low | Probably High | Probably High | Probably High | Low | Probably Low | Low |
| Heinonen 2010 | Low | Probably Low | Low | Low | Probably High | Probably High | Probably High | Low | Low | Low |
| Hirotsu 2017 | Probably Low | Low | High | High | High | High | High | Low | Low | Probably Low |
| Ison 2020, Takazono 2021 | Low | Low | Low | Low | Low | Low | Low | Low | Low | Low |
| Johnston 2005, WV15759/WV15871 | Low | Low | Low | Low | Low | Low | Low | Low | Low | Low |
| Kashiwagi 2000, JV15823 | Low | Probably Low | Low | Low | Probably High | Probably High | Probably High | Low | Probably Low | Low |
| Kato 2021 | Probably Low | Probably High | High | High | High | High | High | Low | Low | Low |
| Kohno 2010 | Low | Low | Low | Low | Probably High | Probably High | Probably High | Low | Probably Low | Low |
| Kohno 2011 | Probably Low | Probably High | Low | Low | Probably High | Probably High | Probably High | Low | Probably Low | Low |
| Li 2004 | Probably Low | Probably Low | Low | Low | Probably High | Probably High | Probably High | Low | Probably Low | Low |
| Lin 2006 | Probably Low | Probably High | High | High | High | High | High | Low | Probably Low | Low |
| Makela 2000, NAIB3002 | Low | Low | Low | Low | Low | Low | Probably Low | Low | Probably Low | Low |
| McLean 2015 | Low | Probably High | Low | Low | Probably High | Probably High | Probably High | Low | Probably Low | Low |
| Monto 1999, NAIA/B2008 | Low | Probably Low | Low | Low | Probably High | Low | Probably High | Low | Low | Low |
| Murphy 2000, NAI30008 | Low | Probably Low | Low | Low | Probably High | Probably High | Probably High | Low | Probably Low | Low |
| Nakamura 2017 | Low | Probably Low | High | High | High | High | High | Low | Probably Low | Probably High |
| Nicholson 2000, WV15670 | Low | Low | Low | Low | Low | Low | Low | Low | Low | Low |
| Puhakka 2003, NAI30015 | Low | Probably Low | Low | Low | Probably High | Low | Probably High | Low | Low | Low |
| Sugaya 2010 | Low | Probably Low | Low | Low | Probably High | Probably High | Probably High | Low | Low | Low |
| Treanor 2000, WV15671 | Low | Low | Low | Low | Low | Low | Low | Low | Low | Low |
| Wang 2004 | Low | Probably Low | Low | Low | Probably High | Probably High | Probably High | Low | Probably Low | Low |
| Watanabe 2010 | Low | Probably Low | Low | Low | Probably High | Probably High | Probably High | Low | Low | Low |
| Watanabe 2013 | Low | Probably Low | Low | Low | Probably High | Probably High | Probably High | Low | Probably Low | Low |
| NCT01793883 2013 | Probably High | Probably High | Low | Low | Probably High | Low | Probably High | Low | Low | Low |
| Hayden 2000, NAI30010 | Probably Low | Probably Low | Low | Low | Probably High | Probably High | Probably High | Low | Low | Low |
| NCT00705406 2009 | Probably High | Probably High | Low | Low | Probably High | Probably High | Probably High | Low | Low | Low |
| NCT00610935 2008 | Probably High | Probably High | Low | Low | Probably High | Probably High | Probably High | Low | Low | Probably High |
| JNAI-04 1999 | Probably High | Low | High | High | High | High | High | Low | Low | Probably Low |
| JNAI-07 1999 | Probably High | Low | High | High | High | High | High | Low | Low | High |
| M76001 2000 | Low | Low | Low | Low | Low | Low | Low | Low | Low | Low |
| NAI30011 2001 | Low | Low | Low | Low | Probably High | Low | Probably High | Low | Low | Low |
| NAI30012 2002 | Probably Low | Low | Low | Low | Probably High | Low | Probably High | Low | Low | Low |
| NAI30028 2003 | Low | Low | Low | Low | Probably High | Low | Probably High | Low | Probably Low | Low |
| NAIA2005 1998, Hayden 1997 | Low | Low | Low | Low | Probably High | Probably High | Probably High | Low | Low | Low |
| NAIA3002 1998 | Low | Low | Low | Low | Probably High | Probably High | Probably High | Low | Low | Low |
| NAIB2005 1998, Hayden 1997 | Low | Low | Low | Low | Probably High | Probably High | Probably High | Low | Low | Low |
| WV15707 1999 | Low | Low | Low | Low | Probably High | Low | Low | Low | Low | Probably High |
| WV15730 1999 | Low | Low | Low | Low | Probably High | Low | Low | Low | Low | Probably High |
| WV15758 2000 | Low | Low | Low | Low | Probably High | Low | Low | Low | Low | Low |
| WV15812/WV15872 2000 | Low | Low | Low | Low | Probably High | Low | Low | Low | Low | Probably High |
| WV15819/WV15876/WV15978 2000 | Low | Low | Low | Low | Probably High | Low | Low | Low | Low | Probably Low |
| WV16277 2003 | Low | Low | Low | Low | Probably High | Low | Low | Low | Low | Probably High |
| NV16871 2004 | Low | Low | Low | Low | Low | Low | Low | Low | Low | Low |
| Whitley 2001 | Probably Low | Probably High | Low | Low | Probably High | Probably High | Probably High | Low | Probably Low | Low |
| NCT00419263 2007 | Probably High | Probably High | Low | Low | Low | Low | Probably High | Low | Probably Low | Low |
| **Duration of hospitalization** | | | | | | | | | | |
| Butler 2020, Li 2021 (a) | Low | Low | High | High | High | High | High | Low | Probably Low | Low |
| Li 2021 (b) | Low | Probably High | Probably High | Probably High | Probably High | Probably High | Probably High | Low | Probably Low | Low |
| Puhakka 2003, NAI30015 | Low | Probably Low | Low | Low | Probably High | Low | Probably High | Low | Low | Low |
| **Any adverse events** | | | | | | | | | | |
| MIST 1998, NAIB3001 | Low | Low | Low | Low | Probably High | Probably High | Probably High | Low | Probably Low | Low |
| Bai 2023 | Low | Probably High | High | High | High | High | High | Low | Low | Low |
| Baker 2020 | Probably Low | Probably High | Low | Low | Probably High | Probably High | Probably High | Low | Low | Low |
| Fan 2019 | Low | Low | Low | Low | Probably High | Probably High | Probably High | Low | Probably Low | Low |
| Hayden 2018 (a), Watanabe 2019 | Low | Low | Low | Low | Low | Low | Low | Low | Low | Low |
| Hayden 2018 (b), Portsmouth 2021 | Low | Low | Low | Low | Low | Low | Low | Low | Low | Low |
| Hayden 2022 (a) | Probably High | Probably High | Low | Low | Low | Low | Probably High | Low | Low | Low |
| Hayden 2022 (b) | Probably High | Probably High | Low | Low | Low | Low | Probably High | Low | Low | Low |
| Hedrick 2000, NAI30009 | Low | Probably Low | Low | Low | Probably High | Probably High | Probably High | Low | Probably Low | Low |
| Hirotsu 2017 | Probably Low | Low | High | High | High | High | High | Low | Low | Probably Low |
| Ison 2020, Takazono 2021 | Low | Low | Low | Low | Low | Low | Low | Low | Low | Low |
| Johnston 2005, WV15759/WV15871 | Low | Low | Low | Low | Low | Low | Low | Low | Low | Low |
| Kashiwagi 2000, JV15823 | Low | Probably Low | Low | Low | Probably High | Probably High | Probably High | Low | Probably Low | Low |
| Kato 2021 | Probably Low | Probably High | High | High | High | High | High | Low | Low | Low |
| Kiselev 2015 | Probably Low | Probably High | Low | Low | Probably High | Probably High | Probably High | Low | Probably Low | Low |
| Kohno 2010 | Low | Low | Low | Low | Probably High | Probably High | Probably High | Low | Probably Low | Low |
| Kohno 2011 | Probably Low | Probably High | Low | Low | Probably High | Probably High | Probably High | Low | Probably Low | Low |
| Lin 2006 | Probably Low | Probably High | High | High | High | High | High | Low | Probably Low | Low |
| Li 2021 (b) | Low | Probably High | Probably High | Probably High | Probably High | Probably High | Probably High | Low | Probably Low | Low |
| Makela 2000, NAIB3002 | Low | Low | Low | Low | Low | Low | Probably Low | Low | Probably Low | Low |
| Matsumoto 1999 | Probably High | Probably High | Low | Low | Probably High | Probably High | Probably High | Low | Probably Low | Low |
| Monto 1999, NAIA/B2008 | Low | Probably Low | Low | Low | Probably High | Low | Probably High | Low | Low | Low |
| Murphy 2000, NAI30008 | Low | Probably Low | Low | Low | Probably High | Probably High | Probably High | Low | Low | Low |
| Nakamura 2017 | Low | Probably Low | High | High | High | High | High | Low | Probably Low | Probably High |
| Nicholson 2000, WV15670 | Low | Low | Low | Low | Low | Low | Low | Low | Low | Low |
| Popov 2018 | Probably High | Probably High | High | High | High | High | High | Low | Probably Low | Low |
| Puhakka 2003, NAI30015 | Low | Probably Low | Low | Low | Probably High | Low | Probably High | Low | Low | Low |
| Roberts 2019 | Probably High | Probably High | Low | Low | Probably High | Probably High | Probably High | Low | Low | Low |
| Treanor 2000, WV15671 | Low | Low | Low | Low | Low | Low | Low | Low | Low | Low |
| Hayden 2000, NAI30010 | Probably Low | Probably Low | Low | Low | Probably High | Probably High | Probably High | Low | Low | Low |
| JNAI-01 2000 | Probably High | Probably Low | Low | Low | Probably High | Probably High | Probably High | High | Low | Probably High |
| JNAI-04 1999 | Probably High | Low | High | High | High | High | High | Low | Low | Probably Low |
| JNAI-07 1999 | Probably High | Low | High | High | High | High | High | Low | Low | High |
| M76001 2000 | Low | Low | Low | Low | Low | Low | Low | Low | Low | Low |
| NAI30011 2001 | Low | Low | Low | Low | Probably High | Low | Probably High | Low | Low | Low |
| NAI30012 2002 | Probably Low | Low | Low | Low | Probably High | Low | Probably High | Low | Low | Low |
| NAI30020 2002 | Probably High | Probably High | Low | Low | Probably High | Probably High | Probably High | Low | Probably Low | Low |
| NAI30028 2003 | Low | Low | Low | Low | Probably High | Low | Probably High | Low | Probably Low | Low |
| NAIA2005 1998, Hayden 1997 | Low | Low | Low | Low | Probably High | Probably High | Probably High | Low | Low | Low |
| NAIA3002 1998 | Low | Low | Low | Low | Probably High | Probably High | Probably High | Low | Low | Low |
| NAIB2005 1998, Hayden 1997 | Low | Low | Low | Low | Probably High | Probably High | Probably High | Low | Low | Low |
| NAIB2007 1998 | Low | Low | Low | Low | Probably High | Probably High | Probably High | Low | Low | Low |
| WV15707 1999 | Low | Low | Low | Low | Probably High | Low | Low | Low | Low | Probably High |
| WV15730 1999 | Low | Low | Low | Low | Probably High | Low | Low | Low | Low | Probably High |
| WV15758 2000 | Low | Low | Low | Low | Probably High | Low | Low | Low | Low | Low |
| WV15812/WV15872 2000 | Low | Low | Low | Low | Probably High | Low | Low | Low | Low | Probably High |
| WV15819/WV15876/WV15978 2000 | Low | Low | Low | Low | Probably High | Low | Low | Low | Low | Probably Low |
| WV16277 2003 | Low | Low | Low | Low | Probably High | Low | Low | Low | Low | Probably High |
| NV16871 2004 | Low | Low | Low | Low | Probably High | Low | Low | Low | Low | Low |
| **Adverse events related to treatments** | | | | | | | | | | |
| MIST 1998, NAIB3001 | Low | Low | Low | Low | Probably High | Probably High | Probably High | Low | Probably Low | Low |
| Baker 2020 | Probably Low | Probably High | Low | Low | Probably High | Probably High | Probably High | Low | Low | Low |
| Escuret 2012 | Low | Low | High | High | High | High | High | Low | Probably Low | Low |
| Hayden 2018 (a), Watanabe 2019 | Low | Low | Low | Low | Low | Low | Low | Low | Low | Low |
| Hayden 2018 (b), Portsmouth 2021 | Low | Low | Low | Low | Low | Low | Low | Low | Low | Low |
| Hayden 2022 (a) | Probably High | Probably High | Low | Low | Low | Low | Probably High | Low | Low | Low |
| Hayden 2022 (b) | Probably High | Probably High | Low | Low | Low | Low | Probably High | Low | Low | Low |
| Hedrick 2000, NAI30009 | Low | Probably Low | Low | Low | Probably High | Probably High | Probably High | Low | Probably Low | Low |
| Ison 2020, Takazono 2021 | Low | Low | Low | Low | Low | Low | Low | Low | Low | Low |
| Johnston 2005, WV15759/WV15871 | Low | Low | Low | Low | Low | Low | Low | Low | Low | Low |
| Kohno 2011 | Probably Low | Probably High | Low | Low | Probably High | Probably High | Probably High | Low | Probably Low | Low |
| Li 2004 | Probably Low | Probably Low | Low | Low | Probably High | Probably High | Probably High | Low | Probably Low | Low |
| Monto 1999, NAIA/B2008 | Low | Probably Low | Low | Low | Probably High | Low | Probably High | Low | Low | Low |
| Murphy 2000, NAI30008 | Low | Probably Low | Low | Low | Probably High | Probably High | Probably High | Low | Low | Low |
| Nicholson 2000, WV15670 | Low | Low | Low | Low | Low | Low | Low | Low | Low | Low |
| Puhakka 2003, NAI30015 | Low | Probably Low | Low | Low | Probably High | Low | Probably High | Low | Low | Low |
| Roberts 2019 | Probably High | Probably High | Low | Low | Probably High | Probably High | Probably High | Low | Low | Low |
| Treanor 2000, WV15671 | Low | Low | Low | Low | Low | Low | Low | Low | Low | Low |
| Wang 2004 | Low | Probably Low | Low | Low | Probably High | Probably High | Probably High | Low | Probably Low | Low |
| Hayden 2000, NAI30010 | Probably Low | Probably Low | Low | Low | Probably High | Probably High | Probably High | Low | Low | Low |
| JNAI-01 2000 | Probably High | Probably Low | Low | Low | Probably High | Probably High | Probably High | High | Low | Probably High |
| JNAI-04 1999 | Probably High | Low | High | High | High | High | High | Low | Low | Probably Low |
| JNAI-07 1999 | Probably High | Low | High | High | High | High | High | Low | Low | High |
| M76001 2000 | Low | Low | Low | Low | Low | Low | Low | Low | Low | Low |
| NAI30011 2001 | Low | Low | Low | Low | Probably High | Low | Probably High | Low | Low | Low |
| NAI30012 2002 | Probably Low | Low | Low | Low | Probably High | Low | Probably High | Low | Low | Low |
| NAI30020 2002 | Probably High | Probably High | Low | Low | Probably High | Probably High | Probably High | Low | Probably Low | Low |
| NAI30028 2003 | Low | Low | Low | Low | Probably High | Low | Probably High | Low | Probably Low | Low |
| NAIA2005 1998, Hayden 1997 | Low | Low | Low | Low | Probably High | Probably High | Probably High | Low | Low | Low |
| NAIA3002 1998 | Low | Low | Low | Low | Probably High | Probably High | Probably High | Low | Low | Low |
| NAIB2005 1998, Hayden 1997 | Low | Low | Low | Low | Probably High | Probably High | Probably High | Low | Low | Low |
| NAIB2007 1998 | Low | Low | Low | Low | Probably High | Probably High | Probably High | Low | Low | Low |
| WV15707 1999 | Low | Low | Low | Low | Probably High | Low | Low | Low | Low | Probably High |
| WV15730 1999 | Low | Low | Low | Low | Probably High | Low | Low | Low | Low | Probably High |
| WV15758 2000 | Low | Low | Low | Low | Probably High | Low | Low | Low | Low | Low |
| NV16871 2004 | Low | Low | Low | Low | Low | Low | Low | Low | Low | Low |
| **Serious adverse events** | | | | | | | | | | |
| MIST 1998, NAIB3001 | Low | Low | Low | Low | Probably High | Probably High | Probably High | Low | Probably Low | Low |
| Baker 2020 | Probably Low | Probably High | Low | Low | Probably High | Probably High | Probably High | Low | Low | Low |
| Beigel 2020 | Low | Low | Low | Low | Low | Low | Low | Low | Low | Low |
| Butler 2020, Li 2021 (a) | Low | Low | High | High | High | High | High | Low | Low | Low |
| Duval 2010 | Low | Low | Low | Low | Low | Low | Low | Low | Low | Probably High |
| Fan 2019 | Low | Low | Low | Low | Probably High | Probably High | Probably High | Low | Probably Low | Low |
| Fry 2014 | Low | Low | Low | Low | Low | Low | Probably High | Low | Low | Low |
| Hayden 2018 (a), Watanabe 2019 | Low | Low | Low | Low | Low | Low | Low | Low | Low | Low |
| Hayden 2018 (b), Portsmouth 2021 | Low | Low | Low | Low | Low | Low | Low | Low | Low | Low |
| Hayden 2022 (a) | Probably High | Probably High | Low | Low | Low | Low | Probably High | Low | Low | Low |
| Hayden 2022 (b) | Probably High | Probably High | Low | Low | Low | Low | Probably High | Low | Low | Low |
| Hedrick 2000, NAI30009 | Low | Probably Low | Low | Low | Probably High | Probably High | Probably High | Low | Probably Low | Low |
| Heinonen 2010 | Low | Probably Low | Low | Low | Probably High | Probably High | Probably High | Low | Low | Low |
| Hsieh 2019 | Low | Probably Low | High | High | High | High | High | Low | Low | Low |
| Ison 2020, Takazono 2021 | Low | Low | Low | Low | Low | Low | Low | Low | Low | Low |
| Johnston 2005, WV15759/WV15871 | Low | Low | Low | Low | Low | Low | Low | Low | Low | Low |
| Kato 2021 | Probably Low | Probably High | High | High | High | High | High | Low | Low | Low |
| Kohno 2010 | Low | Low | Low | Low | Probably High | Probably High | Probably High | Low | Probably Low | Low |
| Kohno 2011 | Probably Low | Probably High | Low | Low | Probably High | Probably High | Probably High | Low | Probably Low | Low |
| Li 2004 | Probably Low | Probably Low | Low | Low | Probably High | Probably High | Probably High | Low | Probably Low | Low |
| Makela 2000, NAIB3002 | Low | Low | Low | Low | Low | Low | Probably Low | Low | Probably Low | Low |
| Matsumoto 1999 | Probably High | Probably High | Low | Low | Probably High | Probably High | Probably High | Low | Probably Low | Low |
| McLean 2015 | Low | Probably High | Low | Low | Probably High | Probably High | Probably High | Low | Probably Low | Low |
| Monto 1999, NAIA/B2008 | Low | Probably Low | Low | Low | Probably High | Low | Probably High | Low | Low | Low |
| Murphy 2000, NAI30008 | Low | Probably Low | Low | Low | Probably High | Probably High | Probably High | Low | Low | Low |
| Nicholson 2000, WV15670 | Low | Low | Low | Low | Low | Low | Low | Low | Low | Low |
| Puhakka 2003, NAI30015 | Low | Probably Low | Low | Low | Probably High | Low | Probably High | Low | Low | Low |
| Roberts 2019 | Probably High | Probably High | Low | Low | Probably High | Probably High | Probably High | Low | Low | Low |
| Treanor 2000, WV15671 | Low | Low | Low | Low | Low | Low | Low | Low | Low | Low |
| Watanabe 2010 | Low | Probably Low | Low | Low | Probably High | Probably High | Probably High | Low | Low | Low |
| Watanabe 2013 | Low | Probably Low | Low | Low | Probably High | Probably High | Probably High | Low | Probably Low | Low |
| Whitley 2001 | Probably Low | Probably High | Low | Low | Probably High | Probably High | Probably High | Low | Probably Low | Low |
| NCT00419263 2007 | Probably High | Probably High | Low | Low | Low | Low | Probably High | Low | Low | Low |
| NCT01068912 2010 | Probably High | Probably High | Low | Low | Low | Low | Probably High | Low | Probably Low | Low |
| NCT01793883 2013 | Probably High | Probably High | Low | Low | Probably High | Low | Probably High | Low | Probably Low | Low |
| NCT02369159 2015 | Probably High | Probably High | High | High | High | High | High | Low | Probably Low | Low |
| Hayden 2000, NAI30010 | Probably Low | Probably Low | Low | Low | Probably High | Probably High | Probably High | Low | Low | Low |
| NCT00705406 2009 | Probably High | Probably High | Low | Low | Probably High | Probably High | Probably High | Low | Probably Low | Low |
| NCT00610935 2008 | Probably High | Probably High | Low | Low | Probably High | Probably High | Probably High | Low | Probably Low | Probably High |
| JNAI-01 2000 | Probably High | Probably Low | Low | Low | Probably High | Probably High | Probably High | High | Low | Probably High |
| JNAI-04 1999 | Probably High | Low | High | High | High | High | High | Low | Low | Probably Low |
| JNAI-07 1999 | Probably High | Low | High | High | High | High | High | Low | Low | High |
| M76001 2000 | Low | Low | Low | Low | Low | Low | Low | Low | Low | Low |
| NAI30011 2001 | Low | Low | Low | Low | Probably High | Low | Probably High | Low | Low | Low |
| NAI30012 2002 | Probably Low | Low | Low | Low | Probably High | Low | Probably High | Low | Low | Low |
| NAI30020 2002 | Probably High | Probably High | Low | Low | Probably High | Probably High | Probably High | Low | Probably Low | Low |
| NAI30028 2003 | Low | Low | Low | Low | Probably High | Low | Probably High | Low | Probably Low | Low |
| NAIA2005 1998, Hayden 1997 | Low | Low | Low | Low | Probably High | Probably High | Probably High | Low | Low | Low |
| NAIA3002 1998 | Low | Low | Low | Low | Probably High | Probably High | Probably High | Low | Low | Low |
| NAIB2005 1998, Hayden 1997 | Low | Low | Low | Low | Probably High | Probably High | Probably High | Low | Low | Low |
| NAIB2007 1998 | Low | Low | Low | Low | Probably High | Probably High | Probably High | Low | Low | Low |
| WV15707 1999 | Low | Low | Low | Low | Probably High | Low | Low | Low | Low | Probably High |
| WV15730 1999 | Low | Low | Low | Low | Probably High | Low | Low | Low | Low | Probably High |
| WV15758 2000 | Low | Low | Low | Low | Probably High | Low | Low | Low | Low | Low |
| WV15812/WV15872 2000 | Low | Low | Low | Low | Probably High | Low | Low | Low | Low | Probably High |
| WV15819/WV15876/WV15978 2000 | Low | Low | Low | Low | Probably High | Low | Low | Low | Low | Probably Low |
| WV16277 2003 | Low | Low | Low | Low | Probably High | Low | Low | Low | Low | Probably High |
| NV16871 2004 | Low | Low | Low | Low | Probably High | Low | Low | Low | Low | Low |

### Appendix 6. Network plots

*The size of the circle represents the number of participants. The width of the line represents the number of studies.

#### 6.1. Network plot for admission to hospital

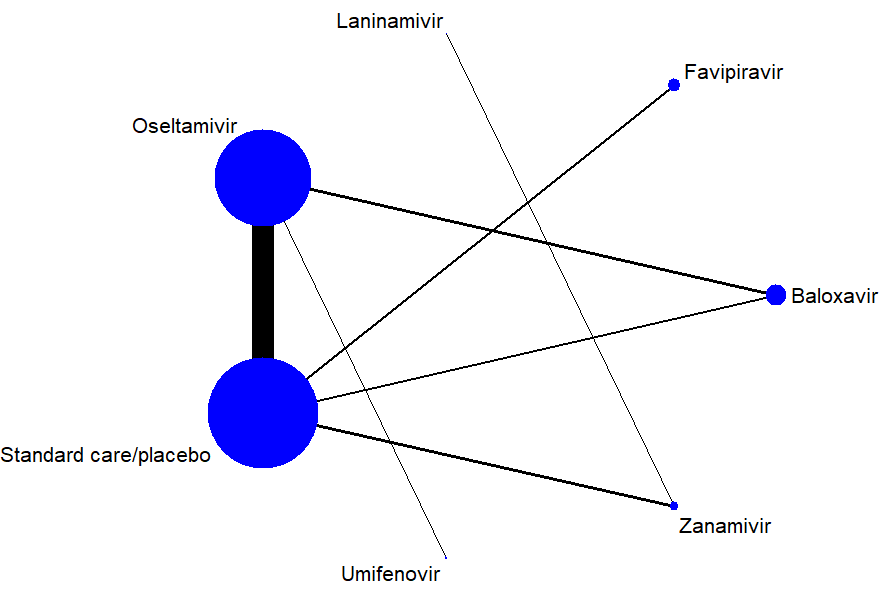

#### 6.2. Network plot for admission to ICU

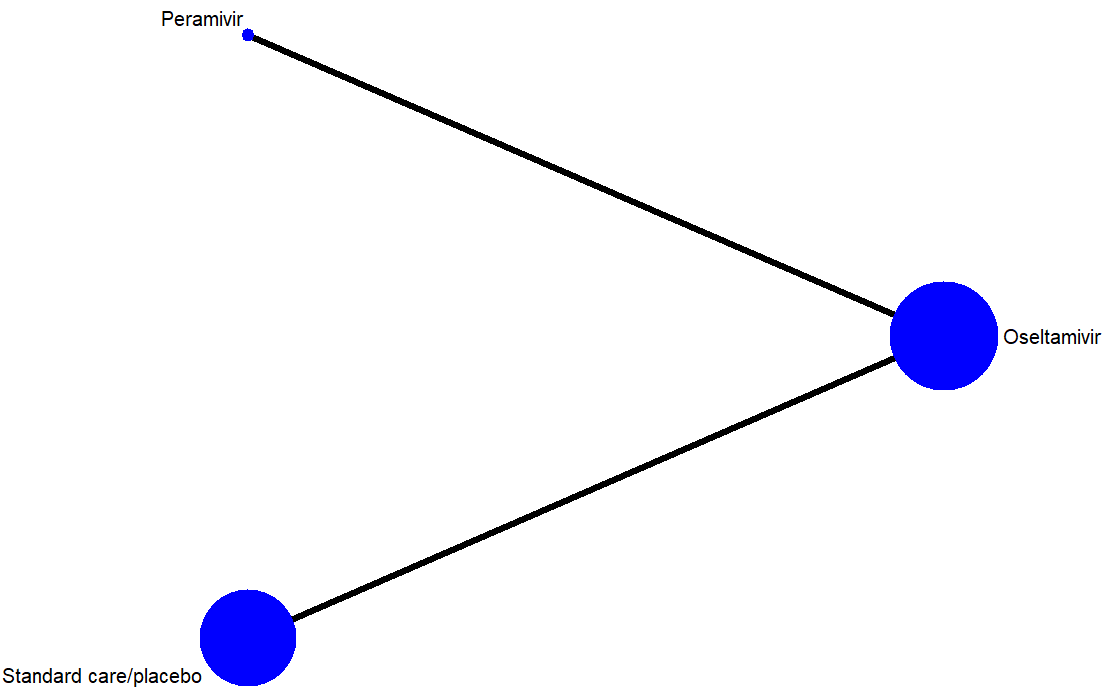

#### 6.3. Network plot for duration of hospitalization

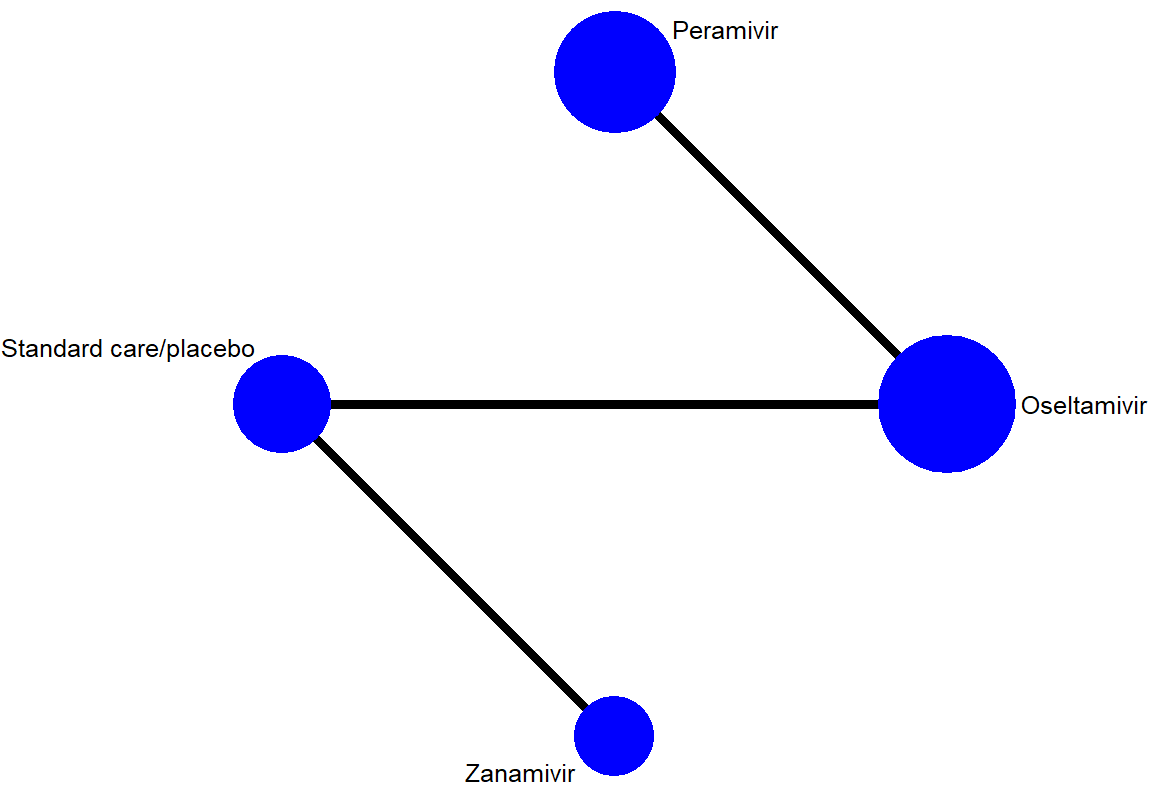

#### 6.4. Network plot for any adverse events

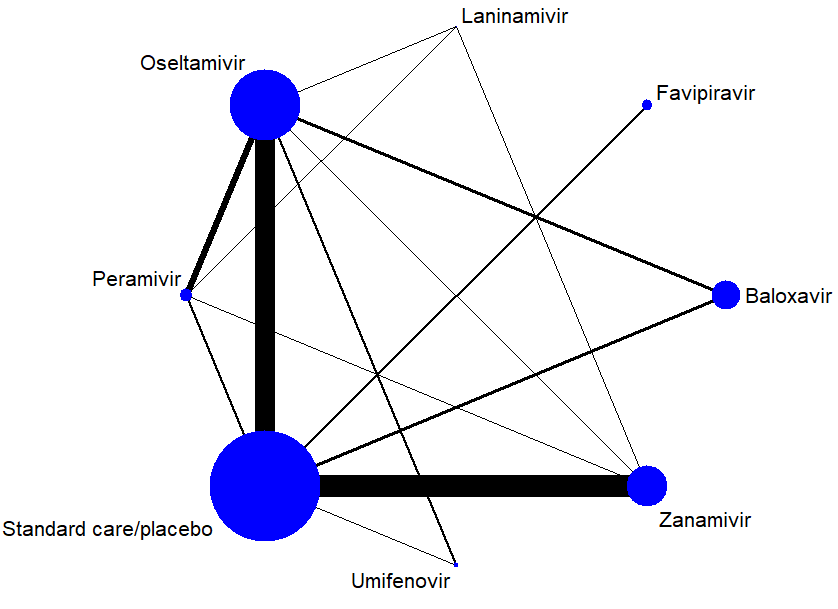

#### 6.5. Network plot for adverse events related to treatments

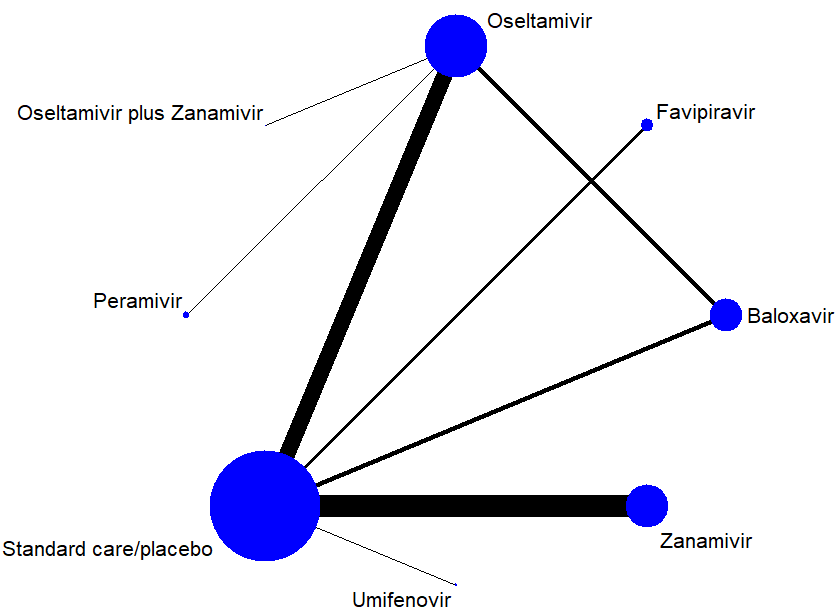

#### 6.6. Network plot for serious adverse events

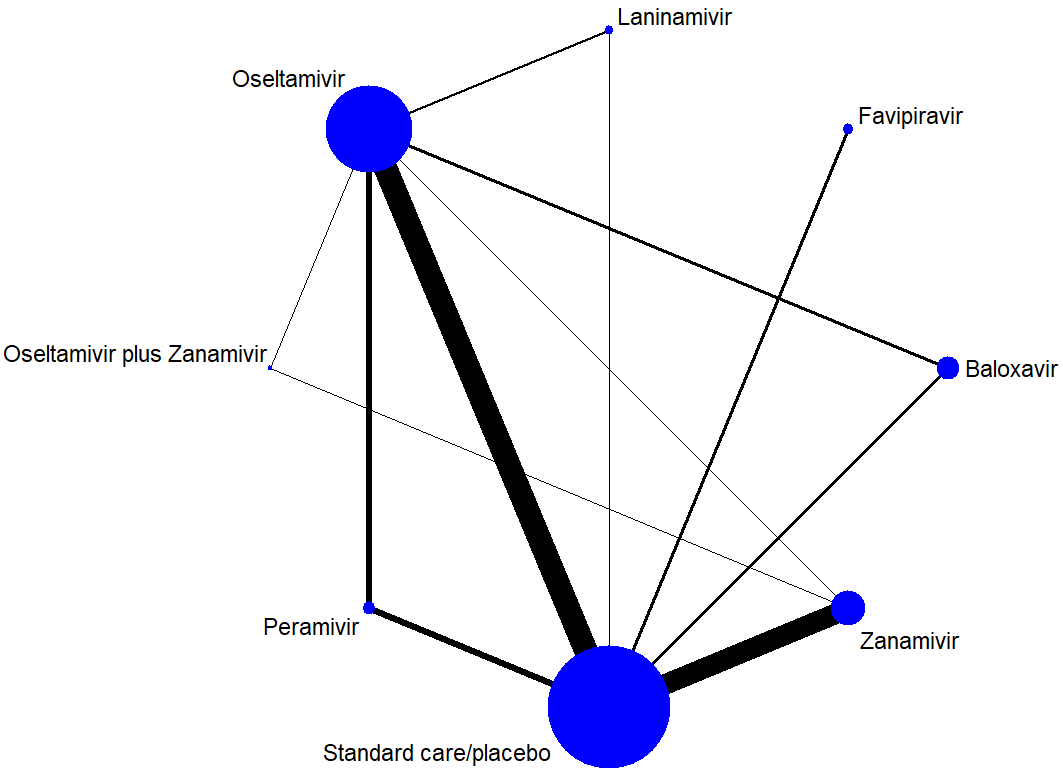

### Appendix 7. Assessment of between-study heterogeneity

| **Outcome** | **Comparison** | **No. study** | **I^2^** |
| --- | --- | --- | --- |
| Mortality | Baloxavir vs. Oseltamivir | 3 | 0% |
|  | Baloxavir vs. Standard care/placebo | 2 | 0% |
|  | Favipiravir vs. Standard care/placebo | 2 | 0% |
|  | Laninamivir vs. Standard care/placebo | 1 | NA |
|  | Oseltamivir vs. Peramivir | 2 | 0% |
|  | Oseltamivir vs. Standard care/placebo | 17 | 0% |
|  | Oseltamivir vs. Umifenovir | 1 | NA |
|  | Peramivir vs. Standard care/placebo | 1 | NA |
|  | Zanamivir vs. Standard care/placebo | 16 | 0% |
| Admission to hospital | Baloxavir vs. Oseltamivir | 3 | 0% |
|  | Baloxavir vs. Standard care/placebo | 2 | 0% |
|  | Favipiravir vs. Standard care/placebo | 2 | 0% |
|  | Laninamivir vs. Standard care/placebo | 1 | NA |
|  | Oseltamivir vs. Standard care/placebo | 20 | 0% |
|  | Oseltamivir vs. Umifenovir | 1 | NA |
|  | Zanamivir vs. Standard care/placebo | 3 | 36% |
| Admission to ICU | Oseltamivir vs. Peramivir | 1 | NA |
|  | Oseltamivir vs. Standard care/placebo | 1 | NA |
| Time to alleviation of symptoms | Amantadine vs. Standard care/placebo | 2 | 76% |
|  | Baloxavir vs. Oseltamivir | 2 | 0% |
|  | Baloxavir vs. Standard care/placebo | 3 | 0% |
|  | Favipiravir vs. Standard care/placebo | 2 | 0% |
|  | Laninamivir vs. Standard care/placebo | 1 | NA |
|  | Laninamivir vs. Oseltamivir | 4 | 36% |
|  | Laninamivir vs. Peramivir | 1 | NA |
|  | Laninamivir vs. Zanamivir | 1 | NA |
|  | Oseltamivir vs. Peramivir | 5 | 0% |
|  | Oseltamivir vs. Standard care/placebo | 22 | 88% |
|  | Oseltamivir vs. Umifenovir | 1 | NA |
|  | Oseltamivir vs. Zanamivir | 2 | 92% |
|  | Oseltamivir vs. Oseltamivir plus Zanamivir | 1 | NA |
|  | Oseltamivir plus Zanamivir vs. Zanamivir | 1 | NA |
|  | Peramivir vs. Zanamivir | 1 | NA |
|  | Peramivir vs. Standard care/placebo | 5 | 0% |
|  | Umifenovir vs. Standard care/placebo | 1 | NA |
|  | Zanamivir vs. Standard care/placebo | 15 | 23% |
| Duration of hospitalization | Oseltamivir vs. Standard care/placebo | 1 | NA |
|  | Oseltamivir vs. Peramivir | 1 | NA |
|  | Zanamivir vs. Standard care/placebo | 1 | NA |
| Any adverse events | Baloxavir vs. Oseltamivir | 3 | 0% |
|  | Baloxavir vs. Standard care/placebo | 3 | 0% |
|  | Favipiravir vs. Standard care/placebo | 2 | 66% |
|  | Laninamivir vs. Oseltamivir | 1 | NA |
|  | Laninamivir vs. Peramivir | 1 | NA |
|  | Laninamivir vs. Zanamivir | 1 | NA |
|  | Oseltamivir vs. Peramivir | 6 | 0% |
|  | Oseltamivir vs. Standard care/placebo | 17 | 24% |
|  | Oseltamivir vs. Umifenovir | 2 | 0% |
|  | Oseltamivir vs. Zanamivir | 1 | NA |
|  | Peramivir vs. Zanamivir | 1 | NA |
|  | Peramivir vs. Standard care/placebo | 2 | 44% |
|  | Umifenovir vs. Standard care/placebo | 1 | NA |
|  | Zanamivir vs. Standard care/placebo | 19 | 0% |
| Adverse events related to treatments | Baloxavir vs. Oseltamivir | 3 | 0% |
|  | Baloxavir vs. Standard care/placebo | 3 | 0% |
|  | Favipiravir vs. Standard care/placebo | 2 | 77% |
|  | Oseltamivir vs. Oseltamivir plus Zanamivir | 1 | NA |
|  | Oseltamivir vs. Peramivir | 1 | NA |
|  | Oseltamivir vs. Standard care/placebo | 12 | 0% |
|  | Umifenovir vs. Standard care/placebo | 1 | NA |
|  | Zanamivir vs. Standard care/placebo | 17 | 0% |
| Serious adverse events | Baloxavir vs. Oseltamivir | 3 | 0% |
|  | Baloxavir vs. Standard care/placebo | 3 | 0% |
|  | Favipiravir vs. Standard care/placebo | 3 | 0% |
|  | Laninamivir vs. Standard care/placebo | 1 | NA |
|  | Laninamivir vs. Oseltamivir | 2 | 0% |
|  | Oseltamivir vs. Peramivir | 5 | 0% |
|  | Oseltamivir vs. Standard care/placebo | 22 | 0% |
|  | Oseltamivir vs. Zanamivir | 1 | NA |
|  | Oseltamivir vs. Oseltamivir plus Zanamivir | 1 | NA |
|  | Oseltamivir plus Zanamivir vs. Zanamivir | 1 | NA |
|  | Peramivir vs. Standard care/placebo | 5 | 0% |
|  | Zanamivir vs. Standard care/placebo | 19 | 0% |

NA, not applicable.

### Appendix 8. Assessment of global inconsistency

| **Outcome** | **P value** |
| --- | --- |
| Mortality | 0.942 |
| Admission to hospital | 0.947 |
| Admission to ICU | NA |
| Time to alleviation of symptoms | 0.377 |
| Duration of hospitalization | NA |
| Any adverse events | 0.989 |
| Adverse events to treatments | 0.780 |
| Serious adverse events | 0.959 |

NA, not applicable.

### Appendix 9. Direct, indirect, and network treatment estimates

#### 9.1. Direct, indirect, and network treatment estimates for mortality

| **Comparison** | **k** | **Prop** | **NMA (95% CI)** | **Direct (95% CI)** | **Indirect (95% CI)** | **RoR (95% CI)** | **z** | **Incoherence p-value** |
| --- | --- | --- | --- | --- | --- | --- | --- | --- |
| Baloxavir vs. Favipiravir | 0 | 0 | 1.45 (0.05, 38.54) | NA | 1.45 (0.05, 38.54) | NA | NA | NA |
| Baloxavir vs. Laninamivir | 0 | 0 | 1.67 (0.02, 122.20) | NA | 1.67 (0.02, 122.20) | NA | NA | NA |
| Baloxavir vs. Oseltamivir | 3 | 0.89 | 0.98 (0.19, 5.11) | 0.83 (0.14, 4.80) | 3.56 (0.03, 473.05) | 0.23 (0.00, 42.18) | -0.6 | 0.584 |
| Baloxavir vs. Peramivir | 0 | 0 | 2.11 (0.13, 34.01) | NA | 2.11 (0.13, 34.01) | NA | NA | NA |
| Baloxavir vs. Standard care/placebo | 2 | 0.5 | 0.83 (0.14, 4.82) | 1.47 (0.12, 17.50) | 0.46 (0.04, 5.65) | 3.17 (0.09, 107.30) | 0.64 | 0.520 |
| Baloxavir vs. Umifenovir | 0 | 0 | 0.97 (0.01, 68.12) | NA | 0.97 (0.01, 68.12) | NA | NA | NA |
| Baloxavir vs. Zanamivir | 0 | 0 | 0.95 (0.13, 7.02) | NA | 0.95 (0.13, 7.02) | NA | NA | NA |
| Favipiravir vs. Laninamivir | 0 | 0 | 1.15 (0.01, 139.67) | NA | 1.15 (0.01, 139.67) | NA | NA | NA |
| Favipiravir vs. Oseltamivir | 0 | 0 | 0.68 (0.04, 12.50) | NA | 0.68 (0.04, 12.50) | NA | NA | NA |
| Favipiravir vs. Peramivir | 0 | 0 | 1.46 (0.04, 54.38) | NA | 1.46 (0.04, 54.38) | NA | NA | NA |
| Favipiravir vs. Standard care/placebo | 2 | 1 | 0.57 (0.04, 9.15) | 0.57 (0.04, 9.15) | NA | NA | NA | NA |
| Favipiravir vs. Umifenovir | 0 | 0 | 0.67 (0.01, 88.54) | NA | 0.67 (0.01, 88.54) | NA | NA | NA |
| Favipiravir vs. Zanamivir | 0 | 0 | 0.65 (0.03, 12.26) | NA | 0.65 (0.03, 12.26) | NA | NA | NA |
| Laninamivir vs. Oseltamivir | 0 | 0 | 0.59 (0.01, 32.78) | NA | 0.59 (0.01, 32.78) | NA | NA | NA |
| Laninamivir vs. Peramivir | 0 | 0 | 1.26 (0.01, 120.47) | NA | 1.26 (0.01, 120.47) | NA | NA | NA |
| Laninamivir vs. Standard care/placebo | 1 | 1 | 0.50 (0.01, 24.96) | 0.50 (0.01, 24.96) | NA | NA | NA | NA |
| Laninamivir vs. Umifenovir | 0 | 0 | 0.58 (0.00, 159.49) | NA | 0.58 (0.00, 159.49) | NA | NA | NA |
| Laninamivir vs. Zanamivir | 0 | 0 | 0.57 (0.01, 31.98) | NA | 0.57 (0.01, 31.98) | NA | NA | NA |
| Oseltamivir vs. Peramivir | 2 | 0.68 | 2.15 (0.22, 20.79) | 2.28 (0.14, 35.98) | 1.89 (0.03, 102.99) | 1.21 (0.01, 155.36) | 0.08 | 0.940 |
| Oseltamivir vs. Standard care/placebo | 17 | 0.96 | 0.84 (0.34, 2.07) | 0.84 (0.33, 2.10) | 1.05 (0.01, 79.37) | 0.80 (0.01, 66.54) | -0.1 | 0.920 |
| Oseltamivir vs. Umifenovir | 1 | 1 | 0.99 (0.02, 49.67) | 0.99 (0.02, 49.67) | NA | NA | NA | NA |
| Oseltamivir vs. Zanamivir | 0 | 0 | 0.96 (0.26, 3.58) | NA | 0.96 (0.26, 3.58) | NA | NA | NA |
| Peramivir vs. Standard care/placebo | 1 | 0.36 | 0.39 (0.04, 4.04) | 0.44 (0.01, 21.74) | 0.37 (0.02, 6.73) | 1.21 (0.01, 155.36) | 0.08 | 0.940 |
| Peramivir vs. Umifenovir | 0 | 0 | 0.46 (0.00, 42.65) | NA | 0.46 (0.00, 42.65) | NA | NA | NA |
| Peramivir vs. Zanamivir | 0 | 0 | 0.45 (0.04, 5.57) | NA | 0.45 (0.04, 5.57) | NA | NA | NA |
| Umifenovir vs. Standard care/placebo | 0 | 0 | 0.85 (0.02, 47.31) | NA | 0.85 (0.02, 47.31) | NA | NA | NA |
| Zanamivir vs. Standard care/placebo | 16 | 1 | 0.88 (0.34, 2.28) | 0.88 (0.34, 2.28) | NA | NA | NA | NA |
| Umifenovir vs. Zanamivir | 0 | 0 | 0.97 (0.02, 60.43) | NA | 0.97 (0.02, 60.43) | NA | NA | NA |

Comparison: treatment comparison; k: number of studies providing direct evidence; prop: direct evidence proportion; NMA: estimated treatment effect (RR) in network meta-analysis; direct: estimated treatment effect (RR) derived from direct evidence; indirect: estimated treatment effect (RR) derived from indirect evidence; RoR: Ratio of Ratios (direct versus indirect); z: z-value of test for disagreement (direct versus indirect): Incoherence p-value: p-value of test for disagreement (direct versus indirect).

#### 9.2. Direct, indirect, and network treatment estimates for admission to hospital

| **Comparison** | **k** | **Prop** | **NMA (95% CI)** | **Direct (95% CI)** | **Indirect (95% CI)** | **RoR (95% CI)** | **z** | **Incoherence p-value** |
| --- | --- | --- | --- | --- | --- | --- | --- | --- |
| Baloxavir vs. Favipiravir | 0 | 0 | 0.42 (0.02, 10.31) | NA | 0.42 (0.02, 10.31) | NA | NA | NA |
| Baloxavir vs. Laninamivir | 0 | 0 | 0.22 (0.00, 15.07) | NA | 0.22 (0.00, 15.07) | NA | NA | NA |
| Baloxavir vs. Oseltamivir | 3 | 0.93 | 0.30 (0.06, 1.47) | 0.29 (0.06, 1.50) | 0.51 (0.00, 186.48) | 0.57 (0.00, 256.42) | -0.2 | 0.855 |
| Baloxavir vs. Standard care/placebo | 2 | 0.72 | 0.24 (0.05, 1.19) | 0.25 (0.04, 1.62) | 0.23 (0.01, 4.54) | 1.07 (0.03, 36.16) | 0.04 | 0.971 |
| Baloxavir vs. Umifenovir | 0 | 0 | 0.90 (0.03, 31.77) | NA | 0.90 (0.03, 31.77) | NA | NA | NA |
| Baloxavir vs. Zanamivir | 0 | 0 | 0.21 (0.04, 1.05) | NA | 0.21 (0.04, 1.05) | NA | NA | NA |
| Favipiravir vs. Laninamivir | 0 | 0 | 0.52 (0.00, 63.05) | NA | 0.52 (0.00, 63.05) | NA | NA | NA |
| Favipiravir vs. Oseltamivir | 0 | 0 | 0.72 (0.04, 11.76) | NA | 0.72 (0.04, 11.76) | NA | NA | NA |
| Favipiravir vs. Standard care/placebo | 2 | 1 | 0.57 (0.04, 9.15) | 0.57 (0.04, 9.15) | NA | NA | NA | NA |
| Favipiravir vs. Umifenovir | 0 | 0 | 2.13 (0.03, 148.75) | NA | 2.13 (0.03, 148.75) | NA | NA | NA |
| Favipiravir vs. Zanamivir | 0 | 0 | 0.49 (0.03, 7.96) | NA | 0.49 (0.03, 7.96) | NA | NA | NA |
| Laninamivir vs. Oseltamivir | 0 | 0 | 1.38 (0.03, 70.38) | NA | 1.38 (0.03, 70.38) | NA | NA | NA |
| Laninamivir vs. Standard care/placebo | 0 | 0 | 1.10 (0.02, 55.19) | NA | 1.10 (0.02, 55.19) | NA | NA | NA |
| Laninamivir vs. Umifenovir | 0 | 0 | 4.09 (0.03, 649.73) | NA | 4.09 (0.03, 649.73) | NA | NA | NA |
| Laninamivir vs. Zanamivir | 1 | 1 | 0.93 (0.02, 45.94) | 0.93 (0.02, 45.94) | NA | NA | NA | NA |
| Oseltamivir vs. Standard care/placebo | 20 | 1 | 0.80 (0.54, 1.18) | 0.80 (0.54, 1.18) | 0.24 (0.00, 371.95) | 3.36 (0.00, 5279.20) | 0.32 | 0.747 |
| Oseltamivir vs. Umifenovir | 1 | 1 | 2.97 (0.12, 72.51) | 2.97 (0.12, 72.51) | NA | NA | NA | NA |
| Oseltamivir vs. Zanamivir | 0 | 0 | 0.68 (0.39, 1.17) | NA | 0.68 (0.39, 1.17) | NA | NA | NA |
| Umifenovir vs. Standard care/placebo | 0 | 0 | 0.27 (0.01, 6.72) | NA | 0.27 (0.01, 6.72) | NA | NA | NA |
| Zanamivir vs. Standard care/placebo | 3 | 1 | 1.18 (0.81, 1.72) | 1.18 (0.81, 1.72) | NA | NA | NA | NA |
| Umifenovir vs. Zanamivir | 0 | 0 | 0.23 (0.01, 5.83) | NA | 0.23 (0.01, 5.83) | NA | NA | NA |

Comparison: treatment comparison; k: number of studies providing direct evidence; prop: direct evidence proportion; NMA: estimated treatment effect (RR) in network meta-analysis; direct: estimated treatment effect (RR) derived from direct evidence; indirect: estimated treatment effect (RR) derived from indirect evidence; RoR: Ratio of Ratios (direct versus indirect); z: z-value of test for disagreement (direct versus indirect): Incoherence p-value: p-value of test for disagreement (direct versus indirect).

#### 9.3. Direct, indirect, and network treatment estimates for admission to ICU

| **Comparison** | **k** | **Prop** | **NMA (95% CI)** | **Direct (95% CI)** | **Indirect (95% CI)** | **Diff (95% CI)** | **z** | **Incoherence p-value** |
| --- | --- | --- | --- | --- | --- | --- | --- | --- |
| Oseltamivir vs. Peramivir | 1 | 1 | -0.021 (-0.057, 0.015) | -0.021 (-0.057, 0.015) | NA | NA | NA | NA |
| Oseltamivir vs. Standard care/placebo | 1 | 1 | -0.002 (-0.007, 0.003) | -0.002 (-0.007, 0.003) | NA | NA | NA | NA |
| Peramivir vs. Standard care/placebo | 0 | 0 | 0.019 (-0.017, 0.055) | NA | 0.019 (-0.017, 0.055) | NA | NA | NA |

Comparison: treatment comparison; k: number of studies providing direct evidence; prop: direct evidence proportion; NMA: estimated treatment effect (RD) in network meta-analysis; direct: estimated treatment effect (RD) derived from direct evidence; indirect: estimated treatment effect (RD) derived from indirect evidence; Diff: difference between direct and indirect treatment estimates; z: z-value of test for disagreement (direct versus indirect): Incoherence p-value: p-value of test for disagreement (direct versus indirect).

#### 9.4. Direct, indirect, and network treatment estimates for time to alleviation of symptoms

| **Comparison** | **k** | **Prop** | **NMA (95% CI)** | **Direct (95% CI)** | **Indirect (95% CI)** | **Diff (95% CI)** | **z** | **Incoherence p-value** |
| --- | --- | --- | --- | --- | --- | --- | --- | --- |
| Amantadine vs. Baloxavir | 0 | 0 | 0.24 (-0.41, 0.89) | NA | 0.24 (-0.41, 0.89) | NA | NA | NA |
| Amantadine vs. Favipiravir | 0 | 0 | -0.32 (-1.04, 0.40) | NA | -0.32 (-1.04, 0.40) | NA | NA | NA |
| Amantadine vs. Laninamivir | 0 | 0 | -0.21 (-0.89, 0.47) | NA | -0.21 (-0.89, 0.47) | NA | NA | NA |
| Amantadine vs. Oseltamivir | 0 | 0 | -0.03 (-0.59, 0.52) | NA | -0.03 (-0.59, 0.52) | NA | NA | NA |
| Amantadine vs. Oseltamivir plus Zanamivir | 0 | 0 | -2.14 (-3.65, -0.62) | NA | -2.14 (-3.65, -0.62) | NA | NA | NA |
| Amantadine vs. Peramivir | 0 | 0 | 0.17 (-0.45, 0.79) | NA | 0.17 (-0.45, 0.79) | NA | NA | NA |
| Amantadine vs. Standard care/placebo | 2 | 1 | -0.78 (-1.30, -0.26) | -0.78 (-1.30, -0.26) | NA | NA | NA | NA |
| Amantadine vs. Umifenovir | 0 | 0 | 0.32 (-0.39, 1.02) | NA | 0.32 (-0.39, 1.02) | NA | NA | NA |
| Amantadine vs. Zanamivir | 0 | 0 | -0.10 (-0.68, 0.48) | NA | -0.10 (-0.68, 0.48) | NA | NA | NA |
| Baloxavir vs. Favipiravir | 0 | 0 | -0.56 (-1.19, 0.07) | NA | -0.56 (-1.19, 0.07) | NA | NA | NA |
| Baloxavir vs. Laninamivir | 0 | 0 | -0.45 (-1.02, 0.12) | NA | -0.45 (-1.02, 0.12) | NA | NA | NA |
| Baloxavir vs. Oseltamivir | 2 | 0.34 | -0.27 (-0.68, 0.14) | -0.27 (-0.97, 0.44) | -0.27 (-0.78, 0.23) | 0.01 (-0.86, 0.87) | 0.02 | 0.984 |
| Baloxavir vs. Oseltamivir plus Zanamivir | 0 | 0 | -2.38 (-3.85, -0.91) | NA | -2.38 (-3.85, -0.91) | NA | NA | NA |
| Baloxavir vs. Peramivir | 0 | 0 | -0.07 (-0.57, 0.43) | NA | -0.07 (-0.57, 0.43) | NA | NA | NA |
| Baloxavir vs. Standard care/placebo | 3 | 0.85 | -1.02 (-1.41, -0.63) | -1.04 (-1.46, -0.61) | -0.95 (-1.94, 0.04) | -0.09 (-1.17, 0.99) | -0.16 | 0.872 |
| Baloxavir vs. Umifenovir | 0 | 0 | 0.08 (-0.53, 0.68) | NA | 0.08 (-0.53, 0.68) | NA | NA | NA |
| Baloxavir vs. Zanamivir | 0 | 0 | -0.34 (-0.80, 0.12) | NA | -0.34 (-0.80, 0.12) | NA | NA | NA |
| Favipiravir vs. Laninamivir | 0 | 0 | 0.11 (-0.54, 0.77) | NA | 0.11 (-0.54, 0.77) | NA | NA | NA |
| Favipiravir vs. Oseltamivir | 0 | 0 | 0.29 (-0.24, 0.81) | NA | 0.29 (-0.24, 0.81) | NA | NA | NA |
| Favipiravir vs. Oseltamivir plus Zanamivir | 0 | 0 | -1.82 (-3.33, -0.31) | NA | -1.82 (-3.33, -0.31) | NA | NA | NA |
| Favipiravir vs. Peramivir | 0 | 0 | 0.49 (-0.11, 1.08) | NA | 0.49 (-0.11, 1.08) | NA | NA | NA |
| Favipiravir vs. Standard care/placebo | 2 | 1 | -0.46 (-0.96, 0.03) | -0.46 (-0.96, 0.03) | NA | NA | NA | NA |
| Favipiravir vs. Umifenovir | 0 | 0 | 0.64 (-0.05, 1.32) | NA | 0.64 (-0.05, 1.32) | NA | NA | NA |
| Favipiravir vs. Zanamivir | 0 | 0 | 0.22 (-0.34, 0.77) | NA | 0.22 (-0.34, 0.77) | NA | NA | NA |
| Laninamivir vs. Oseltamivir | 4 | 0.76 | 0.18 (-0.24, 0.59) | 0.03 (-0.45, 0.50) | 0.64 (-0.20, 1.49) | -0.61 (-1.59, 0.36) | -1.24 | 0.215 |
| Laninamivir vs. Oseltamivir plus Zanamivir | 0 | 0 | -1.93 (-3.40, -0.46) | NA | -1.93 (-3.40, -0.46) | NA | NA | NA |
| Laninamivir vs. Peramivir | 1 | 0.2 | 0.38 (-0.13, 0.88) | 0.24 (-0.89, 1.37) | 0.41 (-0.15, 0.98) | -0.17 (-1.43, 1.09) | -0.27 | 0.790 |
| Laninamivir vs. Standard care/placebo | 1 | 0.25 | -0.57 (-1.01, -0.14) | -0.06 (-0.93, 0.81) | -0.74 (-1.24, -0.24) | 0.68 (-0.32, 1.68) | 1.33 | 0.184 |
| Laninamivir vs. Umifenovir | 0 | 0 | 0.52 (-0.09, 1.14) | NA | 0.52 (-0.09, 1.14) | NA | NA | NA |
| Laninamivir vs. Zanamivir | 1 | 0.2 | 0.11 (-0.38, 0.59) | -0.06 (-1.16, 1.04) | 0.15 (-0.39, 0.69) | -0.21 (-1.43, 1.02) | -0.33 | 0.740 |
| Oseltamivir vs. Oseltamivir plus Zanamivir | 1 | 0.96 | -2.11 (-3.52, -0.69) | -2.67 (-4.12, -1.22) | 10.39 (3.58, 17.21) | -13.06 (-20.03, -6.09) | -3.67 | 0.000 |
| Oseltamivir vs. Peramivir | 5 | 0.72 | 0.20 (-0.12, 0.52) | 0.23 (-0.14, 0.60) | 0.13 (-0.48, 0.73) | 0.10 (-0.61, 0.81) | 0.29 | 0.775 |
| Oseltamivir vs. Standard care/placebo | 22 | 0.79 | -0.75 (-0.93, -0.57) | -0.74 (-0.95, -0.53) | -0.79 (-1.20, -0.39) | 0.05 (-0.40, 0.51) | 0.22 | 0.822 |
| Oseltamivir vs. Umifenovir | 1 | 0.59 | 0.35 (-0.12, 0.82) | 0.42 (-0.19, 1.03) | 0.24 (-0.49, 0.98) | 0.18 (-0.77, 1.13) | 0.36 | 0.715 |
| Oseltamivir vs. Zanamivir | 2 | 0.16 | -0.07 (-0.37, 0.23) | -0.74 (-1.47, 0.00) | 0.06 (-0.27, 0.39) | -0.80 (-1.60, 0.01) | -1.94 | 0.053 |
| Oseltamivir plus Zanamivir vs. Peramivir | 0 | 0 | 2.31 (0.86, 3.76) | NA | 2.31 (0.86, 3.76) | NA | NA | NA |
| Oseltamivir plus Zanamivir vs. Standard care/placebo | 0 | 0 | 1.36 (-0.07, 2.78) | NA | 1.36 (-0.07, 2.78) | NA | NA | NA |
| Oseltamivir plus Zanamivir vs. Umifenovir | 0 | 0 | 2.46 (0.97, 3.94) | NA | 2.46 (0.97, 3.94) | NA | NA | NA |
| Oseltamivir plus Zanamivir vs. Zanamivir | 1 | 0.6 | 2.04 (0.60, 3.47) | -0.16 (-2.01, 1.69) | 5.32 (3.06, 7.59) | -5.48 (-8.41, -2.56) | -3.67 | 0.000 |
| Peramivir vs. Standard care/placebo | 5 | 0.38 | -0.95 (-1.28, -0.62) | -0.98 (-1.52, -0.44) | -0.93 (-1.35, -0.51) | -0.04 (-0.73, 0.64) | -0.13 | 0.898 |
| Peramivir vs. Umifenovir | 0 | 0 | 0.15 (-0.41, 0.70) | NA | 0.15 (-0.41, 0.70) | NA | NA | NA |
| Peramivir vs. Zanamivir | 1 | 0.21 | -0.27 (-0.67, 0.13) | -0.30 (-1.18, 0.58) | -0.26 (-0.71, 0.19) | -0.04 (-1.02, 0.95) | -0.07 | 0.941 |
| Umifenovir vs. Standard care/placebo | 1 | 0.45 | -1.10 (-1.57, -0.63) | -1.00 (-1.71, -0.29) | -1.18 (-1.81, -0.54) | 0.18 (-0.77, 1.13) | 0.36 | 0.715 |
| Zanamivir vs. Standard care/placebo | 15 | 0.86 | -0.68 (-0.93, -0.43) | -0.78 (-1.05, -0.51) | -0.08 (-0.76, 0.59) | -0.69 (-1.42, 0.03) | -1.87 | 0.062 |
| Umifenovir vs. Zanamivir | 0 | 0 | -0.42 (-0.95, 0.11) | NA | -0.42 (-0.95, 0.11) | NA | NA | NA |

Comparison: treatment comparison; k: number of studies providing direct evidence; prop: direct evidence proportion; NMA: estimated treatment effect (MD) in network meta-analysis; direct: estimated treatment effect (MD) derived from direct evidence; indirect: estimated treatment effect (MD) derived from indirect evidence; Diff: difference between direct and indirect treatment estimates; z: z-value of test for disagreement (direct versus indirect): Incoherence p-value: p-value of test for disagreement (direct versus indirect).

#### 9.5. Direct, indirect, and network treatment estimates for duration of hospitalization

| **Comparison** | **k** | **Prop** | **NMA (95% CI)** | **Direct (95% CI)** | **Indirect (95% CI)** | **Diff (95% CI)** | **z** | **Incoherence p-value** |
| --- | --- | --- | --- | --- | --- | --- | --- | --- |
| Oseltamivir vs. Peramivir | 1 | 1 | 0.69 (0.35, 1.03) | 0.69 (0.35, 1.03) | NA | NA | NA | NA |
| Oseltamivir vs. Standard care/placebo | 1 | 1 | -0.35 (-3.19, 2.49) | -0.35 (-3.19, 2.49) | NA | NA | NA | NA |
| Oseltamivir vs. Zanamivir | 0 | 0 | 0.75 (-2.33, 3.83) | NA | 0.75 (-2.33, 3.83) | NA | NA | NA |
| Peramivir vs. Standard care/placebo | 0 | 0 | -1.04 (-3.90, 1.82) | NA | -1.04 (-3.90, 1.82) | NA | NA | NA |
| Peramivir vs. Zanamivir | 0 | 0 | 0.06 (-3.04, 3.16) | NA | 0.06 (-3.04, 3.16) | NA | NA | NA |
| Zanamivir vs. Standard care/placebo | 1 | 1 | -1.10 (-2.30, 0.10) | -1.10 (-2.30, 0.10) | NA | NA | NA | NA |

Comparison: treatment comparison; k: number of studies providing direct evidence; prop: direct evidence proportion; NMA: estimated treatment effect (MD) in network meta-analysis; direct: estimated treatment effect (MD) derived from direct evidence; indirect: estimated treatment effect (MD) derived from indirect evidence; Diff: difference between direct and indirect treatment estimates; z: z-value of test for disagreement (direct versus indirect): Incoherence p-value: p-value of test for disagreement (direct versus indirect).

#### 9.6. Direct, indirect, and network treatment estimates for any adverse events

| **Comparison** | **k** | **Prop** | **NMA (95% CI)** | **Direct (95% CI)** | **Indirect (95% CI)** | **RoR (95% CI)** | **z** | **Incoherence p-value** |
| --- | --- | --- | --- | --- | --- | --- | --- | --- |
| Baloxavir vs. Favipiravir | 0 | 0 | 0.88 (0.72, 1.07) | NA | 0.88 (0.72, 1.07) | NA | NA | NA |
| Baloxavir vs. Laninamivir | 0 | 0 | 0.95 (0.04, 22.84) | NA | 0.95 (0.04, 22.84) | NA | NA | NA |
| Baloxavir vs. Oseltamivir | 3 | 0.77 | 0.87 (0.78, 0.97) | 0.87 (0.76, 0.99) | 0.87 (0.69, 1.10) | 1.00 (0.76, 1.31) | -0.01 | 0.994 |
| Baloxavir vs. Peramivir | 0 | 0 | 0.88 (0.76, 1.01) | NA | 0.88 (0.76, 1.01) | NA | NA | NA |
| Baloxavir vs. Standard care/placebo | 3 | 0.71 | 0.85 (0.75, 0.95) | 0.85 (0.74, 0.97) | 0.84 (0.68, 1.04) | 1.00 (0.78, 1.29) | 0.04 | 0.969 |
| Baloxavir vs. Umifenovir | 0 | 0 | 1.26 (0.50, 3.18) | NA | 1.26 (0.50, 3.18) | NA | NA | NA |
| Baloxavir vs. Zanamivir | 0 | 0 | 0.92 (0.81, 1.06) | NA | 0.92 (0.81, 1.06) | NA | NA | NA |
| Favipiravir vs. Laninamivir | 0 | 0 | 1.09 (0.05, 26.09) | NA | 1.09 (0.05, 26.09) | NA | NA | NA |
| Favipiravir vs. Oseltamivir | 0 | 0 | 0.99 (0.83, 1.18) | NA | 0.99 (0.83, 1.18) | NA | NA | NA |
| Favipiravir vs. Peramivir | 0 | 0 | 1.00 (0.83, 1.20) | NA | 1.00 (0.83, 1.20) | NA | NA | NA |
| Favipiravir vs. Standard care/placebo | 2 | 1 | 0.96 (0.82, 1.13) | 0.96 (0.82, 1.13) | NA | NA | NA | NA |
| Favipiravir vs. Umifenovir | 0 | 0 | 1.44 (0.57, 3.66) | NA | 1.44 (0.57, 3.66) | NA | NA | NA |
| Favipiravir vs. Zanamivir | 0 | 0 | 1.05 (0.88, 1.26) | NA | 1.05 (0.88, 1.26) | NA | NA | NA |
| Laninamivir vs. Oseltamivir | 1 | 0.67 | 0.91 (0.04, 21.80) | 1.00 (0.02, 48.82) | 0.76 (0.00, 185.00) | 1.32 (0.00, 1109.20) | 0.08 | 0.936 |
| Laninamivir vs. Peramivir | 1 | 0.67 | 0.92 (0.04, 21.98) | 0.93 (0.02, 45.57) | 0.89 (0.00, 217.91) | 1.05 (0.00, 886.21) | 0.01 | 0.988 |
| Laninamivir vs. Standard care/placebo | 0 | 0 | 0.89 (0.04, 21.20) | NA | 0.89 (0.04, 21.20) | NA | NA | NA |
| Laninamivir vs. Umifenovir | 0 | 0 | 1.32 (0.05, 36.05) | NA | 1.32 (0.05, 36.05) | NA | NA | NA |
| Laninamivir vs. Zanamivir | 1 | 0.67 | 0.97 (0.04, 23.18) | 0.87 (0.02, 42.31) | 1.21 (0.00, 296.81) | 0.72 (0.00, 608.52) | -0.1 | 0.924 |
| Oseltamivir vs. Peramivir | 6 | 0.42 | 1.01 (0.92, 1.10) | 1.04 (0.90, 1.19) | 0.99 (0.88, 1.11) | 1.05 (0.88, 1.25) | 0.52 | 0.603 |
| Oseltamivir vs. Standard care/placebo | 17 | 0.89 | 0.97 (0.92, 1.03) | 0.97 (0.91, 1.03) | 1.01 (0.86, 1.19) | 0.96 (0.81, 1.14) | -0.49 | 0.621 |
| Oseltamivir vs. Umifenovir | 2 | 0.39 | 1.45 (0.58, 3.64) | 0.99 (0.23, 4.32) | 1.85 (0.57, 6.01) | 0.54 (0.08, 3.53) | -0.65 | 0.516 |
| Oseltamivir vs. Zanamivir | 1 | < 0.01 | 1.06 (0.98, 1.16) | NA | 1.06 (0.98, 1.16) | NA | NA | NA |
| Peramivir vs. Standard care/placebo | 2 | 0.69 | 0.96 (0.89, 1.05) | 0.97 (0.88, 1.08) | 0.94 (0.81, 1.10) | 1.03 (0.86, 1.23) | 0.33 | 0.739 |
| Peramivir vs. Umifenovir | 0 | 0 | 1.44 (0.57, 3.62) | NA | 1.44 (0.57, 3.62) | NA | NA | NA |
| Peramivir vs. Zanamivir | 1 | < 0.01 | 1.05 (0.95, 1.17) | 0.93 (0.02, 45.23) | 1.05 (0.95, 1.17) | 0.88 (0.02, 42.96) | -0.06 | 0.949 |
| Umifenovir vs. Standard care/placebo | 1 | 0.61 | 0.67 (0.27, 1.68) | 0.53 (0.16, 1.70) | 0.98 (0.22, 4.28) | 0.54 (0.08, 3.53) | -0.65 | 0.516 |
| Zanamivir vs. Standard care/placebo | 19 | 1 | 0.92 (0.86, 0.98) | 0.92 (0.86, 0.98) | NA | NA | NA | NA |
| Umifenovir vs. Zanamivir | 0 | 0 | 0.73 (0.29, 1.84) | NA | 0.73 (0.29, 1.84) | NA | NA | NA |

Comparison: treatment comparison; k: number of studies providing direct evidence; prop: direct evidence proportion; NMA: estimated treatment effect (RR) in network meta-analysis; direct: estimated treatment effect (RR) derived from direct evidence; indirect: estimated treatment effect (RR) derived from indirect evidence; RoR: Ratio of Ratios (direct versus indirect); z: z-value of test for disagreement (direct versus indirect): Incoherence p-value: p-value of test for disagreement (direct versus indirect).

#### 9.7. Direct, indirect, and network treatment estimates for adverse events related to treatments

| **Comparison** | **k** | **Prop** | **NMA (95% CI)** | **Direct (95% CI)** | **Indirect (95% CI)** | | **RoR (95% CI)** | | **z** | | **Incoherence p-value** |
| --- | --- | --- | --- | --- | --- | --- | --- | --- | --- | --- | --- |
| Baloxavir vs. Favipiravir | 0 | 0 | 0.83 (0.56, 1.23) | NA | 0.83 (0.56, 1.23) | | NA | | NA | | NA |
| Baloxavir vs. Oseltamivir | 3 | 0.77 | 0.60 (0.46, 0.77) | 0.61 (0.46, 0.82) | 0.55 (0.32, 0.95) | | 1.10 (0.60, 2.03) | | 0.31 | | 0.759 |
| Baloxavir vs. Oseltamivir plus Zanamivir | 0 | 0 | 1.79 (0.21, 15.09) | NA | 1.79 (0.21, 15.09) | | NA | | NA | | NA |
| Baloxavir vs. Peramivir | 0 | 0 | 0.74 (0.51, 1.07) | NA | 0.74 (0.51, 1.07) | | NA | | NA | | NA |
| Baloxavir vs. Standard care/placebo | 3 | 0.74 | 0.74 (0.57, 0.95) | 0.77 (0.57, 1.04) | 0.66 (0.40, 1.09) | | 1.16 (0.65, 2.09) | | 0.51 | | 0.610 |
| Baloxavir vs. Umifenovir | 0 | 0 | 1.30 (0.52, 3.25) | NA | 1.30 (0.52, 3.25) | | NA | | NA | | NA |
| Baloxavir vs. Zanamivir | 0 | 0 | 0.70 (0.52, 0.95) | NA | 0.70 (0.52, 0.95) | | NA | | NA | | NA |
| Favipiravir vs. Oseltamivir | 0 | 0 | 0.72 (0.52, 0.99) | NA | 0.72 (0.52, 0.99) | | NA | | NA | | NA |
| Favipiravir vs. Oseltamivir plus Zanamivir | 0 | 0 | 2.15 (0.25, 18.33) | NA | 2.15 (0.25, 18.33) | | NA | | NA | | NA |
| Favipiravir vs. Peramivir | 0 | 0 | 0.89 (0.59, 1.35) | NA | 0.89 (0.59, 1.35) | | NA | | NA | | NA |
| Favipiravir vs. Standard care/placebo | 2 | 1 | 0.89 (0.66, 1.20) | 0.89 (0.66, 1.20) | NA | | NA | | NA | | NA |
| Favipiravir vs. Umifenovir | 0 | 0 | 1.56 (0.62, 3.97) | NA | 1.56 (0.62, 3.97) | | NA | | NA | | NA |
| Favipiravir vs. Zanamivir | 0 | 0 | 0.85 (0.61, 1.18) | NA | 0.85 (0.61, 1.18) | | NA | | NA | | NA |
| Oseltamivir vs. Oseltamivir plus Zanamivir | 1 | 1 | 3.00 (0.36, 24.92) | 3.00 (0.36, 24.92) | NA | | NA | | NA | | NA |
| Oseltamivir vs. Peramivir | 1 | 1 | 1.24 (0.96, 1.62) | 1.24 (0.96, 1.62) | NA | | NA | | NA | | NA |
| Oseltamivir vs. Standard care/placebo | 12 | 0.99 | 1.23 (1.10, 1.39) | 1.23 (1.10, 1.38) | 1.48 (0.55, 3.97) | | 0.83 (0.31, 2.26) | | -0.36 | | 0.723 |
| Oseltamivir vs. Umifenovir | 0 | 0 | 2.18 (0.89, 5.30) | NA | 2.18 (0.89, 5.30) | | NA | | NA | | NA |
| Oseltamivir vs. Zanamivir | 0 | 0 | 1.18 (0.98, 1.43) | NA | 1.18 (0.98, 1.43) | | NA | | NA | | NA |
| Oseltamivir plus Zanamivir vs. Peramivir | 0 | 0 | 0.41 (0.05, 3.50) | NA | 0.41 (0.05, 3.50) | | NA | | NA | | NA |
| Oseltamivir plus Zanamivir vs. Standard care/placebo | 0 | 0 | 0.41 (0.05, 3.43) | NA | 0.41 (0.05, 3.43) | | NA | | NA | | NA |
| Oseltamivir plus Zanamivir vs. Umifenovir | 0 | 0 | 0.73 (0.07, 7.21) | NA | 0.73 (0.07, 7.21) | | NA | | NA | | NA |
| Oseltamivir plus Zanamivir vs. Zanamivir | 0 | 0 | 0.39 (0.05, 3.30) | NA | 0.39 (0.05, 3.30) | | NA | | NA | | NA |
| Peramivir vs. Standard care/placebo | 0 | 0 | 0.99 (0.74, 1.32) | NA | 0.99 (0.74, 1.32) | | NA | | NA | | NA |
| Peramivir vs. Umifenovir | 0 | 0 | 1.75 (0.69, 4.42) | NA | 1.75 (0.69, 4.42) | | NA | | NA | | NA |
| Peramivir vs. Zanamivir | 0 | 0 | 0.95 (0.69, 1.31) | NA | 0.95 (0.69, 1.31) | | NA | | NA | | NA |
| Umifenovir vs. Standard care/placebo | 1 | 1 | 0.57 (0.23, 1.37) | 0.57 (0.23, 1.37) | NA | NA | | NA | | NA | |
| Zanamivir vs. Standard care/placebo | 17 | 1 | 1.05 (0.90, 1.21) | 1.05 (0.90, 1.21) | NA | | NA | | NA | | NA |
| Umifenovir vs. Zanamivir | 0 | 0 | 0.54 (0.22, 1.33) | NA | 0.54 (0.22, 1.33) | | NA | | NA | | NA |

Comparison: treatment comparison; k: number of studies providing direct evidence; prop: direct evidence proportion; NMA: estimated treatment effect (RR) in network meta-analysis; direct: estimated treatment effect (RR) derived from direct evidence; indirect: estimated treatment effect (RR) derived from indirect evidence; RoR: Ratio of Ratios (direct versus indirect); z: z-value of test for disagreement (direct versus indirect): Incoherence p-value: p-value of test for disagreement (direct versus indirect).

#### 9.8. Direct, indirect, and network treatment estimates for serious adverse events

| **Comparison** | **k** | **Prop** | **NMA (95% CI)** | **Direct (95% CI)** | **Indirect (95% CI)** | **Diff (95% CI)** | **z** | **Incoherence p-value** |
| --- | --- | --- | --- | --- | --- | --- | --- | --- |
| Baloxavir vs. Favipiravir | 0 | 0 | 0.002 (-0.005, 0.010) | NA | 0.002 (-0.005, 0.010) | NA | NA | NA |
| Baloxavir vs. Laninamivir | 0 | 0 | 0.005 (-0.004, 0.013) | NA | 0.005 (-0.004, 0.013) | NA | NA | NA |
| Baloxavir vs. Oseltamivir | 3 | 0.83 | 0.001 (-0.004, 0.005) | 0.001 (-0.004, 0.006) | -0.002 (-0.013, 0.008) | 0.004 (-0.008, 0.015) | 0.62 | 0.537 |
| Baloxavir vs. Oseltamivir plus Zanamivir | 0 | 0 | -0.015 (-0.036, 0.005) | NA | -0.015 (-0.036, 0.005) | NA | NA | NA |
| Baloxavir vs. Peramivir | 0 | 0 | 0.001 (-0.007, 0.008) | NA | 0.001 (-0.007, 0.008) | NA | NA | NA |
| Baloxavir vs. Standard care/placebo | 3 | 0.77 | 0.001 (-0.004, 0.005) | 0.000 (-0.005, 0.006) | 0.001 (-0.009, 0.010) | -0.000 (-0.011, 0.010) | -0.07 | 0.944 |
| Baloxavir vs. Zanamivir | 0 | 0 | -0.002 (-0.007, 0.004) | NA | -0.002 (-0.007, 0.004) | NA | NA | NA |
| Favipiravir vs. Laninamivir | 0 | 0 | 0.002 (-0.007, 0.011) | NA | 0.002 (-0.007, 0.011) | NA | NA | NA |
| Favipiravir vs. Oseltamivir | 0 | 0 | -0.002 (-0.008, 0.005) | NA | -0.002 (-0.008, 0.005) | NA | NA | NA |
| Favipiravir vs. Oseltamivir plus Zanamivir | 0 | 0 | -0.018 (-0.039, 0.004) | NA | -0.018 (-0.039, 0.004) | NA | NA | NA |
| Favipiravir vs. Peramivir | 0 | 0 | -0.001 (-0.010, 0.007) | NA | -0.001 (-0.010, 0.007) | NA | NA | NA |
| Favipiravir vs. Standard care/placebo | 3 | 1 | -0.002 (-0.007, 0.004) | -0.002 (-0.007, 0.004) | NA | NA | NA | NA |
| Favipiravir vs. Zanamivir | 0 | 0 | -0.004 (-0.011, 0.003) | NA | -0.004 (-0.011, 0.003) | NA | NA | NA |
| Laninamivir vs. Oseltamivir | 2 | 0.63 | -0.004 (-0.011, 0.003) | -0.006 (-0.015, 0.002) | 0.000 (-0.011, 0.012) | -0.007 (-0.021, 0.008) | -0.89 | 0.371 |
| Laninamivir vs. Oseltamivir plus Zanamivir | 0 | 0 | -0.020 (-0.041, 0.002) | NA | -0.020 (-0.041, 0.002) | NA | NA | NA |
| Laninamivir vs. Peramivir | 0 | 0 | -0.004 (-0.013, 0.006) | NA | -0.004 (-0.013, 0.006) | NA | NA | NA |
| Laninamivir vs. Standard care/placebo | 1 | 0.4 | -0.004 (-0.011, 0.003) | -0.000 (-0.011, 0.011) | -0.007 (-0.016, 0.003) | 0.007 (-0.008, 0.021) | 0.89 | 0.371 |
| Laninamivir vs. Zanamivir | 0 | 0 | -0.006 (-0.014, 0.002) | NA | -0.006 (-0.014, 0.002) | NA | NA | NA |
| Oseltamivir vs. Oseltamivir plus Zanamivir | 1 | 0.76 | -0.016 (-0.036, 0.004) | -0.010 (-0.033, 0.013) | -0.035 (-0.077, 0.006) | 0.025 (-0.022, 0.073) | 1.04 | 0.298 |
| Oseltamivir vs. Peramivir | 5 | 0.54 | 0.000 (-0.006, 0.007) | -0.002 (-0.011, 0.006) | 0.003 (-0.006, 0.012) | -0.005 (-0.018, 0.007) | -0.83 | 0.409 |
| Oseltamivir vs. Standard care/placebo | 22 | 0.9 | -0.000 (-0.003, 0.002) | -0.000 (-0.003, 0.002) | 0.001 (-0.007, 0.008) | -0.001 (-0.009, 0.007) | -0.18 | 0.857 |
| Oseltamivir vs. Zanamivir | 1 | 0.07 | -0.002 (-0.006, 0.002) | 0.006 (-0.010, 0.021) | -0.003 (-0.007, 0.001) | 0.009 (-0.008, 0.025) | 1.04 | 0.298 |
| Oseltamivir plus Zanamivir vs. Peramivir | 0 | 0 | 0.016 (-0.005, 0.037) | NA | 0.016 (-0.005, 0.037) | NA | NA | NA |
| Oseltamivir plus Zanamivir vs. Standard care/placebo | 0 | 0 | 0.016 (-0.004, 0.036) | NA | 0.016 (-0.004, 0.036) | NA | NA | NA |
| Oseltamivir plus Zanamivir vs. Zanamivir | 1 | 0.96 | 0.014 (-0.007, 0.034) | 0.016 (-0.005, 0.036) | -0.041 (-0.147, 0.064) | 0.057 (-0.050, 0.164) | 1.04 | 0.298 |
| Peramivir vs. Standard care/placebo | 5 | 0.51 | -0.000 (-0.007, 0.006) | -0.003 (-0.012, 0.006) | 0.002 (-0.007, 0.011) | -0.005 (-0.018, 0.007) | -0.82 | 0.410 |
| Peramivir vs. Zanamivir | 0 | 0 | -0.003 (-0.010, 0.004) | NA | -0.003 (-0.010, 0.004) | NA | NA | NA |
| Zanamivir vs. Standard care/placebo | 19 | 0.95 | 0.002 (-0.001, 0.006) | 0.003 (-0.001, 0.006) | -0.006 (-0.022, 0.010) | 0.009 (-0.008, 0.025) | 1.04 | 0.298 |

Comparison: treatment comparison; k: number of studies providing direct evidence; prop: direct evidence proportion; NMA: estimated treatment effect (RD) in network meta-analysis; direct: estimated treatment effect (RD) derived from direct evidence; indirect: estimated treatment effect (RD) derived from indirect evidence; Diff: difference between direct and indirect treatment estimates; z: z-value of test for disagreement (direct versus indirect): Incoherence p-value: p-value of test for disagreement (direct versus indirect).

### Appendix 10. Network estimates with GRADE ratings

#### 10.1. Network estimates with GRADE ratings for mortality

| **Comparison groups** | | **Network relative estimate (RR)** | | | **Low risk patients** | | | | **High risk patients** | | | |
| --- | --- | --- | --- | --- | --- | --- | --- | --- | --- | --- | --- | --- |
|  |  |  |  |  | **Network absolute** **estimate (per 1000)** | | | **GRADE rating** | **Network absolute estimate (per 1000)** | | | **GRADE rating** |
| **Treatment 1** | **Treatment 2** | **Point estimate** | **95% CI lower limit** | **95% CI upper limit** | **Point estimate** | **95% CI lower limit** | **95% CI upper limit** |  | **Point estimate** | **95% CI lower limit** | **95% CI upper limit** |  |
| Baloxavir | Favipiravir | 1.45 | 0.05 | 38.54 | 0.05 | -0.11 | 4.28 | High | 0.51 | -1.08 | 42.80 | High |
| Baloxavir | Laninamivir | 1.67 | 0.02 | 122.20 | 0.07 | -0.10 | 12.12 | High | 0.67 | -0.98 | 121.20 | High |
| Baloxavir | Oseltamivir | 0.98 | 0.19 | 5.11 | 0.00 | -0.14 | 0.69 | High | -0.03 | -1.36 | 6.90 | High |
| Baloxavir | Peramivir | 2.11 | 0.13 | 34.01 | 0.09 | -0.07 | 2.57 | High | 0.87 | -0.68 | 25.75 | High |
| Baloxavir | Standard care/placebo | 0.83 | 0.14 | 4.82 | -0.03 | -0.17 | 0.76 | High | -0.34 | -1.72 | 7.64 | High |
| Baloxavir | Umifenovir | 0.97 | 0.01 | 68.12 | -0.01 | -0.17 | 11.41 | High | -0.05 | -1.68 | 114.10 | High |
| Baloxavir | Zanamivir | 0.95 | 0.13 | 7.02 | -0.01 | -0.15 | 1.06 | High | -0.09 | -1.53 | 10.60 | High |
| Favipiravir | Laninamivir | 1.15 | 0.01 | 139.67 | 0.02 | -0.10 | 13.87 | High | 0.15 | -0.99 | 138.67 | High |
| Favipiravir | Oseltamivir | 0.68 | 0.04 | 12.50 | -0.05 | -0.16 | 1.93 | High | -0.54 | -1.61 | 19.32 | High |
| Favipiravir | Peramivir | 1.46 | 0.04 | 54.38 | 0.04 | -0.07 | 4.16 | High | 0.36 | -0.75 | 41.64 | High |
| Favipiravir | Standard care/placebo | 0.57 | 0.04 | 9.15 | -0.09 | -0.19 | 1.63 | High | -0.86 | -1.92 | 16.30 | High |
| Favipiravir | Umifenovir | 0.67 | 0.01 | 88.54 | -0.06 | -0.17 | 14.88 | High | -0.56 | -1.68 | 148.82 | High |
| Favipiravir | Zanamivir | 0.65 | 0.03 | 12.26 | -0.06 | -0.17 | 1.98 | High | -0.62 | -1.71 | 19.82 | High |
| Laninamivir | Oseltamivir | 0.59 | 0.01 | 32.78 | -0.07 | -0.17 | 5.34 | High | -0.69 | -1.66 | 53.39 | High |
| Laninamivir | Peramivir | 1.26 | 0.01 | 120.47 | 0.02 | -0.08 | 9.32 | High | 0.20 | -0.77 | 93.19 | High |
| Laninamivir | Standard care/placebo | 0.50 | 0.01 | 24.96 | -0.10 | -0.20 | 4.79 | High | -1.00 | -1.98 | 47.92 | High |
| Laninamivir | Umifenovir | 0.58 | 0.00 | 159.49 | -0.07 | -0.17 | 26.94 | High | -0.71 | -1.70 | 269.43 | High |
| Laninamivir | Zanamivir | 0.57 | 0.01 | 31.98 | -0.08 | -0.17 | 5.45 | High | -0.76 | -1.74 | 54.52 | High |
| Oseltamivir | Peramivir | 2.15 | 0.22 | 20.79 | 0.09 | -0.06 | 1.54 | High | 0.90 | -0.61 | 15.44 | High |
| Oseltamivir | Standard care/placebo | 0.84 | 0.34 | 2.07 | -0.03 | -0.13 | 0.21 | High | -0.32 | -1.32 | 2.14 | High |
| Oseltamivir | Umifenovir | 0.99 | 0.02 | 49.67 | 0.00 | -0.17 | 8.27 | High | -0.02 | -1.67 | 82.74 | High |
| Oseltamivir | Zanamivir | 0.96 | 0.26 | 3.58 | -0.01 | -0.13 | 0.45 | High | -0.07 | -1.30 | 4.54 | High |
| Peramivir | Standard care/placebo | 0.39 | 0.04 | 4.04 | -0.12 | -0.19 | 0.61 | High | -1.22 | -1.92 | 6.08 | High |
| Peramivir | Umifenovir | 0.46 | 0.00 | 42.65 | -0.09 | -0.17 | 7.08 | High | -0.92 | -1.70 | 70.81 | High |
| Peramivir | Zanamivir | 0.45 | 0.04 | 5.57 | -0.10 | -0.17 | 0.80 | High | -0.97 | -1.69 | 8.04 | High |
| Umifenovir | Standard care/placebo | 0.85 | 0.02 | 47.31 | -0.03 | -0.20 | 9.26 | High | -0.30 | -1.96 | 92.62 | High |
| Zanamivir | Standard care/placebo | 0.88 | 0.34 | 2.28 | -0.02 | -0.13 | 0.26 | High | -0.24 | -1.32 | 2.56 | High |
| Umifenovir | Zanamivir | 0.97 | 0.02 | 60.43 | -0.01 | -0.17 | 10.46 | High | -0.05 | -1.72 | 104.60 | High |

#### 10.2. Network estimates with GRADE ratings for admission to hospital

| **Comparison groups** | | **Network relative estimate (RR)** | | | **Low risk patients** | | | | | **High risk patients** | | | |
| --- | --- | --- | --- | --- | --- | --- | --- | --- | --- | --- | --- | --- | --- |
|  |  |  |  |  | **Network absolute estimate (per 1000)** | | | **GRADE rating** | **Network absolute estimate (per 1000)** | | | | **GRADE rating** |
| **Treatment 1** | **Treatment 2** | **Point estimate** | **95% CI lower limit** | **95% CI upper limit** | **Point estimate** | **95% CI lower limit** | **95% CI upper limit** |  | **Point estimate** | | **95% CI lower limit** | **95% CI upper limit** |  |
| Baloxavir | Favipiravir | 0.42 | 0.02 | 10.31 | -1 | -2 | 16 | High | -7 | | -12 | 111 | Low |
| Baloxavir | Laninamivir | 0.22 | 0.00 | 15.07 | -3 | -3 | 46 | High | -18 | | -23 | 325 | Very low |
| Baloxavir | Oseltamivir | 0.30 | 0.06 | 1.47 | -2 | -2 | 1 | High | -12 | | -16 | 8 | Moderate |
| Baloxavir | Standard care/placebo | 0.24 | 0.05 | 1.19 | -2 | -3 | 1 | High | -16 | | -20 | 4 | Low |
| Baloxavir | Umifenovir | 0.90 | 0.03 | 31.77 | 0 | -1 | 25 | High | -1 | | -5 | 174 | Low |
| Baloxavir | Zanamivir | 0.21 | 0.04 | 1.05 | -3 | -3 | 0 | High | -20 | | -24 | 1 | Moderate |
| Favipiravir | Laninamivir | 0.52 | 0.00 | 63.05 | -2 | -3 | 205 | High | -11 | | -23 | 977 | Very low |
| Favipiravir | Oseltamivir | 0.72 | 0.04 | 11.76 | -1 | -2 | 26 | High | -5 | | -16 | 181 | Very low |
| Favipiravir | Standard care/placebo | 0.57 | 0.04 | 9.15 | -1 | -3 | 24 | High | -9 | | -20 | 171 | Very low |
| Favipiravir | Umifenovir | 2.13 | 0.03 | 148.75 | 1 | -1 | 120 | High | 6 | | -5 | 838 | Very low |
| Favipiravir | Zanamivir | 0.49 | 0.03 | 7.96 | -2 | -3 | 25 | High | -13 | | -24 | 172 | Very low |
| Laninamivir | Oseltamivir | 1.38 | 0.03 | 70.38 | 1 | -2 | 167 | High | 6 | | -16 | 983 | Very low |
| Laninamivir | Standard care/placebo | 1.10 | 0.02 | 55.19 | 0 | -3 | 163 | High | 2 | | -21 | 979 | Low |
| Laninamivir | Umifenovir | 4.09 | 0.03 | 649.73 | 3 | -1 | 525 | High | 18 | | -5 | 994 | Very low |
| Laninamivir | Zanamivir | 0.93 | 0.02 | 45.94 | 0 | -3 | 159 | High | -2 | | -24 | 975 | Very low |
| Oseltamivir | Standard care/placebo | 0.80 | 0.54 | 1.18 | -1 | -1 | 1 | High | -4 | | -10 | 4 | High |
| Oseltamivir | Umifenovir | 2.97 | 0.12 | 72.51 | 2 | -1 | 58 | High | 11 | | -5 | 405 | Very low |
| Oseltamivir | Zanamivir | 0.68 | 0.39 | 1.17 | -1 | -2 | 1 | High | -8 | | -15 | 4 | Moderate |
| Umifenovir | Standard care/placebo | 0.27 | 0.01 | 6.72 | -2 | -3 | 17 | High | -15 | | -21 | 120 | Very low |
| Zanamivir | Standard care/placebo | 1.18 | 0.81 | 1.72 | 1 | -1 | 2 | High | 4 | | -4 | 15 | High |
| Umifenovir | Zanamivir | 0.23 | 0.01 | 5.83 | -3 | -4 | 17 | High | -19 | | -25 | 120 | Very low |

#### 10.3. Network estimates with GRADE ratings for admission to ICU

| **Comparison groups** | | **Network relative estimate** | | | **Network absolute estimate (per 1000)** | | | **GRADE rating** |
| --- | --- | --- | --- | --- | --- | --- | --- | --- |
| **Treatment 1** | **Treatment 2** | **Point estimate** | **95% CI lower limit** | **95% CI upper limit** | **Point estimate** | **95% CI lower limit** | **95% CI upper limit** |  |
| Oseltamivir | Peramivir | NA | NA | NA | -21 | -57 | 15 | Low |
| Oseltamivir | Standard care/placebo | NA | NA | NA | -2 | -7 | 3 | High |
| Peramivir | Standard care/placebo | NA | NA | NA | 19 | -17 | 55 | Very low |

NA, not applicable.

#### 10.4. Network estimates with GRADE ratings for time to alleviation of symptoms

| **Comparison groups** | | | **Network relative estimate** | | | **Network absolute estimate (days)** | | | **GRADE rating** |
| --- | --- | --- | --- | --- | --- | --- | --- | --- | --- |
| **Treatment 1** | **Treatment 2** | **Point estimate** | | **95% CI lower limit** | **95% CI upper limit** | **Point estimate** | **95% CI lower limit** | **95% CI upper limit** |  |
| Amantadine | Baloxavir | NA | | NA | NA | 0.24 | -0.41 | 0.89 | Moderate |
| Amantadine | Favipiravir | NA | | NA | NA | -0.32 | -1.04 | 0.4 | Low |
| Amantadine | Laninamivir | NA | | NA | NA | -0.21 | -0.89 | 0.47 | Moderate |
| Amantadine | Oseltamivir | NA | | NA | NA | -0.03 | -0.59 | 0.52 | Moderate |
| Amantadine | Oseltamivir plus Zanamivir | NA | | NA | NA | -2.14 | -3.65 | -0.62 | Low |
| Amantadine | Peramivir | NA | | NA | NA | 0.17 | -0.45 | 0.79 | Moderate |
| Amantadine | Standard care/placebo | NA | | NA | NA | -0.78 | -1.3 | -0.26 | Low |
| Amantadine | Umifenovir | NA | | NA | NA | 0.32 | -0.39 | 1.02 | Low |
| Amantadine | Zanamivir | NA | | NA | NA | -0.1 | -0.68 | 0.48 | Moderate |
| Baloxavir | Favipiravir | NA | | NA | NA | -0.56 | -1.19 | 0.07 | Low |
| Baloxavir | Laninamivir | NA | | NA | NA | -0.45 | -1.02 | 0.12 | Low |
| Baloxavir | Oseltamivir | NA | | NA | NA | -0.27 | -0.68 | 0.14 | Moderate |
| Baloxavir | Oseltamivir plus Zanamivir | NA | | NA | NA | -2.38 | -3.85 | -0.91 | Low |
| Baloxavir | Peramivir | NA | | NA | NA | -0.07 | -0.57 | 0.43 | Moderate |
| Baloxavir | Standard care/placebo | NA | | NA | NA | -1.02 | -1.41 | -0.63 | Moderate |
| Baloxavir | Umifenovir | NA | | NA | NA | 0.08 | -0.53 | 0.68 | Moderate |
| Baloxavir | Zanamivir | NA | | NA | NA | -0.34 | -0.8 | 0.12 | Moderate |
| Favipiravir | Laninamivir | NA | | NA | NA | 0.11 | -0.54 | 0.77 | Moderate |
| Favipiravir | Oseltamivir | NA | | NA | NA | 0.29 | -0.24 | 0.81 | Moderate |
| Favipiravir | Oseltamivir plus Zanamivir | NA | | NA | NA | -1.82 | -3.33 | -0.31 | Low |
| Favipiravir | Peramivir | NA | | NA | NA | 0.49 | -0.11 | 1.08 | Low |
| Favipiravir | Standard care/placebo | NA | | NA | NA | -0.46 | -0.96 | 0.03 | Moderate |
| Favipiravir | Umifenovir | NA | | NA | NA | 0.64 | -0.05 | 1.32 | Low |
| Favipiravir | Zanamivir | NA | | NA | NA | 0.22 | -0.34 | 0.77 | Moderate |
| Laninamivir | Oseltamivir | NA | | NA | NA | 0.18 | -0.24 | 0.59 | Moderate |
| Laninamivir | Oseltamivir plus Zanamivir | NA | | NA | NA | -1.93 | -3.4 | -0.46 | Low |
| Laninamivir | Peramivir | NA | | NA | NA | 0.38 | -0.13 | 0.88 | Moderate |
| Laninamivir | Standard care/placebo | NA | | NA | NA | -0.57 | -1.01 | -0.14 | Low |
| Laninamivir | Umifenovir | NA | | NA | NA | 0.52 | -0.09 | 1.14 | Low |
| Laninamivir | Zanamivir | NA | | NA | NA | 0.11 | -0.38 | 0.59 | Moderate |
| Oseltamivir | Oseltamivir plus Zanamivir | NA | | NA | NA | -2.67 | -4.12 | -1.22 | Moderate |
| Oseltamivir | Peramivir | NA | | NA | NA | 0.2* | -0.12* | 0.52* | Moderate |
| Oseltamivir | Standard care/placebo | NA | | NA | NA | -0.75 | -0.93 | -0.57 | Moderate |
| Oseltamivir | Umifenovir | NA | | NA | NA | 0.35 | -0.12 | 0.82 | Moderate |
| Oseltamivir | Zanamivir | NA | | NA | NA | -0.07 | -0.37 | 0.23 | Moderate |
| Oseltamivir plus Zanamivir | Peramivir | NA | | NA | NA | 2.31 | 0.86 | 3.76 | Low |
| Oseltamivir plus Zanamivir | Standard care/placebo | NA | | NA | NA | 1.36 | -0.07 | 2.78 | Low |
| Oseltamivir plus Zanamivir | Umifenovir | NA | | NA | NA | 2.46 | 0.97 | 3.94 | Low |
| Oseltamivir plus Zanamivir | Zanamivir | NA | | NA | NA | -0.16* | -2.01* | 1.69* | V low |
| Peramivir | Standard care/placebo | NA | | NA | NA | -0.95 | -1.28 | -0.62 | Low |
| Peramivir | Umifenovir | NA | | NA | NA | 0.15 | -0.41 | 0.7 | Moderate |
| Peramivir | Zanamivir | NA | | NA | NA | -0.27 | -0.67 | 0.13 | Moderate |
| Umifenovir | Standard care/placebo | NA | | NA | NA | -1.1 | -1.57 | -0.63 | Low |
| Zanamivir | Standard care/placebo | NA | | NA | NA | -0.68 | -0.93 | -0.43 | Moderate |
| Umifenovir | Zanamivir | NA | | NA | NA | -0.42 | -0.95 | 0.11 | Moderate |

NA, not applicable. *Estimate was informed by direct evidence because of serious incoherence in these comparisons.

#### 10.5. Network estimates with GRADE ratings for duration of hospitalization

| **Comparison groups** | | **Network relative estimate** | | | **Network absolute estimate (days)** | | | **GRADE rating** |
| --- | --- | --- | --- | --- | --- | --- | --- | --- |
| **Treatment 1** | **Treatment 2** | **Point estimate** | **95% CI lower limit** | **95% CI upper limit** | **Point estimate** | **95% CI lower limit** | **95% CI upper limit** |  |
| Oseltamivir | Peramivir | NA | NA | NA | 0.69 | 0.35 | 1.03 | Low |
| Oseltamivir | Standard care/placebo | NA | NA | NA | -0.35 | -3.19 | 2.49 | Very low |
| Oseltamivir | Zanamivir | NA | NA | NA | 0.75 | -2.33 | 3.83 | Very low |
| Peramivir | Standard care/placebo | NA | NA | NA | -1.04 | -3.90 | 1.82 | Very low |
| Peramivir | Zanamivir | NA | NA | NA | 0.06 | -3.04 | 3.16 | Very low |
| Zanamivir | Standard care/placebo | NA | NA | NA | -1.10 | -2.30 | 0.10 | Very low |

NA, not applicable.

#### 10.6. Network estimates with GRADE ratings for any adverse events

| **Comparison groups** | | **Network relative estimate (RR)** | | | **Network absolute estimate (per 1000)** | | | **GRADE rating** |
| --- | --- | --- | --- | --- | --- | --- | --- | --- |
| **Treatment 1** | **Treatment 2** | **Point estimate** | **95% CI lower limit** | **95% CI upper limit** | **Point estimate** | **95% CI lower limit** | **95% CI upper limit** |  |
| Baloxavir | Favipiravir | 0.88 | 0.72 | 1.07 | -40 | -93 | 23 | Very low |
| Baloxavir | Laninamivir | 0.95 | 0.04 | 22.84 | -15 | -296 | 692 | Very low |
| Baloxavir | Oseltamivir | 0.87 | 0.78 | 0.97 | -44 | -74 | -10 | Moderate |
| Baloxavir | Peramivir | 0.88 | 0.76 | 1.01 | -40 | -80 | 3 | Low |
| Baloxavir | Standard care/placebo | 0.85 | 0.75 | 0.95 | -52 | -87 | -17 | High |
| Baloxavir | Umifenovir | 1.26 | 0.50 | 3.18 | 60 | -116 | 505 | Very low |
| Baloxavir | Zanamivir | 0.92 | 0.81 | 1.06 | -25 | -60 | 19 | Very low |
| Favipiravir | Laninamivir | 1.09 | 0.05 | 26.09 | 28 | -293 | 692 | Very low |
| Favipiravir | Oseltamivir | 0.99 | 0.83 | 1.18 | -3 | -57 | 60 | Very low |
| Favipiravir | Peramivir | 1.00 | 0.83 | 1.20 | 0 | -56 | 66 | Very low |
| Favipiravir | Standard care/placebo | 0.96 | 0.82 | 1.13 | -14 | -62 | 45 | Very low |
| Favipiravir | Umifenovir | 1.44 | 0.57 | 3.66 | 102 | -100 | 617 | Very low |
| Favipiravir | Zanamivir | 1.05 | 0.88 | 1.26 | 16 | -38 | 83 | Very low |
| Laninamivir | Oseltamivir | 0.91 | 0.04 | 21.80 | -30 | -322 | 664 | Very low |
| Laninamivir | Peramivir | 0.92 | 0.04 | 21.98 | -27 | -319 | 668 | Very low |
| Laninamivir | Standard care/placebo | 0.89 | 0.04 | 21.20 | -38 | -332 | 654 | Very low |
| Laninamivir | Umifenovir | 1.32 | 0.05 | 36.05 | 74 | -220 | 768 | Very low |
| Laninamivir | Zanamivir | 0.97 | 0.04 | 23.18 | -10 | -306 | 682 | Very low |
| Oseltamivir | Peramivir | 1.01 | 0.92 | 1.10 | 3 | -27 | 33 | Very low |
| Oseltamivir | Standard care/placebo | 0.97 | 0.92 | 1.03 | -10 | -28 | 10 | Very low |
| Oseltamivir | Umifenovir | 1.45 | 0.58 | 3.64 | 104 | -97 | 612 | Very low |
| Oseltamivir | Zanamivir | 1.06 | 0.98 | 1.16 | 19 | -6 | 51 | Low |
| Peramivir | Standard care/placebo | 0.96 | 0.89 | 1.05 | -14 | -38 | 17 | Very low |
| Peramivir | Umifenovir | 1.44 | 0.57 | 3.62 | 102 | -100 | 607 | Very low |
| Peramivir | Zanamivir | 1.05 | 0.95 | 1.17 | 16 | -16 | 54 | Very low |
| Umifenovir | Standard care/placebo | 0.67 | 0.27 | 1.68 | -114 | -253 | 235 | Very low |
| Zanamivir | Standard care/placebo | 0.92 | 0.86 | 0.98 | -28 | -48 | -7 | Low |
| Umifenovir | Zanamivir | 0.73 | 0.29 | 1.84 | -86 | -226 | 267 | Very low |

#### 10.7. Network estimates with GRADE ratings for adverse events related to treatments

| **Comparison groups** | | **Network relative estimate (RR)** | | | **Network absolute estimate (per 1000)** | | | **GRADE rating** |
| --- | --- | --- | --- | --- | --- | --- | --- | --- |
| **Treatment 1** | **Treatment 2** | **Point estimate** | **95% CI lower limit** | **95% CI upper limit** | **Point estimate** | **95% CI lower limit** | **95% CI upper limit** |  |
| Baloxavir | Favipiravir | 0.83 | 0.56 | 1.23 | -18 | -48 | 25 | Very low |
| Baloxavir | Oseltamivir | 0.60 | 0.46 | 0.77 | -60 | -81 | -35 | High |
| Baloxavir | Oseltamivir plus Zanamivir | 1.79 | 0.21 | 15.09 | 40 | -40 | 705 | Very low |
| Baloxavir | Peramivir | 0.74 | 0.51 | 1.07 | -31 | -59 | 8 | Low |
| Baloxavir | Standard care/placebo | 0.74 | 0.57 | 0.95 | -32 | -52 | -6 | High |
| Baloxavir | Umifenovir | 1.30 | 0.52 | 3.25 | 21 | -33 | 156 | Very low |
| Baloxavir | Zanamivir | 0.70 | 0.52 | 0.95 | -38 | -61 | -6 | Low |
| Favipiravir | Oseltamivir | 0.72 | 0.52 | 0.99 | -42 | -72 | -2 | Low |
| Favipiravir | Oseltamivir plus Zanamivir | 2.15 | 0.25 | 18.33 | 58 | -38 | 867 | Very low |
| Favipiravir | Peramivir | 0.89 | 0.59 | 1.35 | -13 | -50 | 42 | Very low |
| Favipiravir | Standard care/placebo | 0.89 | 0.66 | 1.20 | -13 | -41 | 24 | Low |
| Favipiravir | Umifenovir | 1.56 | 0.62 | 3.97 | 39 | -26 | 207 | Very low |
| Favipiravir | Zanamivir | 0.85 | 0.61 | 1.18 | -19 | -50 | 23 | Very low |
| Oseltamivir | Oseltamivir plus Zanamivir | 3.00 | 0.36 | 24.92 | 100 | -32 | 950 | Very low |
| Oseltamivir | Peramivir | 1.24 | 0.96 | 1.62 | 29 | -5 | 75 | Low |
| Oseltamivir | Standard care/placebo | 1.23 | 1.10 | 1.39 | 28 | 12 | 48 | Moderate |
| Oseltamivir | Umifenovir | 2.18 | 0.89 | 5.30 | 82 | -8 | 299 | Very low |
| Oseltamivir | Zanamivir | 1.18 | 0.98 | 1.43 | 23 | -3 | 55 | Low |
| Oseltamivir plus Zanamivir | Peramivir | 0.41 | 0.05 | 3.50 | -71 | -115 | 302 | Very low |
| Oseltamivir plus Zanamivir | Standard care/placebo | 0.41 | 0.05 | 3.43 | -72 | -116 | 296 | Very low |
| Oseltamivir plus Zanamivir | Umifenovir | 0.73 | 0.07 | 7.21 | -19 | -65 | 432 | Very low |
| Oseltamivir plus Zanamivir | Zanamivir | 0.39 | 0.05 | 3.30 | -78 | -122 | 295 | Very low |
| Peramivir | Standard care/placebo | 0.99 | 0.74 | 1.32 | -1 | -32 | 39 | Low |
| Peramivir | Umifenovir | 1.75 | 0.69 | 4.42 | 52 | -22 | 238 | Very low |
| Peramivir | Zanamivir | 0.95 | 0.69 | 1.31 | -6 | -40 | 40 | Very low |
| Umifenovir | Standard care/placebo | 0.57 | 0.23 | 1.37 | -52 | -94 | 45 | Very low |
| Zanamivir | Standard care/placebo | 1.05 | 0.90 | 1.21 | 6 | -12 | 26 | Low |
| Umifenovir | Zanamivir | 0.54 | 0.22 | 1.33 | -59 | -100 | 42 | Very low |

#### 10.8. Network estimates with GRADE ratings for serious adverse events

| **Comparison groups** | | **Network relative estimate** | | | **Network absolute estimate (per 1000)** | | | **GRADE rating** |
| --- | --- | --- | --- | --- | --- | --- | --- | --- |
| **Treatment 1** | **Treatment 2** | **Point estimate** | **95% CI lower limit** | **95% CI upper limit** | **Point estimate** | **95% CI lower limit** | **95% CI upper limit** |  |
| Baloxavir | Favipiravir | NA | NA | NA | 2 | -5 | 10 | Very low |
| Baloxavir | Laninamivir | NA | NA | NA | 5 | -4 | 13 | Low |
| Baloxavir | Oseltamivir | NA | NA | NA | 1 | -4 | 5 | Moderate |
| Baloxavir | Oseltamivir plus Zanamivir | NA | NA | NA | -15 | -36 | 5 | Very low |
| Baloxavir | Peramivir | NA | NA | NA | 1 | -7 | 8 | Very low |
| Baloxavir | Standard care/placebo | NA | NA | NA | 1 | -4 | 5 | Moderate |
| Baloxavir | Zanamivir | NA | NA | NA | -2 | -7 | 4 | Low |
| Favipiravir | Laninamivir | NA | NA | NA | 2 | -7 | 11 | Very low |
| Favipiravir | Oseltamivir | NA | NA | NA | -2 | -8 | 5 | Very low |
| Favipiravir | Oseltamivir plus Zanamivir | NA | NA | NA | -18 | -39 | 4 | Low |
| Favipiravir | Peramivir | NA | NA | NA | -1 | -10 | 7 | Very low |
| Favipiravir | Standard care/placebo | NA | NA | NA | -2 | -7 | 4 | Moderate |
| Favipiravir | Zanamivir | NA | NA | NA | -4 | -11 | 3 | Low |
| Laninamivir | Oseltamivir | NA | NA | NA | -4 | -11 | 3 | Low |
| Laninamivir | Oseltamivir plus Zanamivir | NA | NA | NA | -20 | -41 | 2 | Low |
| Laninamivir | Peramivir | NA | NA | NA | -4 | -13 | 6 | Very low |
| Laninamivir | Standard care/placebo | NA | NA | NA | -4 | -11 | 3 | Moderate |
| Laninamivir | Zanamivir | NA | NA | NA | -6 | -14 | 2 | Low |
| Oseltamivir | Oseltamivir plus Zanamivir | NA | NA | NA | -16 | -36 | 4 | Low |
| Oseltamivir | Peramivir | NA | NA | NA | 0 | -6 | 7 | Very low |
| Oseltamivir | Standard care/placebo | NA | NA | NA | 0 | -3 | 2 | Moderate |
| Oseltamivir | Zanamivir | NA | NA | NA | -2 | -6 | 2 | Low |
| Oseltamivir plus Zanamivir | Peramivir | NA | NA | NA | 16 | -5 | 37 | Very low |
| Oseltamivir plus Zanamivir | Standard care/placebo | NA | NA | NA | 16 | -4 | 36 | Low |
| Oseltamivir plus Zanamivir | Zanamivir | NA | NA | NA | 14 | -7 | 34 | Very low |
| Peramivir | Standard care/placebo | NA | NA | NA | 0 | -7 | 6 | Low |
| Peramivir | Zanamivir | NA | NA | NA | -3 | -10 | 4 | Moderate |
| Zanamivir | Standard care/placebo | NA | NA | NA | 2 | -1 | 6 | Low |

NA, not applicable.

### Appendix 11. Summary of findings table for antivirals versus standard care or placebo

#### 11.1. Summary of findings table for favipiravir versus standard care or placebo

| **Outcome** | **Study results and measurements** | **Absolute effect estimates** | | **Certainty of the Evidence**  (Quality of evidence) | **Summary** |
| --- | --- | --- | --- | --- | --- |
|  |  | Standard care/placebo | Favipiravir |  |  |
| Mortality (Low-risk) | Relative risk: 0.57  (CI 95% 0.04 - 9.15)  Based on data from 1999 participants in 2 studies | **0.20**  per 1000 | **0.11**  per 1000 | **High** | Favipiravir has little or no effect on mortality. |
|  |  | Difference: **0.09 fewer per 1000**  (CI 95% 0.19 fewer - 1.63 more) | |  |  |
| Mortality (High-risk) | Relative risk: 0.57  (CI 95% 0.04 - 9.15)  Based on data from 1999 participants in 2 studies | **2.0**  per 1000 | **1.14**  per 1000 | **High** | Favipiravir has little or no effect on mortality. |
|  |  | Difference: **0.86 fewer per 1000**  (CI 95% 1.92 fewer - 16.3 more) | |  |  |
| Admission to hospital (Low-risk) | Relative risk: 0.57  (CI 95% 0.04 - 9.15)  Based on data from 1999 participants in 2 studies | **3**  per 1000 | **2**  per 1000 | **High** | Favipiravir has little or no effect on admission to hospital. |
|  |  | Difference: **1 fewer per 1000**  (CI 95% 3 fewer - 24 more) | |  |  |
| Admission to hospital (High-risk) | Relative risk: 0.57  (CI 95% 0.04 - 9.15)  Based on data from 1999 participants in 2 studies | **21**  per 1000 | **12**  per 1000 | **Very low**  Due to serious risk of bias, Due to very serious imprecision | Whether favipiravir reduces admission to hospital is very uncertain. |
|  |  | Difference: **9 fewer per 1000**  (CI 95% 20 fewer - 171 more) | |  |  |
| Adverse events related to treatments | Relative risk: 0.89  (CI 95% 0.66 - 1.2)  Based on data from 1999 participants in 2 studies | **122**  per 1000 | **109**  per 1000 | **Low**  Due to serious risk of bias, Due to serious imprecision | Favipiravir may not increase important adverse events related to treatments. |
|  |  | Difference: **13 fewer per 1000**  (CI 95% 41 fewer - 24 more) | |  |  |
| Serious adverse events | Risk difference: -0.002  (CI 95% -0.007 – 0.004)  Based on data from 2517 participants in 3 studies | **5**  per 1000 | **3**  per 1000 | **Moderate**  Due to serious risk of bias | Favipiravir probably has little or no effect on serious adverse events. |
|  |  | Difference: **2 fewer per 1000**  (CI 95% 7 fewer - 4 more) | |  |  |
| Time to alleviation of symptoms | Measured by: day  Lower better  Based on data from 1317 participants in 2 studies | **4.92**  Mean | **4.46**  Mean | **Moderate**  Due to serious risk of bias | Favipiravir probably has little or no effect on time to alleviation of symptoms. |
|  |  | Difference: **MD 0.46 lower**  (CI 95% 0.96 lower - 0.03 higher) | |  |  |

#### 11.2. Summary of findings table for laninamivir versus standard care or placebo

| **Outcome** | **Study results and measurements** | **Absolute effect estimates** | | **Certainty of the Evidence**  (Quality of evidence) | **Summary** |
| --- | --- | --- | --- | --- | --- |
|  |  | Standard care/placebo | Laninamivir |  |  |
| Mortality (Low-risk) | Relative risk: 0.50  (CI 95% 0.01 - 24.96)  Based on data from 639 participants in 1 study | **0.2**  per 1000 | **0.1**  per 1000 | **High** | Laninamivir has little or no effect on mortality. |
|  |  | Difference: **0.1 fewer per 1000**  (CI 95% 0.2 fewer - 4.79 more) | |  |  |
| Mortality (High-risk) | Relative risk: 0.50  (CI 95% 0.01 - 24.96)  Based on data from 639 participants in 1 study | **2.0**  per 1000 | **1.0**  per 1000 | **High** | Laninamivir has little or no effect on mortality. |
|  |  | Difference: **1 fewer per 1000**  (CI 95% 1.98 fewer - 47.92 more) | |  |  |
| Admission to hospital (Low-risk) | Relative risk: 1.10  (CI 95% 0.02 - 55.19)  Indirect evidence | **3**  per 1000 | **3**  per 1000 | **High** | Laninamivir has little or no effect on admission to hospital. |
|  |  | Difference: **0 fewer per 1000**  (CI 95% 3 fewer - 163 more) | |  |  |
| Admission to hospital (High-risk) | Relative risk: 1.10  (CI 95% 0.02 - 55.19)  Indirect evidence | **21**  per 1000 | **23**  per 1000 | **Low**  Due to serious risk of bias, Due to serious imprecision | Laninamivir may have little or no effect on admission to hospital. |
|  |  | Difference: **2 more per 1000**  (CI 95% 21 fewer - 979 more) | |  |  |
| Adverse events related to treatments (No data from RCT) |  |  |  |  | Whether laninamivir increases adverse events related to treatments is very uncertain. |
| Serious adverse events | Risk difference: -0.004  (CI 95% -0.011 – 0.003)  Based on data from 634 participants in 1 study | **5**  per 1000 | **1**  per 1000 | **Moderate**  Due to serious risk of bias | Laninamivir probably has little or no effect on serious adverse events. |
|  |  | Difference: **4 fewer per 1000**  (CI 95% 11 fewer - 3 more) | |  |  |
| Time to alleviation of symptoms | Measured by: day  Lower better  Based on data from 231 participants in 1 study | **4.92**  Mean | **4.35**  Mean | **Low**  Due to serious risk of bias, Due to serious imprecision | Laninamivir may have no important effect on time to alleviation of symptoms. |
|  |  | Difference: **MD 0.57 lower**  (CI 95% 1.01 lower - 0.14 lower) | |  |  |

#### 11.3. Summary of findings table for peramivir versus standard care or placebo

| **Outcome** | **Study results and measurements** | **Absolute effect estimates** | | **Certainty of the Evidence**  (Quality of evidence) | **Summary** |
| --- | --- | --- | --- | --- | --- |
|  |  | Standard care/placebo | Peramivir |  |  |
| Mortality (Low-risk) | Relative risk: 0.39  (CI 95% 0.04 - 4.04)  Based on data from 82 participants in 1 study | **0.2**  per 1000 | **0.08**  per 1000 | **High** | Peramivir has little or no effect on mortality. |
|  |  | Difference: **0.12 fewer per 1000**  (CI 95% 0.19 fewer - 0.61 more) | |  |  |
| Mortality (High-risk) | Relative risk: 0.39  (CI 95% 0.04 - 4.04)  Based on data from 82 participants in 1 study | **2.0**  per 1000 | **0.78**  per 1000 | **High** | Peramivir has little or no effect on mortality. |
|  |  | Difference: **1.22 fewer per 1000**  (CI 95% 1.92 fewer - 6.08 more) | |  |  |
| Admission to hospital (No data from RCT) |  |  |  |  | Whether peramivir reduces admission to hospital is very uncertain. |
| Adverse events related to treatments | Relative risk: 0.99  (CI 95% 0.74 - 1.32)  Indirect evidence | **122**  per 1000 | **121**  per 1000 | **Low**  Due to serious risk of bias, Due to serious imprecision | Peramivir may not increase adverse events related to treatments. |
|  |  | Difference: **1 fewer per 1000**  (CI 95% 32 fewer - 39 more) | |  |  |
| Serious adverse events | Risk difference: 0.0  (CI 95% -0.007 – 0.006)  Based on data from 1199 participants in 5 studies | **5**  per 1000 | **5**  per 1000 | **Low**  Due to serious risk of bias, Due to serious imprecision | Peramivir may not increase serious adverse events. |
|  |  | Difference: **0 fewer per 1000**  (CI 95% 7 fewer - 6 more) | |  |  |
| Time to alleviation of symptoms | Measured by: day  Lower better  Based on data from 1046 participants in 5 studies | **4.92**  Mean | **3.97**  Mean | **Low**  Due to serious risk of bias, Due to serious imprecision | Peramivir may not have an important reduction in time to alleviation of symptoms. |
|  |  | Difference: **MD 0.95 lower**  (CI 95% 1.28 lower - 0.62 lower) | |  |  |

#### 11.4. Summary of findings table for zanamivir versus standard care or placebo

| **Outcome** | **Study results and measurements** | **Absolute effect estimates** | | **Certainty of the Evidence**  (Quality of evidence) | **Summary** |
| --- | --- | --- | --- | --- | --- |
|  |  | Standard care/placebo | Zanamivir |  |  |
| Mortality (Low-risk) | Relative risk: 0.88  (CI 95% 0.34 - 2.28)  Based on data from 7174 participants in 16 studies | **0.2**  per 1000 | **0.18**  per 1000 | **High** | Zanamivir has little or no effect on mortality. |
|  |  | Difference: **0.02 fewer per 1000**  (CI 95% 0.13 fewer - 0.26 more) | |  |  |
| Mortality (High-risk) | Relative risk: 0.88  (CI 95% 0.34 - 2.28)  Based on data from 7174 participants in 16 studies | **2.0**  per 1000 | **1.76**  per 1000 | **High** | Zanamivir has little or no effect on mortality. |
|  |  | Difference: **0.24 fewer per 1000**  (CI 95% 1.32 fewer - 2.56 more) | |  |  |
| Admission to hospital (Low-risk) | Relative risk: 1.18  (CI 95% 0.81 - 1.72)  Based on data from 1160 participants in 3 studies | **3**  per 1000 | **4**  per 1000 | **High** | Zanamivir has little or no effect on admission to hospital. |
|  |  | Difference: **1 more per 1000**  (CI 95% 1 fewer - 2 more) | |  |  |
| Admission to hospital (High-risk) | Relative risk: 1.18  (CI 95% 0.81 - 1.72)  Based on data from 1160 participants in 3 studies | **21**  per 1000 | **25**  per 1000 | **High** | Zanamivir has little or no effect on admission to hospital. |
|  |  | Difference: **4 more per 1000**  (CI 95% 4 fewer - 15 more) | |  |  |
| Adverse events related to treatments | Relative risk: 1.05  (CI 95% 0.9 - 1.21)  Based on data from 7257 participants in 17 studies | **122**  per 1000 | **128**  per 1000 | **Low**  Due to serious risk of bias, Due to serious imprecision^2^ | Zanamivir may not increase adverse events related to treatments. |
|  |  | Difference: **6 more per 1000**  (CI 95% 12 fewer - 26 more) | |  |  |
| Serious adverse events | Risk difference: 0.002  (CI 95% -0.001 – 0.006)  Based on data from 7735 participants in 19 studies | **5**  per 1000 | **7**  per 1000 | **Low**  Due to serious risk of bias, Due to serious imprecision^3^ | Zanamivir may have little or no effect on serious adverse events. |
|  |  | Difference: **2 more per 1000**  (CI 95% 1 fewer - 6 more) | |  |  |
| Time to alleviation of symptoms | Measured by: day  Lower better  Based on data from 6617 participants in 15 studies | **4.92**  Mean | **4.24**  Mean | **Moderate**  Due to serious risk of bias^4^ | Zanamivir probably has no important effect on time to alleviation of symptoms. |
|  |  | Difference: **MD 0.68 lower**  (CI 95% 0.93 lower - 0.43 lower) | |  |  |

#### 11.5. Summary of findings table for umifenovir versus standard care or placebo

| **Outcome** | **Study results and measurements** | **Absolute effect estimates** | | **Certainty of the Evidence**  (Quality of evidence) | **Summary** |
| --- | --- | --- | --- | --- | --- |
|  |  | Standard care/placebo | Umifenovir |  |  |
| Mortality (Low-risk) | Relative risk: 0.85  (CI 95% 0.02 - 47.31)  Indirect evidence | **0.2**  per 1000 | **0.17**  per 1000 | **High** | Umifenovir has little or no effect on mortality. |
|  |  | Difference: **0.03 fewer per 1000**  (CI 95% 0.2 fewer - 9.26 more) | |  |  |
| Mortality (High-risk) | Relative risk: 0.85  (CI 95% 0.02 - 47.31)  Indirect evidence | **2.0**  per 1000 | **1.7**  per 1000 | **High** | Umifenovir has little or no effect on mortality. |
|  |  | Difference: **0.30 fewer per 1000**  (CI 95% 1.96 fewer - 92.62 more) | |  |  |
| Admission to hospital (Low-risk) | Relative risk: 0.27  (CI 95% 0.01 - 6.72)  Indirect evidence | **3**  per 1000 | **1**  per 1000 | **High** | Umifenovir has little or no effect on admission to hospital. |
|  |  | Difference: **2 fewer per 1000**  (CI 95% 3 fewer - 17 more) | |  |  |
| Admission to hospital (High-risk) | Relative risk: 0.27  (CI 95% 0.01 - 6.72)  Indirect evidence | **21**  per 1000 | **6**  per 1000 | **Very low**  Due to serious risk of bias, Due to very serious imprecision | Whether umifenovir reduces admission to hospital is very uncertain. |
|  |  | Difference: **15 fewer per 1000**  (CI 95% 21 fewer - 120 more) | |  |  |
| Adverse events related to treatments | Relative risk: 0.57  (CI 95% 0.23 - 1.37)  Based on data from 232 participants in 1 study | **122**  per 1000 | **70**  per 1000 | **Very low**  Due to serious risk of bias, Due to very serious imprecision^2^ | Whether umifenovir increases adverse events related to treatments is very uncertain. |
|  |  | Difference: **52 fewer per 1000**  (CI 95% 94 fewer - 45 more) | |  |  |
| Serious adverse events (No data from RCT) |  |  |  |  | Whether umifenovir increases serious adverse events is very uncertain. |
| Time to alleviation of symptoms | Measured by: day  Lower better  Based on data from 210 participants in 1 study | **4.92**  Mean | **3.82**  Mean | **Low**  Due to serious risk of bias, Due to serious imprecision^3^ | Umifenovir may reduce time to alleviation of symptoms. |
|  |  | Difference: **MD 1.10 lower**  (CI 95% 1.57 lower - 0.63 lower) | |  |  |

### Appendix 12. Funnel plots

#### 12.1. Funnel plot of oseltamivir vs standard care/placebo for mortality

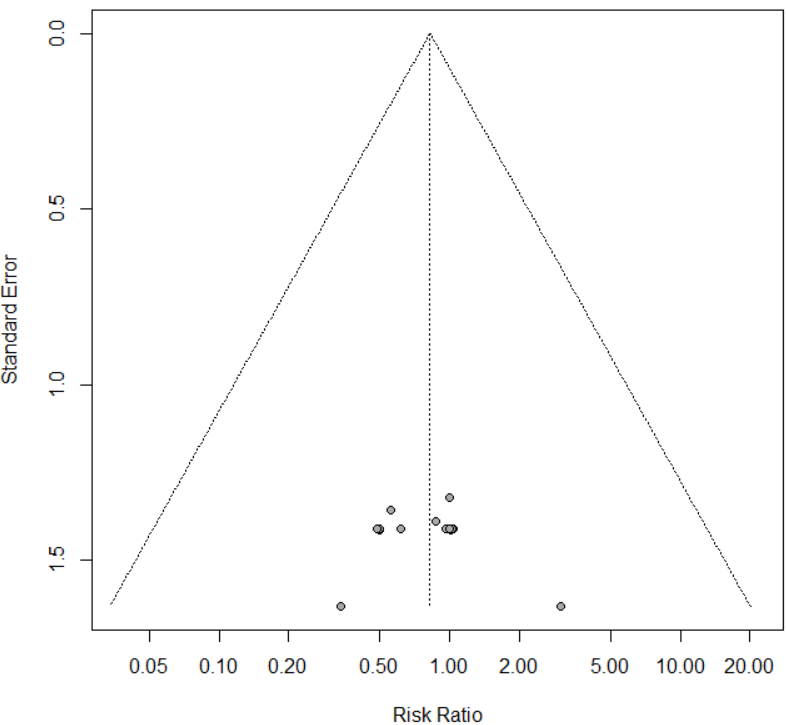

Harbord’s test p-value = 0.949

#### 12.2. Funnel plot of zanamivir vs standard care/placebo for mortality

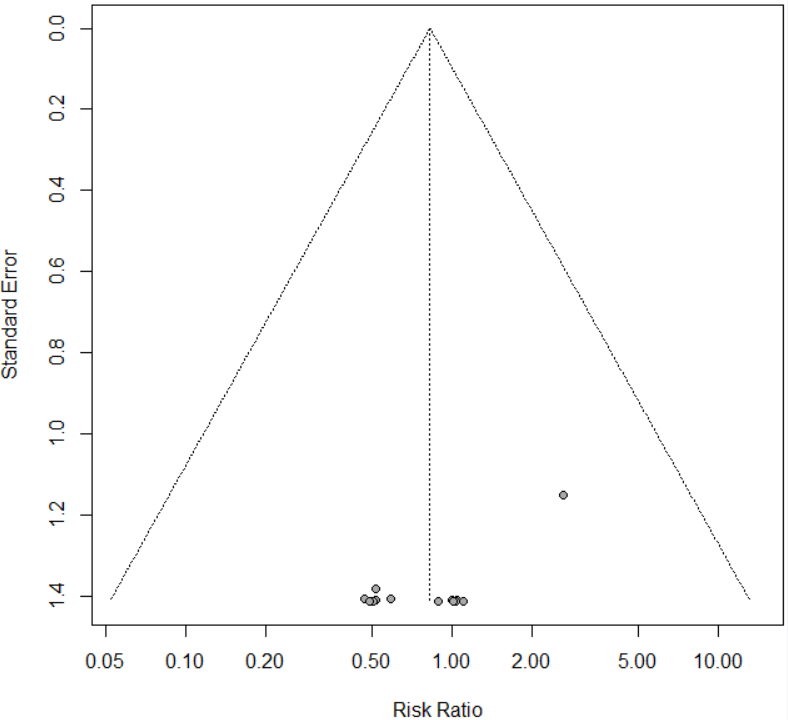

Harbord’s test p-value < 0.0001

#### 12.3. Filled Funnel plot of zanamivir vs standard care/placebo for mortality

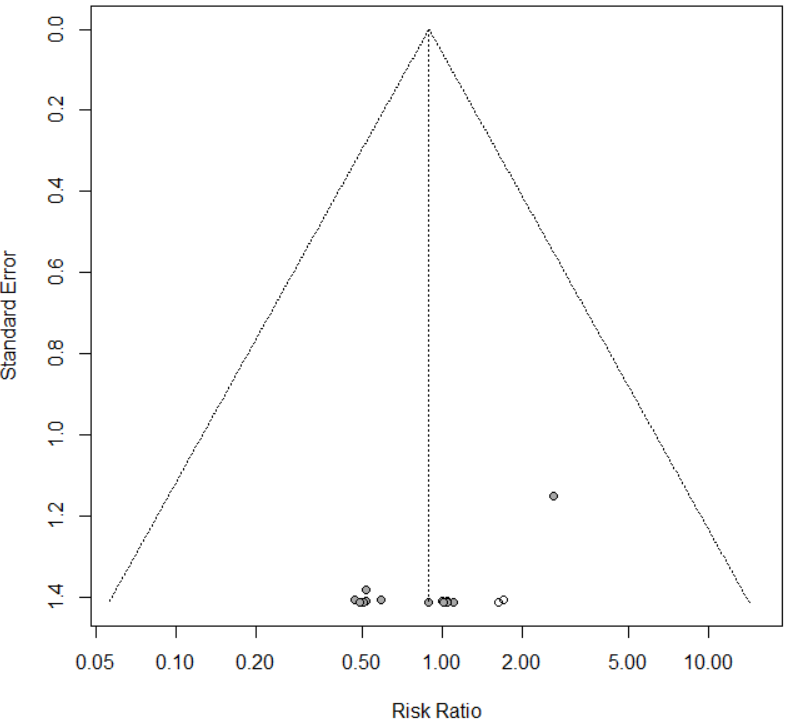

#### 12.4. Funnel plot of oseltamivir vs standard care/placebo for admission to hospital

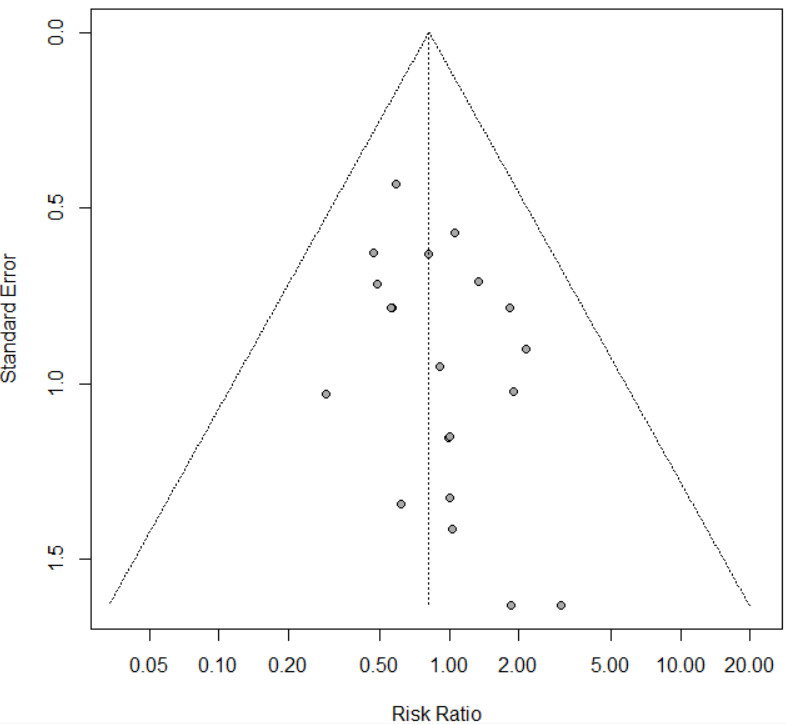

Harbord’s test p-value = 0.121

#### 12.5. Funnel plot of oseltamivir vs standard care/placebo for time to alleviation of symptoms

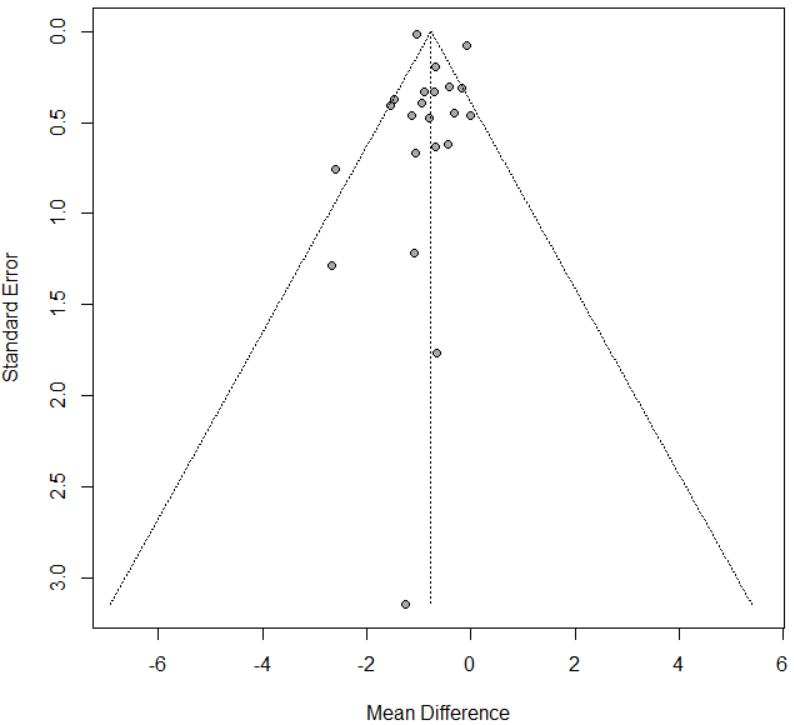

Egger’s test p-value = 0.216

#### 12.6. Funnel plot of zanamivir vs standard care/placebo for time to alleviation of symptoms

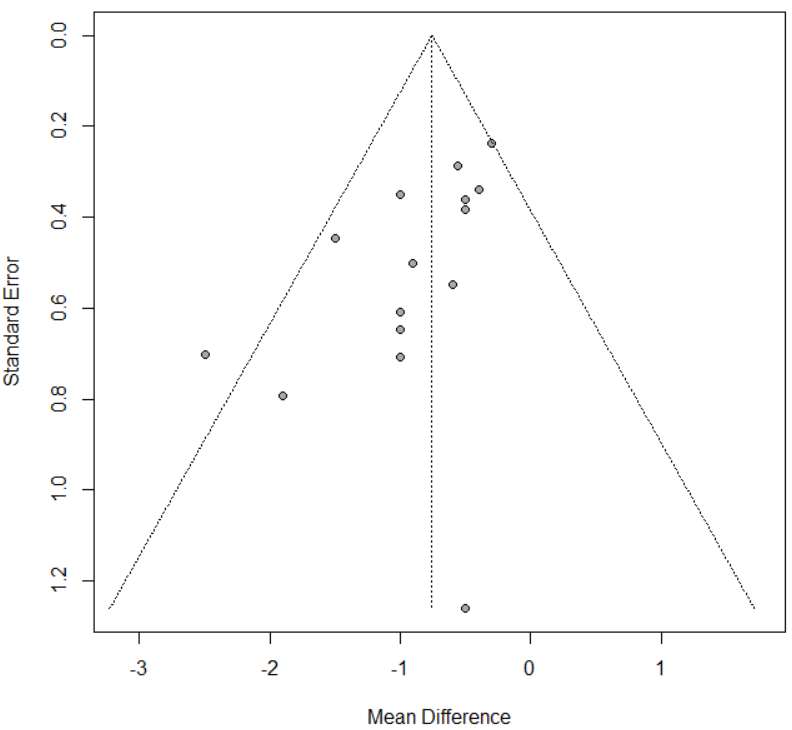

Egger’s test p-value = 0.011

#### 12.7. Filled Funnel plot of zanamivir vs standard care/placebo for time to alleviation of symptoms

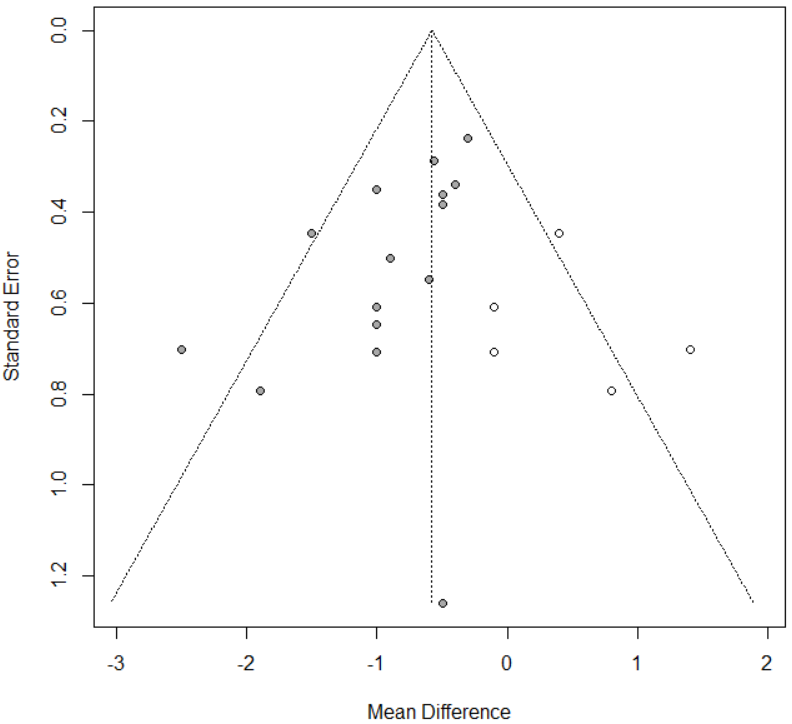

#### 12.8. Funnel plot of oseltamivir vs standard care/placebo for any adverse events

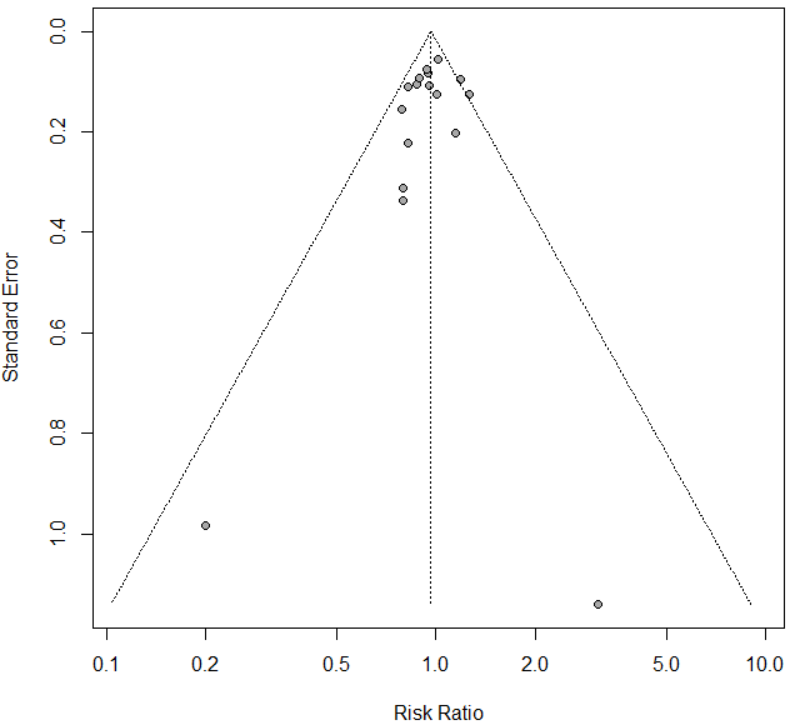

Harbord’s test p-value = 0.298

#### 12.9. Funnel plot of zanamivir vs standard care/placebo for any adverse events

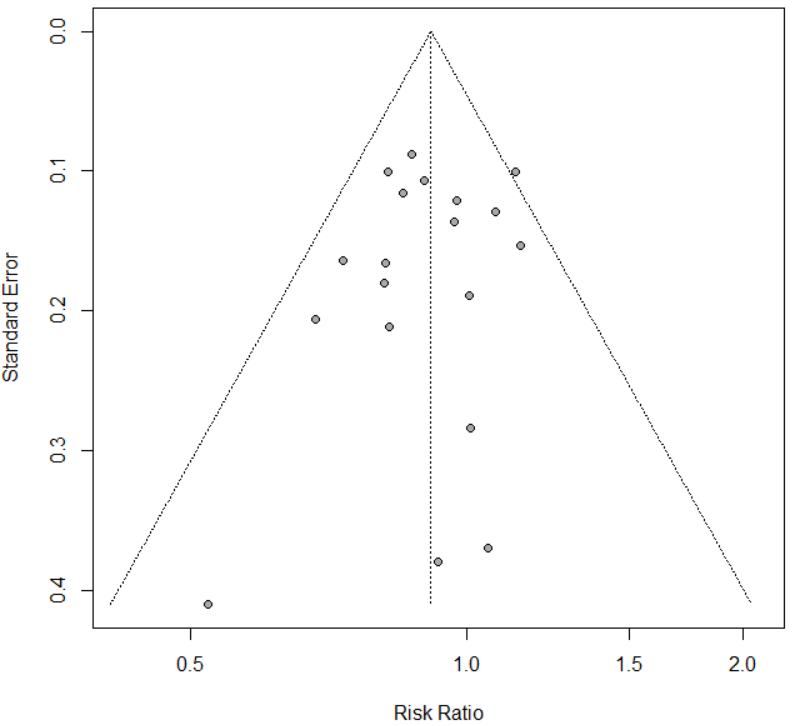

Harbord’s test p-value = 0.388

#### 12.10. Funnel plot of oseltamivir vs standard care/placebo for adverse events related to treatments

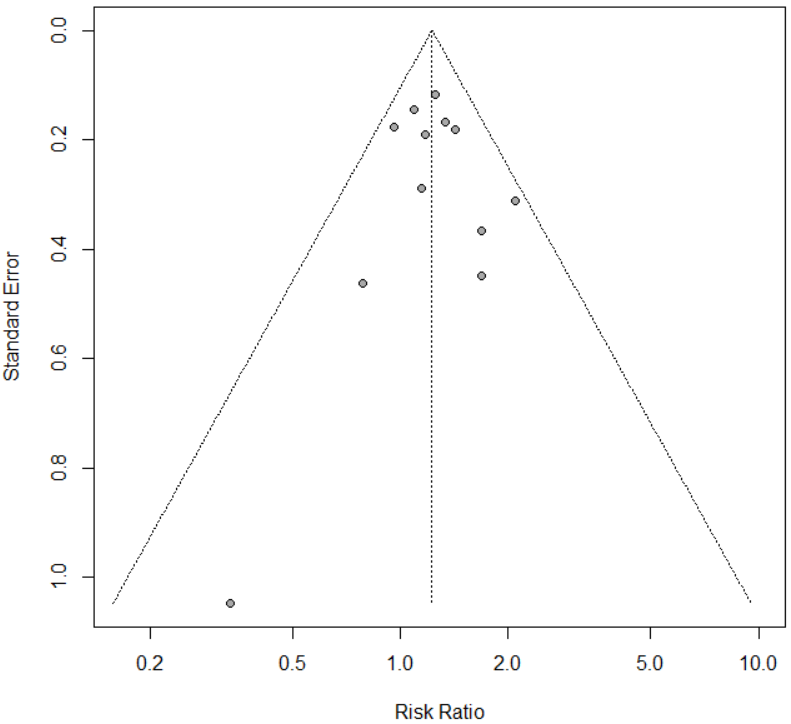

Harbord’s test p-value = 0.821

#### 12.11. Funnel plot of zanamivir vs standard care/placebo for adverse events related to treatments

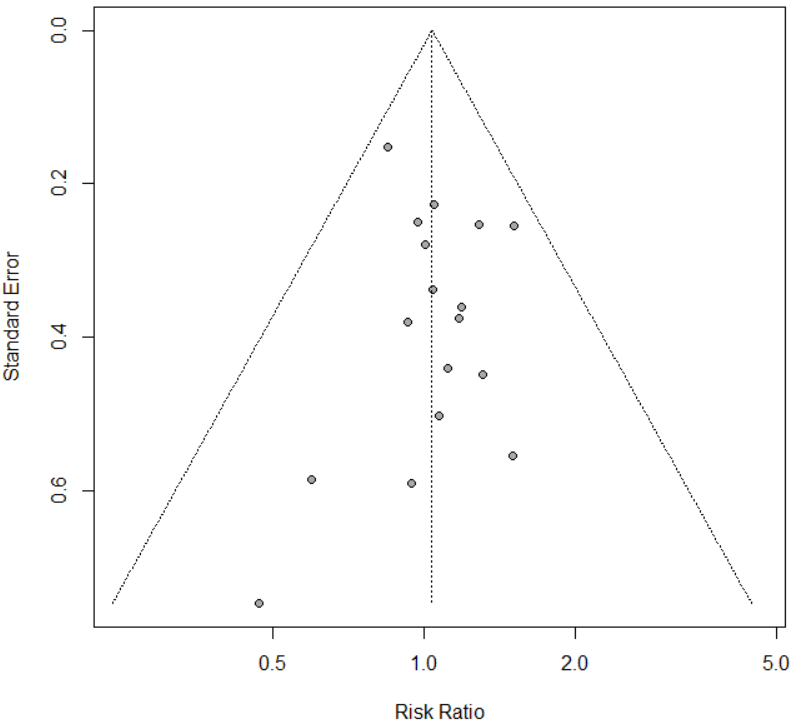

Harbord’s test p-value = 0.818

#### 12.12. Funnel plot of oseltamivir vs standard care/placebo for serious adverse events

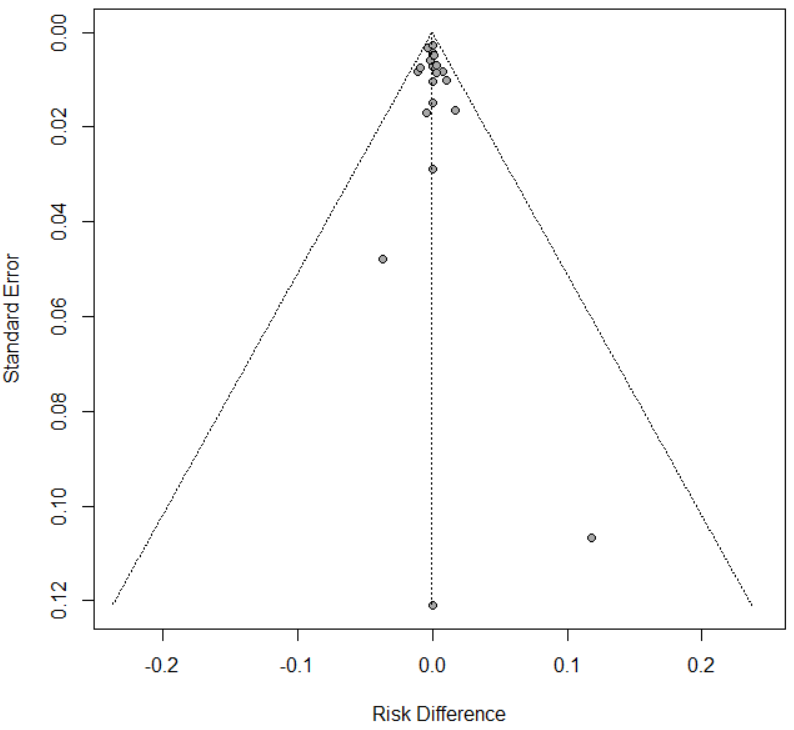

Harbord’s test p-value = 0.997

#### 12.13. Funnel plot of zanamivir vs standard care/placebo for serious adverse events

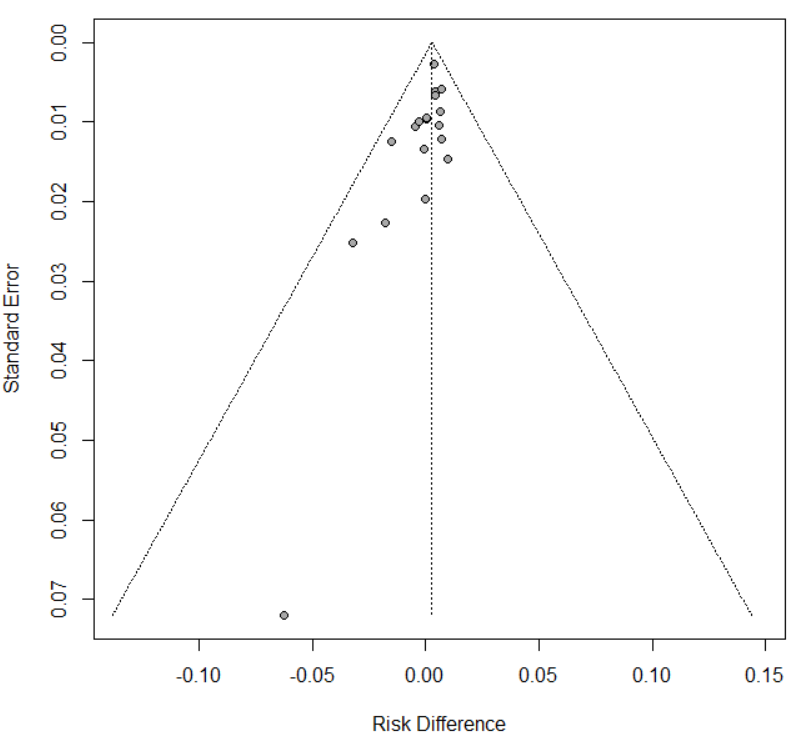

Harbord’s test p-value = 0.200

### Appendix 13. Results of for emergence of resistance

#### 13.1. Forest plot of meta-analysis for emergence of resistance

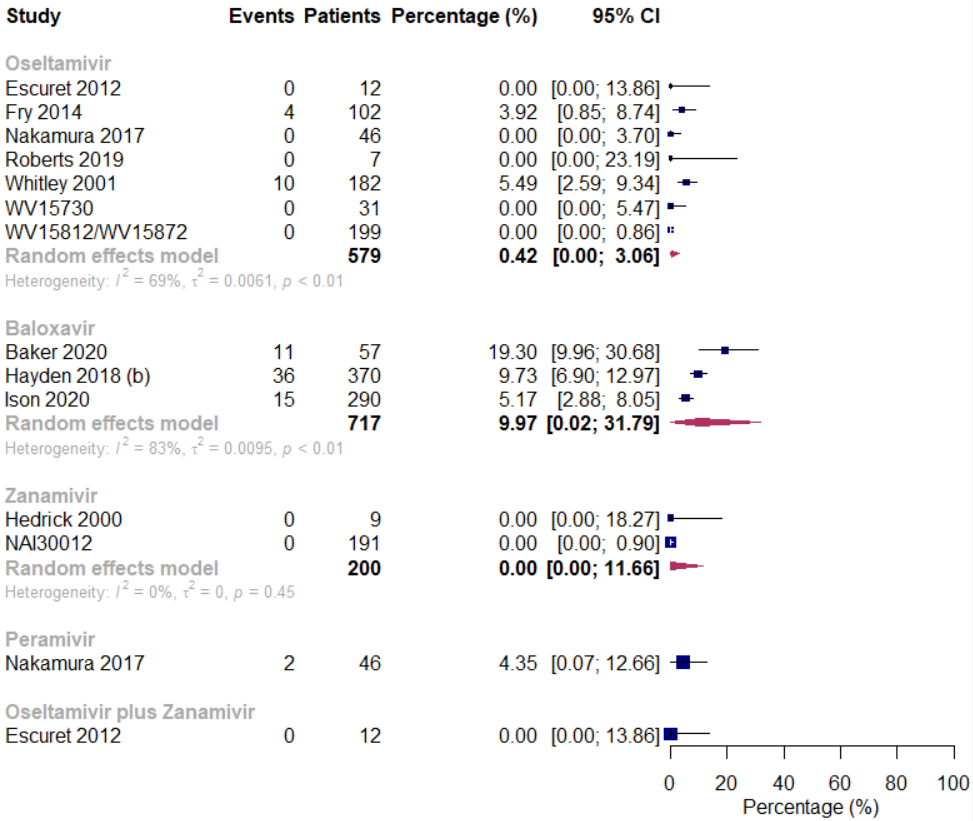

#### 13.2. GRADE assessment for emergence of resistance

| **Antivirals** | **№ of studies** | **Effect** | | | **Certainty of the evidence** |
| --- | --- | --- | --- | --- | --- |
|  |  | **№ of events** | **№ of individuals** | **Percentage**  **(95% CI)** |  |
| Baloxavir | 3 | 62 | 717 | pooled percentage  9.97 per 100 (0.02 to 31.79) | ⨁⨁◯◯Low^†‡^ |
| Oseltamivir | 7 | 14 | 579 | pooled percentage  0.42 per 100 (0.00 to 3.06) | ⨁◯◯◯  Very low^†#§^ |
| Peramivir | 1 | 2 | 46 | pooled percentage  4.35 per 100 (0.07 to 12.66) | ⨁◯◯◯  Very low^§^* |
| Zanamivir | 2 | 0 | 200 | pooled percentage  0 per 100 (0 to 11.66) | ⨁⨁◯◯  Low* |
| Oseltamivir plus Zanamivir | 1 | 0 | 12 | pooled percentage  0 per 100 (0 to 13.86) | ⨁⨁◯◯  Low* |

†Rated down one level for inconsistency due to the high heterogeneity.

‡Rated down one level for imprecision due to the wide 95% confidence interval.

#Rated down one level for imprecision due to the small sample size.

§Rated down one level for risk of bias.

*Rated down two levels for imprecision due to the wide 95% confidence interval and small sample size.

### Appendix 14. Within subgroup analysis

#### 14.1. Within subgroup analysis by type of influenza

Zanamivir versus standard care or placebo for time to alleviation of symptoms

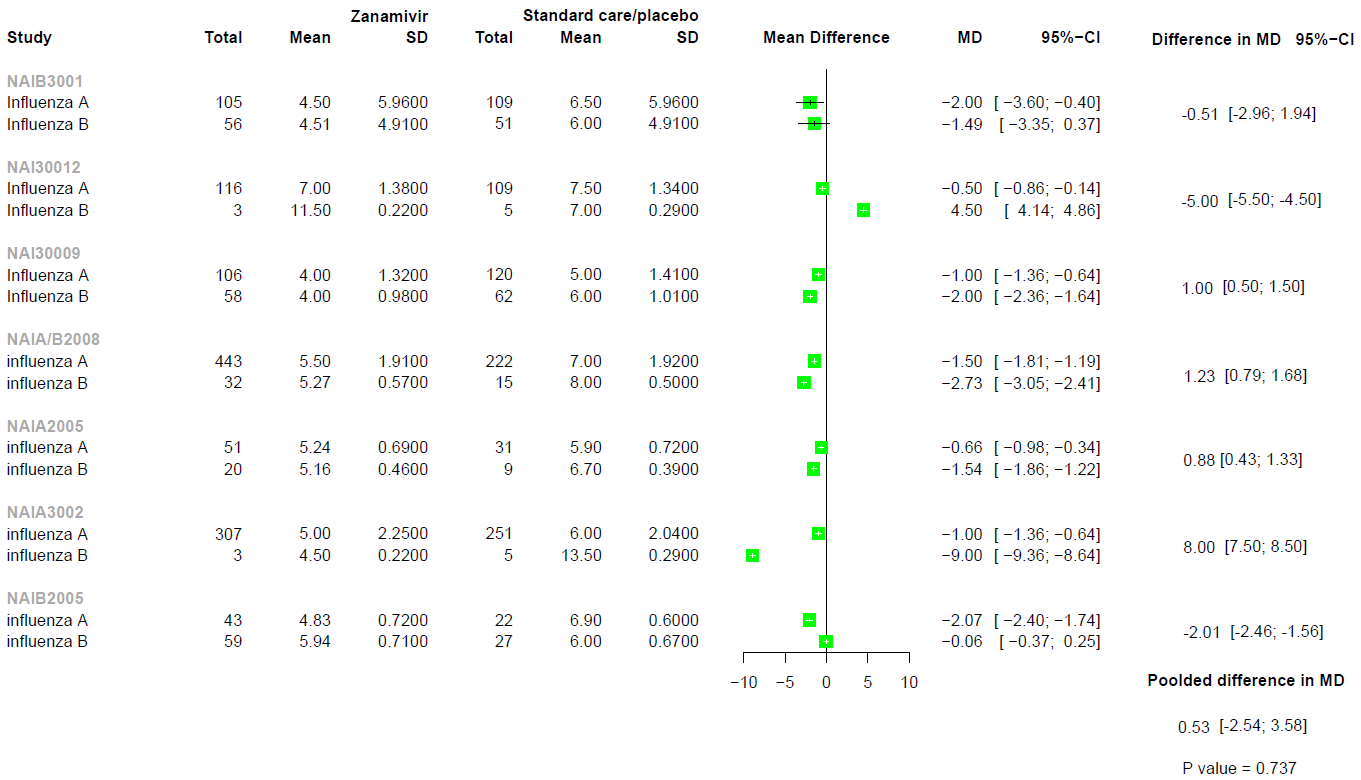

#### 14.2. Within subgroup analysis by confirmed or suspected influenza

Zanamivir versus standard care or placebo for time to alleviation of symptoms

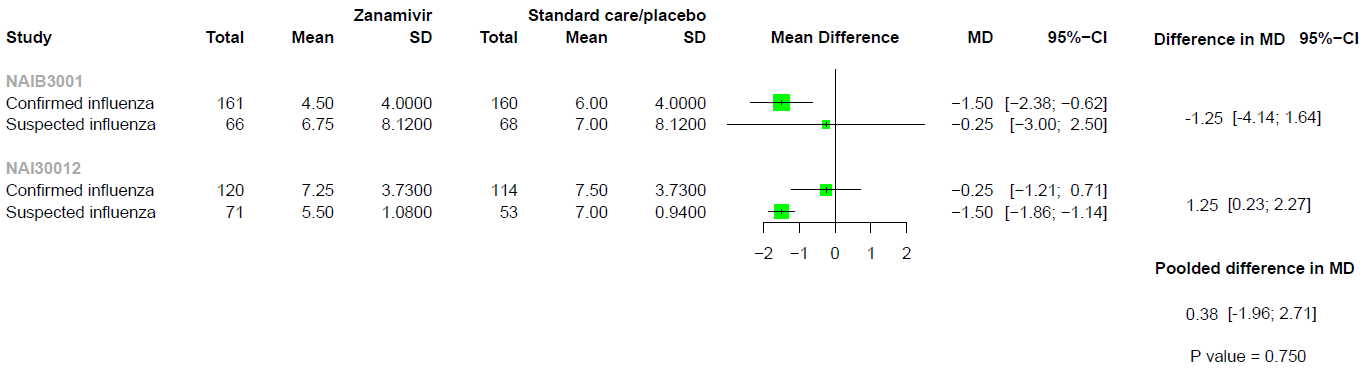

#### 14.3. Within subgroup analysis by age

Baloxavir versus standard care or placebo for mortality

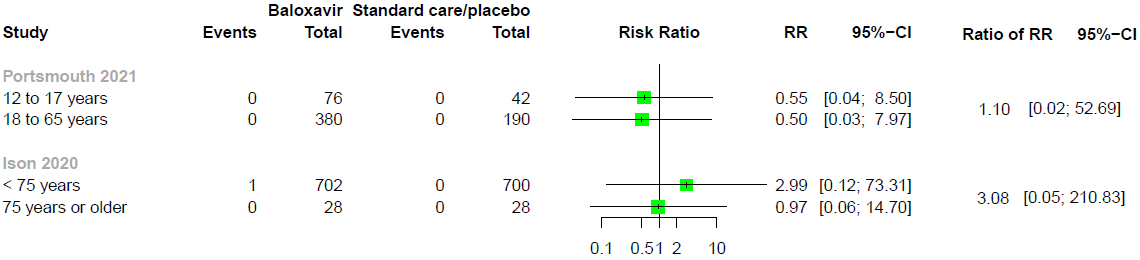

Baloxavir versus standard care or placebo for admission to hospital

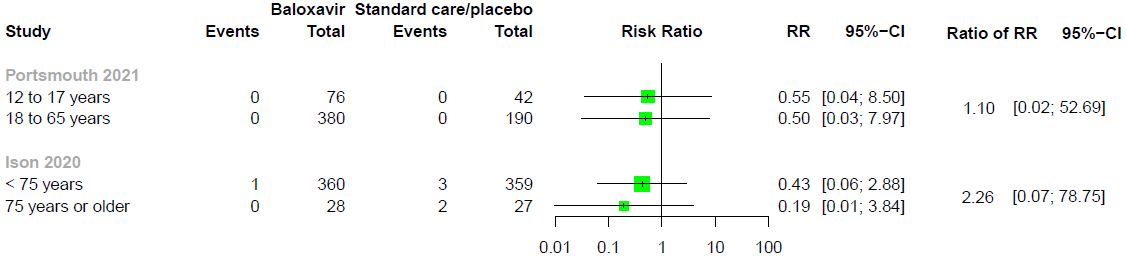

Baloxavir versus standard care or placebo for any adverse events

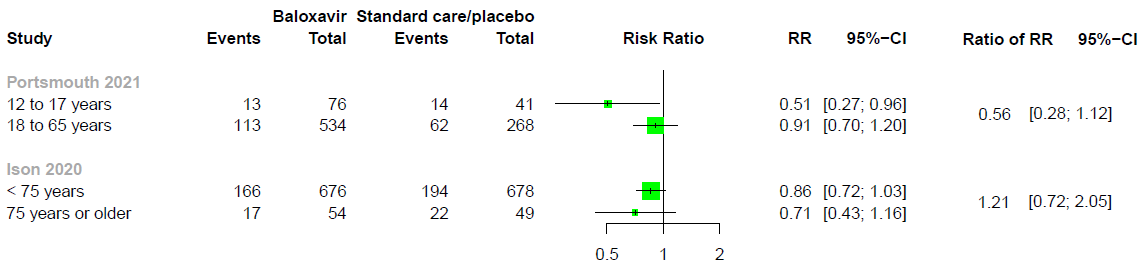

Baloxavir versus standard care or placebo for adverse events related to treatments

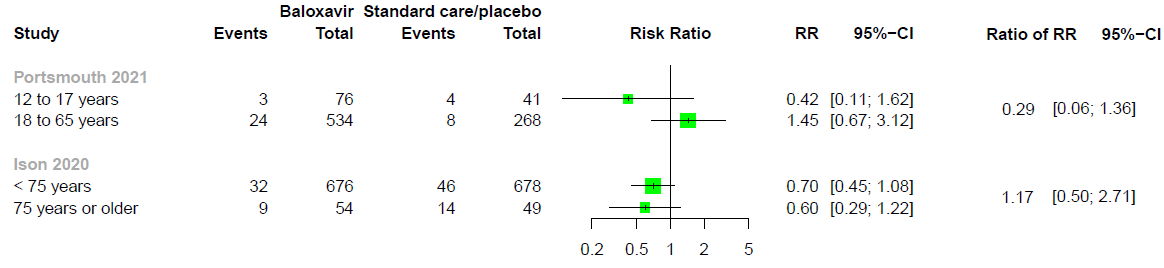

Baloxavir versus standard care or placebo for serious adverse events

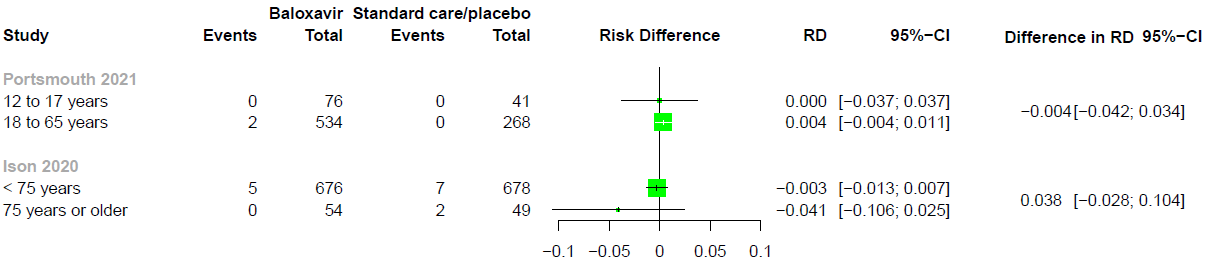

Favipiravir versus standard care or placebo for time to alleviation of symptoms

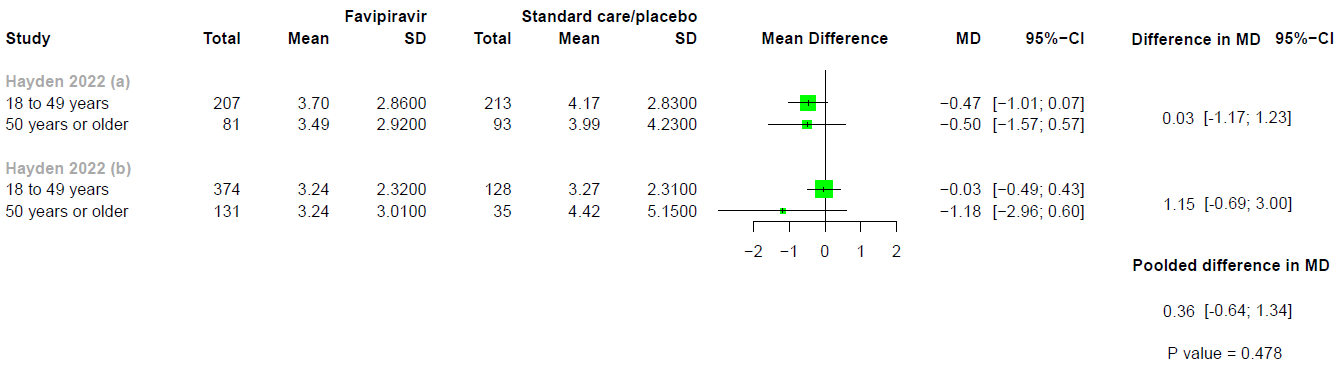

Oseltamivir versus standard care or placebo for serious adverse events

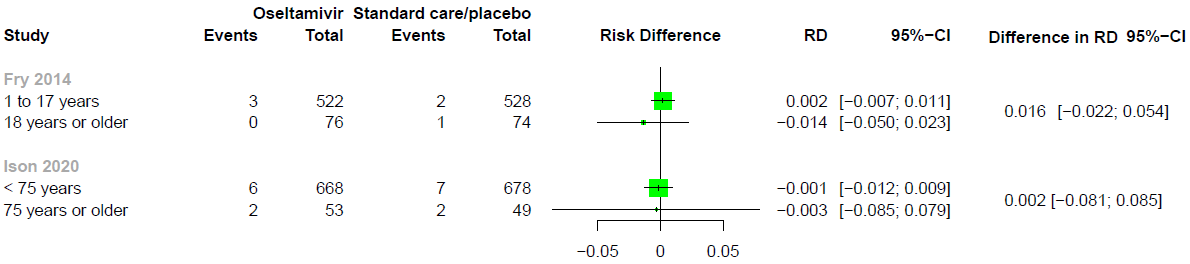

#### 14.4. Within subgroup analysis by risk of patients

Zanamivir versus standard care or placebo for any adverse events

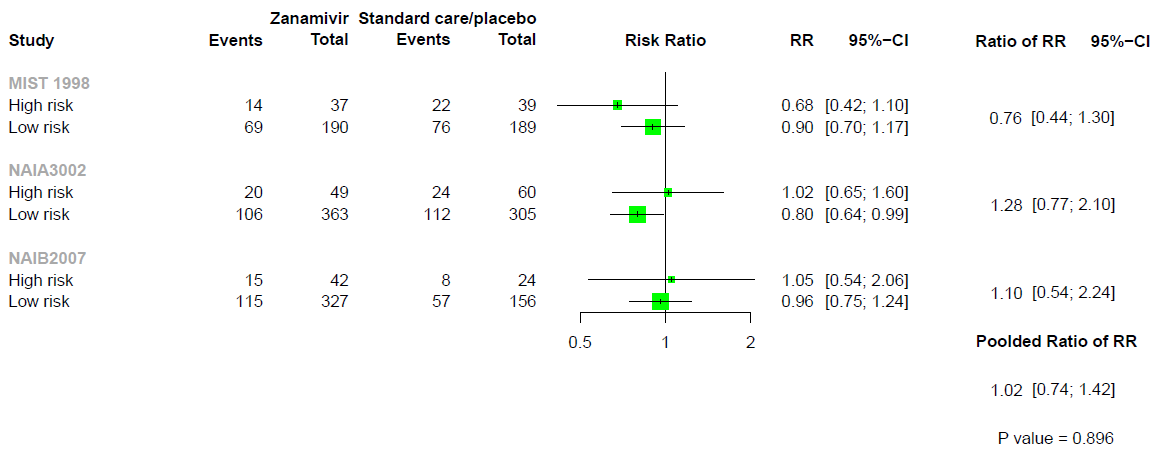

Zanamivir versus standard care or placebo for serious adverse events

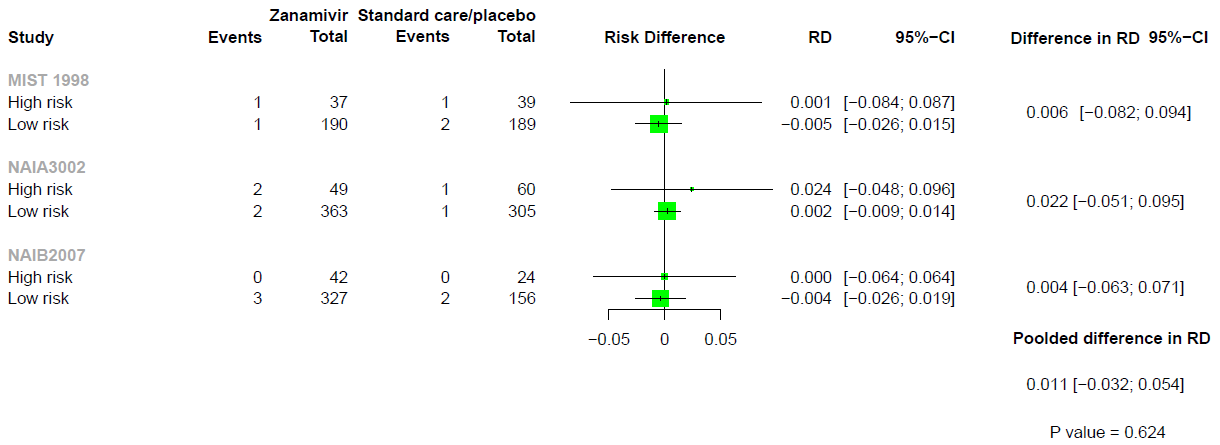

### Appendix 15. Network meta-regression results

| **Outcomes** | **Covariates** | B | 50% | 95%Crl |
| --- | --- | --- | --- | --- |
| Mortality | Mean age (years) | B | 1.12 | (-1.62, 6.53) |
|  | Proportion of Influenza A | B | 0.61 | (-3.09, 8.29) |
|  | Time from onset of symptoms to treatment/random (days) | B | 0.00 | (-7.79, 7.56) |
|  | Proportion of influenza vaccination | B | 0.85 | (-2.03, 6.71) |
| Admission to hospital | Mean age (years) | B | -0.80 | (-1.70, 0.04) |
|  | Proportion of Influenza A | B | 0.43 | (-0.68, 1.49) |
|  | Time from onset of symptoms to treatment/random (days) | B | -0.18 | (-2.87, 2.15) |
|  | Proportion of influenza vaccination | B | -0.28 | (-1.19, 0.66) |
| Time to alleviation of symptoms | Mean age (years) | B | 0.13 | (-0.23, 0.49) |
|  | Proportion of Influenza A | B | 0.30 | (-0.16, 0.77) |
|  | Time from onset of symptoms to treatment/random (days) | B | 0.19 | (-0.56, 1.02) |
|  | Proportion of influenza vaccination | B | 0.14 | (-0.47, 0.76) |
| Any adverse events | Mean age (years) | B | -0.01 | (-0.11, 0.09) |
|  | Proportion of Influenza A | B | -0.01 | (-0.12, 0.11) |
|  | Time from onset of symptoms to treatment/random (days) | B | 0.05 | (-0.25, 0.32) |
|  | Proportion of influenza vaccination | B | -0.06 | (-0.21, 0.08) |
| Adverse events related to treatments | Mean age (years) | B | -0.03 | (-0.27, 0.21) |
|  | Proportion of Influenza A | B | 0.10 | (-0.18, 0.37) |
|  | Time from onset of symptoms to treatment/random (days) | B | -0.17 | (-0.75, 0.39) |
|  | Proportion of influenza vaccination | B | -0.15 | (-0.46, 0.16) |

### Appendix 16. Sensitivity analysis using risk difference for mortality and admission to hospital

#### 16.1. Sensitivity analysis using risk difference for mortality

| **Baloxavir** | -0.000 (-0.003 to 0.003) | NA | NA | NA | NA | NA | 0.001 (-0.002 to 0.004) |
| --- | --- | --- | --- | --- | --- | --- | --- |
| 0.001 (-0.002 to 0.003) | **Oseltamivir** | NA | 0.000 (-0.020 to 0.020) | NA | NA | 0.000 (-0.009 to 0.009) | 0.000 (-0.001 to 0.001) |
| 0.001 (-0.007 to 0.008) | 0.000 (-0.007 to 0.007) | **Laninamivir** | NA | NA | NA | NA | 0.000 (-0.007 to 0.007) |
| 0.001 (-0.019 to 0.020) | 0.000 (-0.019 to 0.019) | -0.000 (-0.021 to 0.020) | **Peramivir** | NA | NA | NA | 0.000 (-0.058 to 0.058) |
| 0.000 (-0.003 to 0.004) | -0.000 (-0.002 to 0.002) | -0.000 (-0.008 to 0.007) | -0.000 (-0.019 to 0.019) | **Zanamivir** | NA | NA | 0.000 (-0.002 to 0.002) |
| 0.001 (-0.004 to 0.005) | 0.000 (-0.003 to 0.004) | -0.000 (-0.008 to 0.008) | 0.000 (-0.019 to 0.020) | 0.000 (-0.004 to 0.004) | **Favipiravir** | NA | 0.000 (-0.003 to 0.003) |
| 0.001 (-0.009 to 0.010) | 0.000 (-0.009 to 0.009) | -0.000 (-0.012 to 0.012) | -0.000 (-0.021 to 0.021) | 0.000 (-0.010 to 0.010) | -0.000 (-0.010 to 0.010) | **Umifenovir** | NA |
| 0.001 (-0.002 to 0.003) | 0.000 (-0.001 to 0.001) | -0.000 (-0.007 to 0.007) | 0.000 (-0.019 to 0.019) | 0.000 (-0.002 to 0.002) | 0.000 (-0.003 to 0.003) | 0.000 (-0.009 to 0.010) | **Standard care/placebo** |

Values are risk difference (95% CI) for network estimates (lower triangle) and direct estimates (upper triangle). Comparisons, column versus row, should be read from left, right. NA, not applicable.

#### 16.2. Sensitivity analysis using risk difference for admission to hospital

| **Baloxavir** | -0.004 (-0.010 to 0.002) | NA | NA | NA | NA | -0.002 (-0.008 to 0.004) |
| --- | --- | --- | --- | --- | --- | --- |
| -0.003 (-0.008 to 0.002) | **Oseltamivir** | NA | NA | NA | 0.005 (-0.008 to 0.018) | -0.000 (-0.003 to 0.002) |
| 0.003 (-0.047 to 0.052) | 0.006 (-0.044 to 0.055) | **Laninamivir** | 0.000 (-0.045 to 0.045) | NA | NA | NA |
| 0.003 (-0.018 to 0.024) | 0.006 (-0.015 to 0.026) | 0.000 (-0.045 to 0.045) | **Zanamivir** | NA | NA | -0.006 (-0.026 to 0.015) |
| -0.003 (-0.009 to 0.002) | -0.000 (-0.005 to 0.004) | -0.006 (-0.055 to 0.043) | -0.006 (-0.027 to 0.015) | **Favipiravir** | NA | 0.000 (-0.003 to 0.003) |
| 0.002 (-0.012 to 0.016) | 0.005 (-0.008 to 0.018) | -0.001 (-0.052 to 0.050) | -0.001 (-0.025 to 0.024) | 0.005 (-0.009 to 0.019) | **Umifenovir** | NA |
| -0.003 (-0.008 to 0.001) | -0.000 (-0.003 to 0.002) | -0.006 (-0.055 to 0.043) | -0.006 (-0.026 to 0.015) | -0.000 (-0.003 to 0.003) | -0.005 (-0.019 to 0.008) | **Standard care/placebo** |

Values are risk difference (95% CI) for network estimates (lower triangle) and direct estimates (upper triangle). Comparisons, column versus row, should be read from left, right. NA, not applicable.

### Appendix 17. Sensitivity analysis by only including studies with confirmed influenza patients

#### 17.1. Sensitivity analysis by only including studies with confirmed influenza patients for mortality

| **Baloxavir** | 0.94 (0.13 to 6.70) | NA | NA | NA | 1.47 (0.12 to 17.50) |
| --- | --- | --- | --- | --- | --- |
| 1.03 (0.16 to 6.62) | **Oseltamivir** | 1.13 (0.02 to 56.34) | NA | NA | 1.27 (0.24 to 6.69) |
| 1.75 (0.06 to 47.73) | 1.71 (0.10 to 30.18) | **Peramivir** | NA | NA | 0.44 (0.01 to 21.74) |
| 2.35 (0.07 to 75.34) | 2.29 (0.09 to 55.39) | 1.34 (0.02 to 71.95) | **Zanamivir** | NA | 0.50 (0.03 to 7.97) |
| 2.05 (0.06 to 65.95) | 2.00 (0.08 to 48.50) | 1.17 (0.02 to 62.96) | 0.87 (0.02 to 43.84) | **Favipiravir** | 0.57 (0.04 to 9.15) |
| 1.17 (0.14 to 9.51) | 1.14 (0.24 to 5.56) | 0.67 (0.04 to 11.78) | 0.50 (0.03 to 7.97) | 0.57 (0.04 to 9.15) | Standard care/placebo |

Values are risk ratio (95% CI) for network estimates (lower triangle) and direct estimates (upper triangle). Comparisons, column versus row, should be read from left, right. NA, not applicable.

#### 17.2. Sensitivity analysis by only including studies with confirmed influenza patients for admission to hospital

| **Baloxavir** | 0.26 (0.04 to 1.57) | NA | NA | NA | 0.25 (0.04 to 1.62) |
| --- | --- | --- | --- | --- | --- |
| 0.24 (0.04 to 1.41) | **Oseltamivir** | NA | NA | NA | 1.03 (0.51 to 2.09) |
| 0.16 (0.00 to 12.21) | 0.67 (0.01 to 36.51) | **Laninamivir** | 0.93 (0.02 to 45.94) | NA | NA |
| 0.15 (0.02 to 0.96) | 0.63 (0.26 to 1.51) | 0.93 (0.02 to 45.94) | **Zanamivir** | NA | 1.63 (0.97 to 2.74) |
| 0.44 (0.02 to 11.63) | 1.78 (0.10 to 31.14) | 2.65 (0.02 to 325.63) | 2.85 (0.17 to 47.68) | **Favipiravir** | 0.57 (0.04 to 9.15) |
| 0.25 (0.04 to 1.45) | 1.02 (0.50 to 2.07) | 1.52 (0.03 to 77.55) | 1.63 (0.97 to 2.74) | 0.57 (0.04 to 9.15) | **Standard care/placebo** |

Values are risk ratio (95% CI) for network estimates (lower triangle) and direct estimates (upper triangle). Comparisons, column versus row, should be read from left, right. NA, not applicable.

#### 17.3. Sensitivity analysis by only including studies with confirmed influenza patients for time to alleviation of symptoms

| **Baloxavir** | NA | NA | -0.27 (-0.96 to 0.43) | NA | NA | NA | NA | NA | -1.04 (-1.45 to -0.62) |
| --- | --- | --- | --- | --- | --- | --- | --- | --- | --- |
| -0.58 (-1.20 to 0.04) | **Favipiravir** | NA | NA | NA | NA | NA | NA | NA | -0.46 (-0.95 to 0.03) |
| -0.39 (-0.96 to 0.18) | 0.19 (-0.46 to 0.85) | **Laninamivir** | 0.03 (-0.44 to 0.50) | 0.24 (-0.89 to 1.37) | NA | -0.06 (-1.16 to 1.04) | NA | NA | -0.06 (-0.93 to 0.81) |
| -0.20 (-0.61 to 0.22) | 0.38 (-0.15 to 0.92) | 0.19 (-0.22 to 0.61) | **Oseltamivir** | 0.23 (-0.14 to 0.60) | NA | -0.73 (-1.47 to 0.00) | NA | -2.67 (-4.11 to -1.23) | -0.83 (-1.07 to -0.58) |
| -0.02 (-0.52 to 0.48) | 0.57 (-0.03 to 1.16) | 0.37 (-0.13 to 0.88) | 0.18 (-0.14 to 0.50) | **Peramivir** | NA | -0.30 (-1.17 to 0.57) | NA | NA | -0.98 (-1.52 to -0.44) |
| -0.04 (-0.85 to 0.76) | 0.54 (-0.32 to 1.39) | 0.35 (-0.48 to 1.17) | 0.15 (-0.58 to 0.89) | -0.03 (-0.81 to 0.75) | **Umifenovir** | NA | NA | NA | -1.00 (-1.70 to -0.30) |
| -0.16 (-0.60 to 0.29) | 0.43 (-0.11 to 0.96) | 0.23 (-0.24 to 0.71) | 0.04 (-0.25 to 0.33) | -0.14 (-0.53 to 0.25) | -0.11 (-0.85 to 0.62) | **Zanamivir** | NA | 0.16 (-1.69 to 2.01) | -0.97 (-1.21 to -0.74) |
| -0.26 (-0.91 to 0.38) | 0.32 (-0.39 to 1.03) | 0.13 (-0.55 to 0.81) | -0.07 (-0.63 to 0.50) | -0.25 (-0.87 to 0.37) | -0.22 (-1.09 to 0.66) | -0.11 (-0.67 to 0.46) | **Amantadine** | NA | -0.78 (-1.30 to -0.26) |
| -2.28 (-3.75 to -0.82) | -1.70 (-3.21 to -0.20) | -1.89 (-3.36 to -0.42) | -2.09 (-3.50 to -0.68) | -2.27 (-3.71 to -0.82) | -2.24 (-3.83 to -0.65) | -2.13 (-3.56 to -0.70) | -2.02 (-3.54 to -0.51) | **Oseltamivir plus Zanamivir** | NA |
| -1.04 (-1.43 to -0.66) | -0.46 (-0.95 to 0.03) | -0.65 (-1.09 to -0.22) | -0.85 (-1.06 to -0.63) | -1.03 (-1.37 to -0.69) | -1.00 (-1.70 to -0.30) | -0.89 (-1.11 to -0.67) | -0.78 (-1.30 to -0.26) | 1.24 (-0.18 to 2.66) | **Standard care/placebo** |

Values are mean difference (95% CI) for network estimates (lower triangle) and direct estimates (upper triangle). Comparisons, column versus row, should be read from left, right. NA, not applicable.

#### 17.4. Sensitivity analysis by only including studies with confirmed influenza patients for any adverse events

| **Baloxavir** | NA | NA | 0.89 (0.75 to 1.06) | NA | NA | NA | 0.85 (0.73 to 0.99) |
| --- | --- | --- | --- | --- | --- | --- | --- |
| 0.89 (0.72 to 1.10) | **Favipiravir** | NA | NA | NA | NA | NA | 0.96 (0.82 to 1.13) |
| 0.98 (0.04 to 23.45) | 1.10 (0.05 to 26.32) | **Laninamivir** | 1.00 (0.02 to 48.80) | 0.93 (0.02 to 45.55) | NA | 0.87 (0.02 to 42.29) | NA |
| 0.88 (0.76 to 1.03) | 0.99 (0.82 to 1.20) | 0.90 (0.04 to 21.60) | **Oseltamivir** | 1.04 (0.91 to 1.17) | 1.00 (0.02 to 49.43) | 0.87 (0.02 to 42.29) | 0.93 (0.79 to 1.09) |
| 0.89 (0.76 to 1.03) | 0.99 (0.84 to 1.18) | 0.91 (0.04 to 21.69) | 1.00 (0.91 to 1.11) | **Peramivir** | NA | 0.93 (0.02 to 45.21) | 0.98 (0.90 to 1.06) |
| 1.55 (0.50 to 4.83) | 1.74 (0.56 to 5.42) | 1.59 (0.05 to 46.10) | 1.76 (0.57 to 5.44) | 1.75 (0.57 to 5.41) | **Umifenovir** | NA | 0.53 (0.16 to 1.70) |
| 0.97 (0.77 to 1.23) | 1.09 (0.85 to 1.38) | 0.99 (0.04 to 23.76) | 1.10 (0.89 to 1.36) | 1.09 (0.90 to 1.33) | 0.62 (0.20 to 1.95) | **Zanamivir** | 0.89 (0.74 to 1.07) |
| 0.86 (0.74 to 0.99) | 0.96 (0.82 to 1.13) | 0.88 (0.04 to 21.01) | 0.97 (0.87 to 1.08) | 0.97 (0.90 to 1.04) | 0.55 (0.18 to 1.70) | 0.89 (0.74 to 1.07) | **Standard care/placebo** |

Values are risk ratio (95% CI) for network estimates (lower triangle) and direct estimates (upper triangle). Comparisons, column versus row, should be read from left, right. NA, not applicable.

#### 17.5. Sensitivity analysis by only including studies with confirmed influenza patients for adverse events related to treatments

| **Baloxavir** | NA | 0.71 (0.42 to 1.19) | NA | NA | NA | 0.70 (0.45 to 1.07) |
| --- | --- | --- | --- | --- | --- | --- |
| 0.79 (0.45 to 1.39) | **Favipiravir** | NA | NA | NA | NA | 0.89 (0.61 to 1.31) |
| 0.71 (0.45 to 1.12) | 0.90 (0.52 to 1.54) | **Oseltamivir** | 1.24 (0.81 to 1.91) | NA | NA | 0.99 (0.67 to 1.46) |
| 0.88 (0.47 to 1.65) | 1.12 (0.56 to 2.23) | 1.24 (0.81 to 1.91) | **Peramivir** | NA | NA | NA |
| 1.24 (0.44 to 3.48) | 1.58 (0.57 to 4.37) | 1.75 (0.63 to 4.86) | 1.41 (0.47 to 4.26) | **Umifenovir** | NA | 0.57 (0.22 to 1.46) |
| 0.71 (0.37 to 1.33) | 0.90 (0.49 to 1.66) | 1.00 (0.54 to 1.84) | 0.80 (0.38 to 1.69) | 0.57 (0.20 to 1.64) | **Zanamivir** | 1.00 (0.62 to 1.61) |
| 0.70 (0.46 to 1.07) | 0.89 (0.61 to 1.31) | 0.99 (0.68 to 1.46) | 0.80 (0.45 to 1.42) | 0.57 (0.22 to 1.46) | 1.00 (0.62 to 1.61) | **Standard care/placebo** |

Values are risk ratio (95% CI) for network estimates (lower triangle) and direct estimates (upper triangle). Comparisons, column versus row, should be read from left, right. NA, not applicable.

#### 17.6. Sensitivity analysis by only including studies with confirmed influenza patients for serious adverse events

| **Baloxavir** | NA | NA | -0.004 (-0.014 to 0.005) | NA | NA | NA | -0.004 (-0.012 to 0.004) |
| --- | --- | --- | --- | --- | --- | --- | --- |
| -0.002 (-0.011 to 0.007) | **Favipiravir** | NA | NA | NA | NA | NA | -0.002 (-0.007 to 0.004) |
| 0.003 (-0.009 to 0.014) | 0.004 (-0.007 to 0.016) | **Laninamivir** | -0.006 (-0.015 to 0.002) | NA | NA | NA | NA |
| -0.004 (-0.012 to 0.004) | -0.002 (-0.009 to 0.006) | -0.006 (-0.015 to 0.002) | **Oseltamivir** | -0.002 (-0.011 to 0.006) | 0.006 (-0.010 to 0.021) | -0.010 (-0.033 to 0.013) | 0.001 (-0.005 to 0.006) |
| -0.003 (-0.013 to 0.006) | -0.001 (-0.010 to 0.007) | -0.006 (-0.017 to 0.005) | 0.000 (-0.006 to 0.007) | **Peramivir** | NA | NA | -0.003 (-0.012 to 0.006) |
| -0.002 (-0.014 to 0.010) | -0.000 (-0.011 to 0.011) | -0.005 (-0.018 to 0.009) | 0.002 (-0.008 to 0.012) | 0.001 (-0.010 to 0.013) | **Zanamivir** | -0.016 (-0.036 to 0.005) | 0.000 (-0.011 to 0.012) |
| -0.017 (-0.039 to 0.006) | -0.015 (-0.037 to 0.007) | -0.019 (-0.042 to 0.004) | -0.013 (-0.034 to 0.009) | -0.013 (-0.035 to 0.009) | -0.015 (-0.035 to 0.006) | **Oseltamivir plus Zanamivir** | NA |
| -0.004 (-0.011 to 0.004) | -0.002 (-0.007 to 0.004) | -0.006 (-0.016 to 0.004) | 0.000 (-0.005 to 0.005) | -0.000 (-0.007 to 0.006) | -0.002 (-0.011 to 0.008) | 0.013 (-0.008 to 0.034) | **Standard care/placebo** |

Values are risk difference (95% CI) for network estimates (lower triangle) and direct estimates (upper triangle). Comparisons, column versus row, should be read from left, right. NA, not applicable.

### Appendix 18. Sensitivity analysis by only including studies with patients at high risk for severe complications

#### 18.1. Sensitivity analysis by only including studies with patients at high risk for severe complications for mortality

| **Baloxavir** | 0.99 (0.10 to 9.47) | NA | 2.99 (0.12 to 73.22) |
| --- | --- | --- | --- |
| 1.33 (0.16 to 11.39) | **Oseltamivir** | NA | 0.92 (0.21 to 4.01) |
| 0.62 (0.03 to 14.25) | 0.46 (0.04 to 5.75) | **Zanamivir** | 1.98 (0.26 to 15.24) |
| 1.22 (0.11 to 13.29) | 0.92 (0.21 to 4.01) | 1.98 (0.26 to 15.24) | **Standard care/placebo** |

Values are risk ratio (95% CI) for network estimates (lower triangle) and direct estimates (upper triangle). Comparisons, column versus row, should be read from left, right. NA, not applicable.

#### 18.2. Sensitivity analysis by only including studies with patients at high risk for severe complications for admission to hospital

| **Baloxavir** | 0.25 (0.03 to 2.23) | NA | 0.20 (0.02 to 1.70) |
| --- | --- | --- | --- |
| 0.28 (0.03 to 2.28) | **Oseltamivir** | NA | 0.65 (0.34 to 1.23) |
| 0.36 (0.03 to 4.37) | 1.28 (0.28 to 5.85) | **Zanamivir** | 0.50 (0.13 to 1.99) |
| 0.18 (0.02 to 1.46) | 0.65 (0.34 to 1.23) | 0.50 (0.13 to 1.99) | **Standard care/placebo** |

Values are risk ratio (95% CI) for network estimates (lower triangle) and direct estimates (upper triangle). Comparisons, column versus row, should be read from left, right. NA, not applicable.

#### 18.3. Sensitivity analysis by only including studies with patients at high risk for severe complications for time to alleviation of symptoms

| **Baloxavir** | NA | -0.36 (-1.97 to 1.25) | NA | NA | -1.07 (-2.66 to 0.52) |
| --- | --- | --- | --- | --- | --- |
| -0.57 (-2.82 to 1.68) | **Laninamivir** | 0.21 (-1.52 to 1.94) | NA | NA | NA |
| -0.36 (-1.80 to 1.08) | 0.21 (-1.52 to 1.94) | **Oseltamivir** | 0.38 (-0.92 to 1.69) | NA | -0.71 (-1.46 to 0.04) |
| 0.02 (-1.91 to 1.96) | 0.59 (-1.57 to 2.76) | 0.38 (-0.92 to 1.69) | **Peramivir** | NA | NA |
| -0.58 (-2.28 to 1.13) | -0.01 (-2.10 to 2.09) | -0.22 (-1.41 to 0.97) | -0.60 (-2.36 to 1.16) | **Zanamivir** | -0.49 (-1.42 to 0.43) |
| -1.07 (-2.50 to 0.36) | -0.50 (-2.38 to 1.38) | -0.71 (-1.46 to 0.04) | -1.10 (-2.60 to 0.41) | -0.49 (-1.42 to 0.43) | **Standard care/placebo** |

Values are mean difference (95% CI) for network estimates (lower triangle) and direct estimates (upper triangle). Comparisons, column versus row, should be read from left, right. NA, not applicable.

#### 18.4. Sensitivity analysis by only including studies with patients at high risk for severe complications for any adverse events

| **Baloxavir** | 0.89 (0.75 to 1.06) | NA | NA | 0.84 (0.71 to 1.00) |
| --- | --- | --- | --- | --- |
| 0.90 (0.77 to 1.06) | **Oseltamivir** | 0.86 (0.46 to 1.62) | NA | 0.93 (0.85 to 1.01) |
| 0.78 (0.40 to 1.50) | 0.86 (0.46 to 1.62) | **Peramivir** | NA | NA |
| 0.91 (0.74 to 1.13) | 1.01 (0.85 to 1.20) | 1.17 (0.61 to 2.26) | **Zanamivir** | 0.92 (0.79 to 1.06) |
| 0.84 (0.72 to 0.98) | 0.93 (0.85 to 1.01) | 1.08 (0.57 to 2.04) | 0.92 (0.79 to 1.06) | **Standard care/placebo** |

Values are risk ratio (95% CI) for network estimates (lower triangle) and direct estimates (upper triangle). Comparisons, column versus row, should be read from left, right. NA, not applicable.

#### 18.5. Sensitivity analysis by only including studies with patients at high risk for severe complications for adverse events related to treatments

| **Baloxavir** | 0.71 (0.48 to 1.05) | NA | 0.68 (0.46 to 1.00) |
| --- | --- | --- | --- |
| 0.67 (0.47 to 0.97) | **Oseltamivir** | NA | 1.07 (0.84 to 1.36) |
| 0.71 (0.41 to 1.25) | 1.06 (0.65 to 1.74) | **Zanamivir** | 1.00 (0.65 to 1.54) |
| 0.72 (0.50 to 1.03) | 1.07 (0.84 to 1.36) | 1.00 (0.65 to 1.54) | **Standard care/placebo** |

Values are risk ratio (95% CI) for network estimates (lower triangle) and direct estimates (upper triangle). Comparisons, column versus row, should be read from left, right. NA, not applicable.

#### 18.6. Sensitivity analysis by only including studies with patients at high risk for severe complications for serious adverse events

| **Baloxavir** | NA | -0.004 (-0.014 to 0.005) | NA | NA | -0.006 (-0.016 to 0.004) |
| --- | --- | --- | --- | --- | --- |
| 0.005 (-0.024 to 0.033) | **Laninamivir** | -0.010 (-0.037 to 0.017) | NA | NA | NA |
| -0.005 (-0.014 to 0.004) | -0.010 (-0.037 to 0.017) | **Oseltamivir** | -0.022 (-0.054 to 0.010) | NA | 0.001 (-0.006 to 0.009) |
| -0.027 (-0.061 to 0.006) | -0.032 (-0.074 to 0.010) | -0.022 (-0.054 to 0.010) | **Peramivir** | NA | NA |
| 0.002 (-0.015 to 0.019) | -0.003 (-0.034 to 0.029) | 0.007 (-0.009 to 0.023) | 0.029 (-0.007 to 0.065) | **Zanamivir** | -0.006 (-0.020 to 0.008) |
| -0.004 (-0.013 to 0.005) | -0.009 (-0.037 to 0.019) | 0.001 (-0.006 to 0.009) | 0.023 (-0.010 to 0.056) | -0.006 (-0.020 to 0.008) | **Standard care/placebo** |

Values are risk difference (95% CI) for network estimates (lower triangle) and direct estimates (upper triangle). Comparisons, column versus row, should be read from left, right. NA, not applicable.
